# The interplay between migration and selection on the dynamics of pathogen variants

**DOI:** 10.1101/2025.04.28.25326566

**Authors:** Wakinyan Benhamou, Rémi Choquet, Sylvain Gandon

## Abstract

The fitness advantage of an emerging pathogen variant is typically estimated from its change in frequency over time. This approach relies on the assumption that the pathogen spreads in a well-mixed population, where frequency changes are solely driven by selection. Yet, spatial structure can have major consequences on the spread of new variants. Here we model the change in frequency across time and across space of a new pathogen variant spreading in a two-patch host metapopulation. Crucially, we show that even small rates of migration can interfere with selection and may bias the estimation of fitness. We illustrate this effect with the evolution of SARS-CoV-2 by contrasting the spread of the Alpha and the Delta variants in England. We contend that the observed heterogeneity of fitness estimates across space could result from the influence of pathogen migration. This work highlights the need of a comprehensive theoretical framework accounting for the interplay between selection and migration on the spread of new pathogen variants.

## 1 Introduction

Estimating the fitness advantage of new pathogen variants is essential for evaluating epidemic risk and optimizing control strategies. This fitness advantage, relative to the co-circulating lineage, is usually estimated from the increase in variant frequency on the logit scale (aka the selection coefficient) [1, 2]. This approach has been widely used during the COVID-19 pandemic, in the context of the emergence and replacement of successive variants of concern [3–5]. For instance, **Fig. 1** shows the dynamics of the sweep of the Alpha variant (lineage B.1.1.7) and the Delta variant (lineage B.1.617.2) across regions in England. At the national level, the fitness estimated from the change in frequency is 0.065 day^−1^ (95% CI [0.052, 0.077]) for the Alpha variant (relative to the ancestral strain in October 2020) and 0.097 day^−1^ (95% CI [0.093, 0.101]) for the Delta variant (relative to Alpha). Zooming in, the analysis of the variant dynamics across space is particularly insightful. First, for both variants, the time of introduction of the new variant differs substantially among regions (see also **Fig. S1**), which generates major differences in variant frequency across space throughout their invasion. Second, the fitness estimations for Alpha remain relatively constant across regions, but is much more variable for Delta. In particular, the fitness seems to increase in regions where the variant was introduced later (e.g, about 60% higher in the Yorkshire and Humber region than in the London or South West regions). How can we explain this spatial variation in fitness among different regions? This pattern could emerge if there were systematic sampling biases as pointed out in [4] and [6]. Yet, we want to explore the alternative mechanistic possibility that this spatio-temporal pattern is driven by other evolutionary forces acting on the change in frequency of a new variant. First, demographic stochasticity and superspreading events may have massive effects on the dynamics of a new variant at the start of its invasion, when its population size is relatively small [7–9]. These stochastic events are likely to explain the difference in introduction time among different regions (random founder effect) but should have relatively minor effects on the subsequent dynamics of the variant. Second, the new variant may acquire additional beneficial mutations during its invasion, but this explanation is very unlikely given the speed at which the Delta variant went to fixation in England (less than two months). Third, regional differences in selective pressures could also result in variations in frequency changes over time (e.g., heterogeneity in non-pharmaceutical interventions (NPIs) [10–12] or in vaccination [13–15]). Last, host mobility among regions (allowing pathogen migration) may also alter the dynamics of variant frequency. In this study, we will focus on the influence of pathogen migration and we thus need to develop a model accounting for spatial structure, epidemiological dynamics and pathogen evolution.

**Figure 1:**
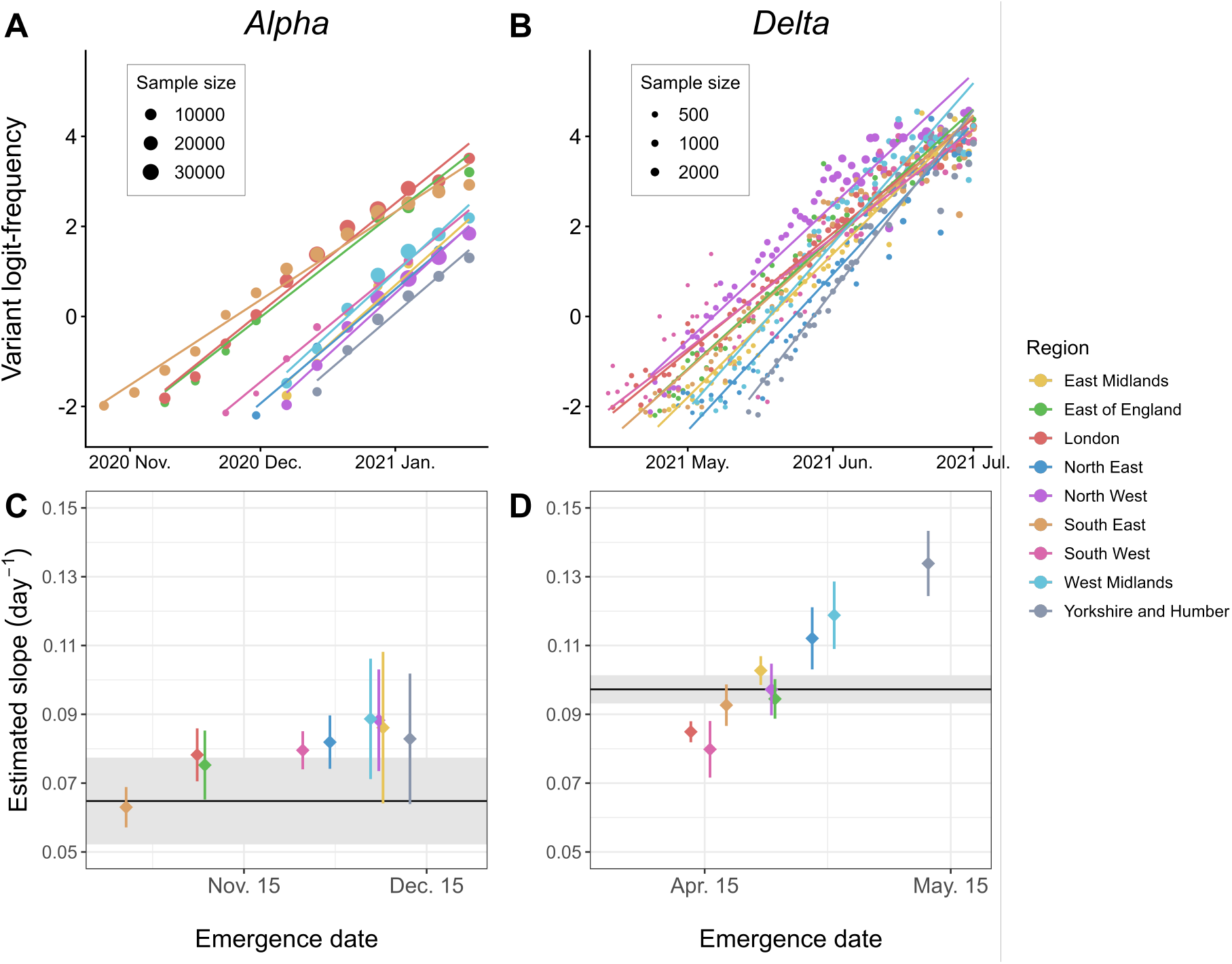
Growth of the Alpha and Delta variants in England. Time series data (points) and linear regressions (lines) of the logit-frequency of two SARS-CoV-2 variants: (A) Alpha (lineage B.1.1.7) and (B) Delta (lineage B.1.617.2). Data for Alpha correspond to time series of regional frequencies of S Gene Target Failure (qPCR performed after swab sampling in the wider population) in England from [96]; data for Delta were shared by Erik Volz who previously used them in [4]. To minimize the influence of demographic stochasticity, we remove all frequencies lower than 0.1 or greater than 0.99. The second row (C-D) displays the estimated values of the slope of each linear regression (mean (diamonds) *±* 95% confidence interval (segments), expressed per day) vs. emergence date (first date the frequency has reached 0.1). These slopes are typically used to estimate the selection coefficient of a variant. The horizontal line and the gray envelope represent the same estimation but for data aggregated at the national scale (no spatial structure). We plot the distributions of intervals between emergence dates in **Fig. S1**.

Classical epidemiological models, such as the Susceptible-Infected-Recovered (SIR) model popularized by [16], assume a homogeneous-mixing population: each individual has the same probability of meeting any other individual [17–19], so that there is no spatial heterogeneity that may impact the spread of the disease. Yet, hosts typically interact more with a reduced set of neighbors [20, 21]. A common way to account for spatial structure is to use metapopulation models, that is, a network of spatially separated patches (e.g., households, cities, regions, countries), interconnected by explicit flows of migration [22, 23]. Such models have been extensively used in epidemiology (e.g., [24–32]), and the recent COVID-19 pandemic was no exception (e.g., [33, 34]). Modeling spatial heterogeneity in selection and migration have also been a major focus in the field of evolutionary biology [35–37], including in the context of host-pathogen systems, focusing for example on life-history traits such as drug resistance [38, 39] or virulence and transmission [15, 40–49]. Yet, most studies focus on the long-term evolutionary outcome and the analysis of the interplay between migration and selection on the transient evolutionary dynamics of pathogens remains unclear.

Evolutionary epidemiology theory is well suited to study the short-term dynamics of host-pathogen systems, when epidemiological and evolutionary time scales overlap [50], but has been mostly used in well-mixed environments. In this study, we extend evolutionary epidemiology theory to follow the evolutionary dynamics of a pathogen population in a two-patch host metapopulation. This fully deterministic model tracks simultaneously the epidemiological dynamics and the change in frequency of a new pathogen variant. We focus on the dynamics of the variant frequency across time and on the differentiation across populations, two quantities that are increasingly becoming available from sequencing data. Our model thus provides a useful theoretical framework to build a more comprehensive understanding of the evolutionary forces that shape the dynamics of emerging pathogen variants.

## 2 Evolutionary epidemiology across time and space

### 2.1 The model

We model the spread of a horizontally transmitted pathogen in a two-patch host metapopulation using an SIRS model. Let *β* refer to the *per capita* transmission rate, *γ* to the *per capita* recovery rate and *ω* to the *per capita* waning rate of immunity. We track the dynamics of two strains of the pathogen: the wildtype (*w*) and the mutant strain (*m*, or variant). The variant may differ phenotypically from the wildtype in terms of transmission rate (*β*_*m*_ = *β*_*w*_ + Δ*β*) and/or recovery rate (*γ*_*m*_ = *γ*_*w*_ + Δ*γ*). We further assume no coinfection and full cross-immunity. Besides, hosts are distributed in two populations, labeled *A* and *B*, and coupled by migration. Throughout, we use superscripts to distinguish the two populations. We denote *N*^*A*^ and *N*^*B*^, the total densities of populations *A* and *B*, respectively, such that *N*^*A*^+*N*^*B*^ = *N*. Define *µ*^*i*^, the probability for a host from the focal population *i* ∈ {*A, B*} to visit the non-focal population. For the sake of simplicity, we assume that *µ*^*i*^ is independent of both the host’s epidemiological status (*S, I* or *R*) and the pathogen’s strain (*w, m*). Visits are assumed to be only instantaneous to-and-fro host movements, such as commuting [19, 26]. The force of infection is thus the sum of four types of intra- or inter-community interactions. Last, due to potential heterogeneities between the two populations (e.g., local NPIs or behaviors), we also allow phenotypic traits *β* and *γ* to vary between populations, so that the variant phenotypes in population *i* are given by 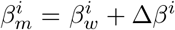 and 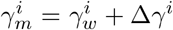. **Fig. 2** provides an illustration of the model and notations are summarized in **Table 1** (more details are available in **SI Appendix §S1-S2**).

**Table 1:** Notations. The subscript *k* ∈ {*w, m*} refers to the strain of the pathogen: wildtype strain *w* or the mutant strain *m* (or variant). The superscript *i* ∈ {*A, B*} refer to the focal population *i*.

| Term | Definition |
| --- | --- |
| $N, N^i$ | Total population densities (global and local, respectively) |
| $S, S^i$ | Susceptible hosts (global and local, respectively) |
| $I, I^i$ | Infected/infectious hosts (global and local, respectively) |
| $I_k, I_k^i$ | Hosts infected by strain $k$ (global and local, respectively) |
| $R, R^i$ | Recovered hosts (global and local, respectively) |
| $q, q^i$ | Frequency of the variant $m$ among infected hosts:<br>$q = I_m/I$ (global) and $q^i = I_m^i/I^i$ (local) |
| $\beta_k^i$ | *Per capita* transmission rate of strain $k$ in population $i$ |
| $\gamma_k^i$ | *Per capita* recovery rate of strain $k$ in population $i$ |
| $\omega$ | *Per capita* waning rate of immunity |
| $\mu^i$ | Probability for hosts from population $i$ to commute to the other population |
| $\Delta\beta^i, \Delta\gamma^i$ | Phenotypic differences between the variant $m$ and the wildtype $w$ in population $i$:<br>$\Delta\beta^i = \beta_m^i - \beta_w^i$ and $\Delta\gamma^i = \gamma_m^i - \gamma_w^i$ |
| $\mathcal{S}^i$ | Effects of local selection and interaction between selection and migration<br>on the dynamics of $q^i$ (variant frequency in population $i$) |
| $\mathcal{H}^i$ | Effect of between-population homogenization on the dynamics of $q^i$ (variant<br>frequency in population $i$) |
| $\mathcal{Q}$ | Spatial differentiation (between populations $A$ and $B$):<br>$\mathcal{Q} = q^A / (1 - q^A) \times (1 - q^B) / q^B$ |

**Figure 2:**
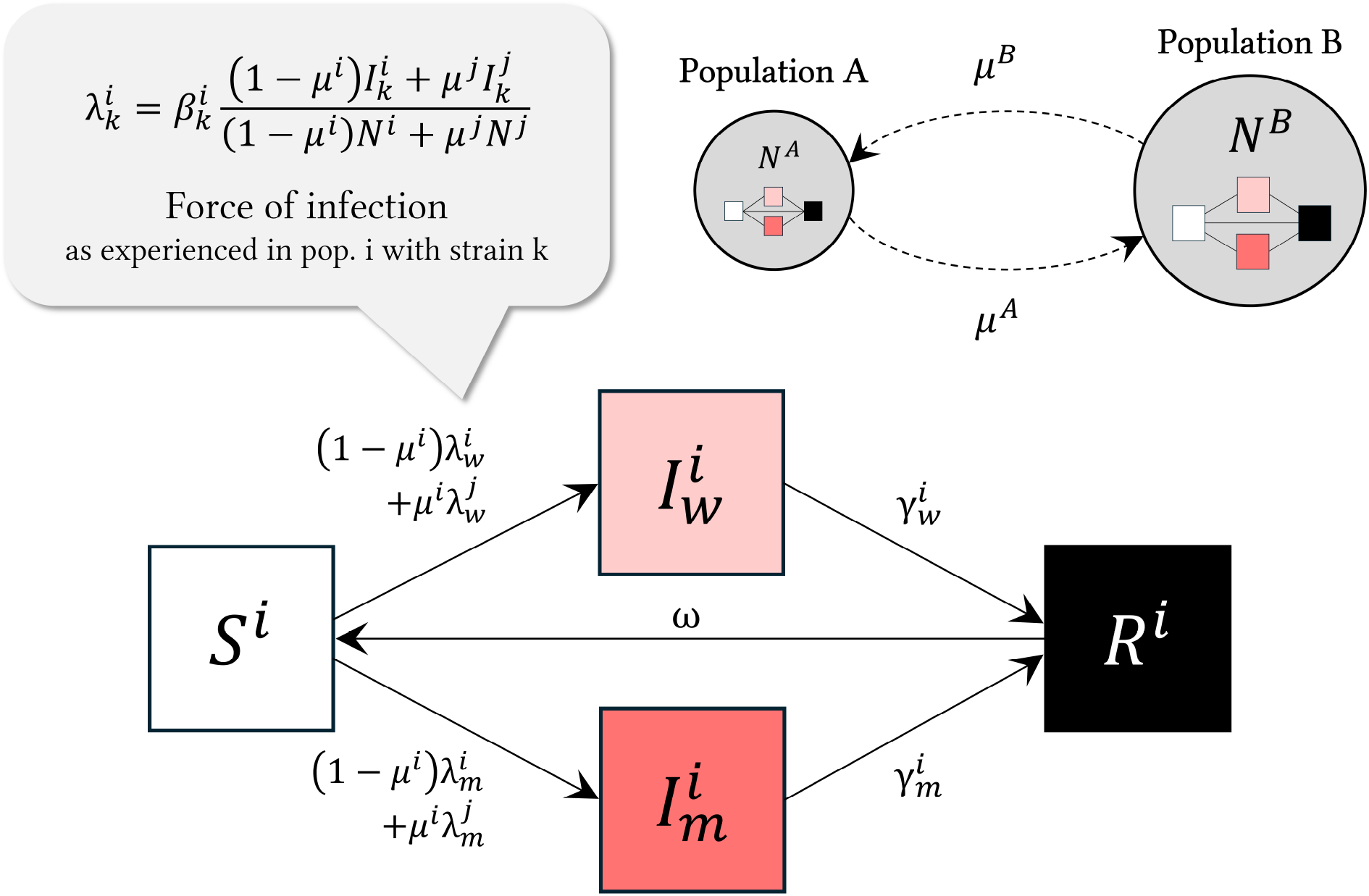
Schematic presentation of the compartmental model. (Top right) We model a two-patch host metapopulation where individuals from population *A* (resp. *B*) may instantaneously visit population *B* (resp. *A*) with probability *µ*^*A*^ (resp. *µ*^*B*^), or remain in their home population with complementary probabilities; circle sizes represent population densities, *N*^*A*^ and *N*^*B*^ (here for example, *N*^*B*^ *> N*^*A*^). (Bottom) SIRS model for the focal population *i* ∈ {*A, B*} (accordingly, the non-focal population is denoted *j*); subscripts *w* and *m* refer to the wildtype and mutant strain (or variant) of the pathogen. (Top left) The force of infection 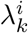 represents the rate at which susceptible hosts are infected in population *i* ∈ {*A, B*} with strain *k* ∈ {*w, m*}. See **Table 1** for notation and **SI Appendix §S1–S2** for more details.

### 2.2 Dynamics of the variant frequency across time

To follow pathogen evolution, we track 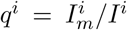, the local frequency of the variant in each population *i* ∈ {*A, B*} (we drop the time dependence in our notations for compactness and readability). Next, we assume that probabilities *µ*^*A*^ and *µ*^*B*^ are relatively small because contact rates are likely to be much higher within than between populations. Under this assumption, we show in **SI Appendix §S4** that the temporal dynamics of the logit-frequency of the variant logit (*q*^*i*^) can be approximated at the first order by

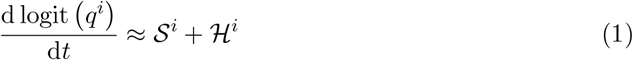

with

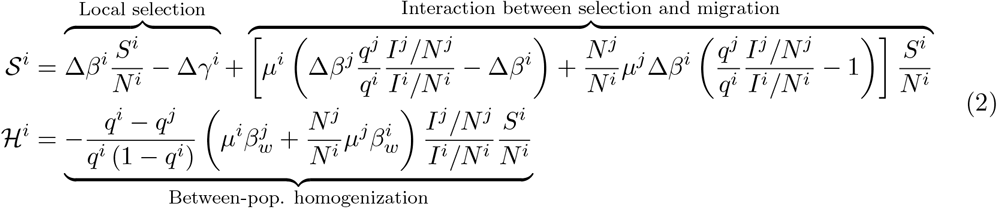

Note that we obtain a similar partitioning following an approach analogous to [51] (**SI Appendix §S5**) and that these equations can be generalized to an arbitrary number of populations (**SI Appendix §S6.1** and **Fig. S2**). Equations (1)-(2) are useful to better understand how selection and migration shape the evolution of the local frequency of the variant. First, 𝒮^*i*^ collects all the terms that depend on the phenotypic differences between the two strains and refer to the effects of local selection on the focal population as well as the interaction between selection and migration between populations. Second, the component ℋ^*i*^ describes a purely neutral process that affects the local dynamics of the variant as soon as the variant frequency varies between the two populations (i.e., *q*^*i*^≠ *q*^*j*^).

As expected, in the absence of host mobility (*µ*^*i*^ = *µ*^*j*^ = 0), we recover the single-population scenario: 𝒮^*i*^ = Δ*β*^*i*^*S*^*i*^*/N*^*i*^ − Δ*γ*^*i*^ and ℋ^*i*^ = 0, where the dynamics of the variant logit-frequency is solely governed by the local selection and thus depends only on the phenotypic differences between the two strains and on the local availability of susceptibles [10, 12, 52, 53]. More generally, if the two strains differ in terms of transmission, 𝒮^*i*^ depends on potential asymmetries in population size and prevalence between the two populations because migration will allow infections by individuals from other populations. This interaction term between selection and migration accounts for (i) local susceptibles who move out and get infected away by non-local hosts and (ii) non-local hosts who move in and infect local hosts. Note that this interaction may arise even when the variant frequency is identical across populations (see also [52, 54, 55]).

When the local variant frequency varies across populations, the direction of ℋ^*i*^ is opposite to the sign of the difference *q*^*i*^ − *q*^*j*^ and its strength is driven by the intensity of pathogen migration (i.e, inter-community transmission), which depends on host mobility (governed by *µ*^*i*^ and *µ*^*j*^), transmission rates, spatial asymmetries in prevalence and population size and on the local availability of susceptible hosts. Interestingly, ℋ^*i*^ is inversely proportional to the local genetic variance *q*^*i*^ (1 − *q*^*i*^). Therefore, even with very low rates of migration, ℋ^*i*^ cannot be neglected when the genetic variance is very small, which is typically what may happen just after the introduction of a new variant in the focal population (**Fig. 3** and **S3**). The effects of migration are magnified when the non-focal population is bigger (**Fig. S4-A**). Even in the absence of selection (𝒮^*i*^ = 0), migration can still lead to a transient increase of the variant frequency until it reaches an equilibrium where *q*^*i*^ = *q*^*j*^ (**Fig. S4-B**).

**Figure 3:**
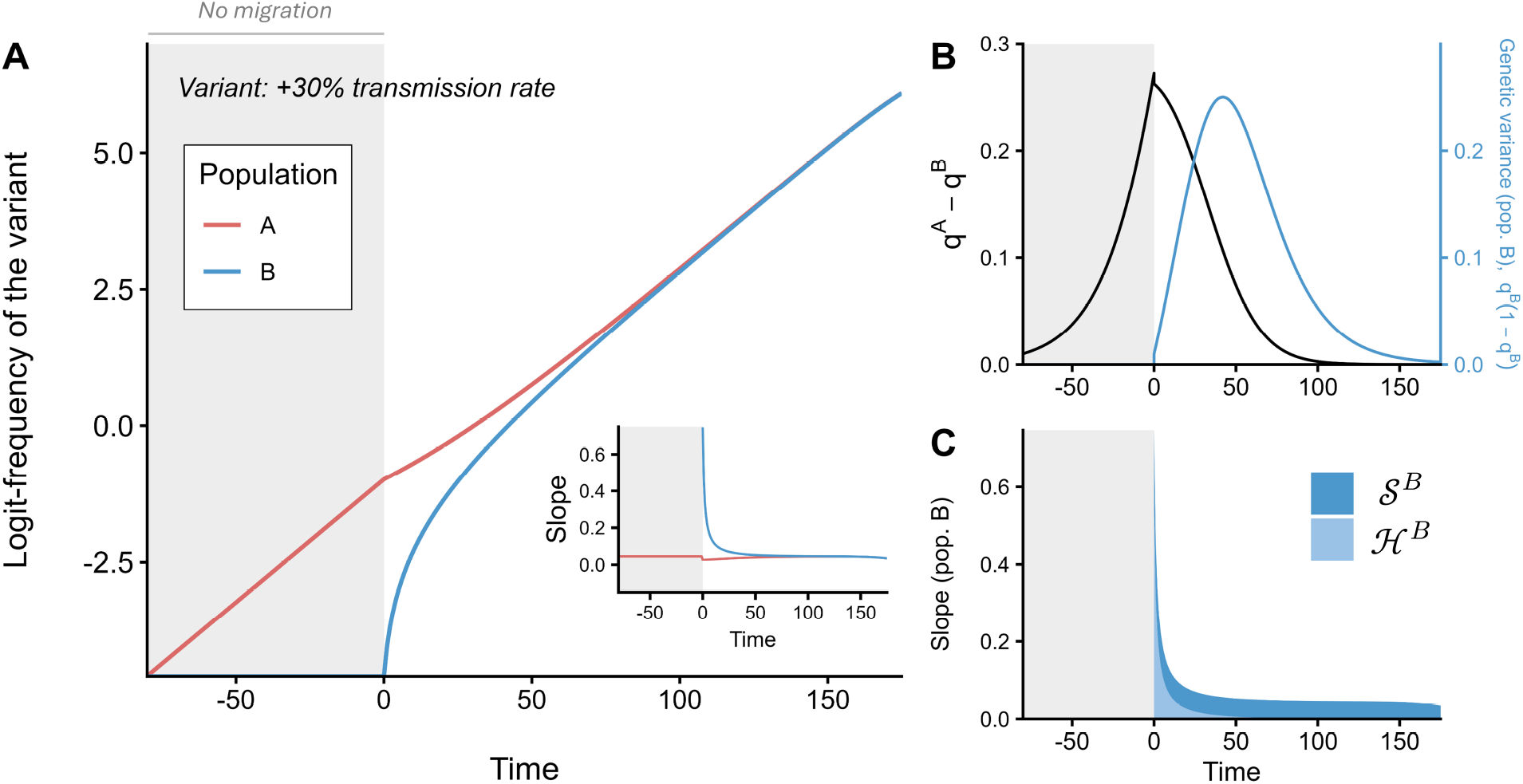
Migration can interfere with selection and change the local dynamics of the frequency of an emerging variant. The host metapopulation is divided in two populations, *A* and *B*. The variant has a +30% transmission rate selective advantage over the wildtype. We simulate the model with identical total population densities (*N*^*A*^ = *N*^*B*^ = 100), and parameter values (same for both populations): *β*_*w*_ = 0.15, *β*_*m*_ = 0.195, *γ*_*w*_ = *γ*_*m*_ = 0.1 and *ω* = 0. The wildtype is initially introduced at very low density (10^−7^) in both populations and the variant is only introduced in pop. *A* such that it represents 1% of the infected hosts *I*^*A*^ – both populations are otherwise fully susceptible. Before *t* = 0, the two populations are isolated (*µ*^*A*^ = *µ*^*B*^ = 0, gray background). At *t* = 0, the variant is introduced in pop. *B* (1% of *I*^*B*^) and, from that time point onward, hosts from pop. *B* may visit pop. *A* (*µ*^*B*^ = 0.1). We plot over time the dynamics of: (A) the variant logit-frequency in pop. *A* (red) and *B* (blue) – the inset plot shows the approximation (1)-(2) (see also **Fig. S3**) –, (B) the frequency difference *q*^*A*^ − *q*^*B*^ (black) and genetic variance in pop. *B* (blue) and (C) the slope of the variant logit-frequency in pop. *B* decomposed into 𝒮^*B*^ (darker blue) and homogenization *H*^*B*^ (lighter blue) terms. Crucially, migration steepens the change of the local frequency of the emerging variant in the population where it is introduced later. Note that epidemiological dynamics (not shown) exhibit only little variations over the simulation period.

### 2.3 Dynamics of the differentiation across space

In this section, we examine the dynamics of the genetic differentiation across space – hereafter referred to as spatial differentiation –, which provides another way to examine the interplay between migration and selection. A simple way to quantify the spatial differentiation is to compute the difference *q*^*i*^ − *q*^*j*^ which appears in ℋ^*i*^ in (2). Yet, the dynamics of this measure of spatial differentiation is more difficult to interpret because it depends on the local genetic variances which are also dynamical variables (see **SI Appendix §S4.2.3**).

In the following, we focus on an alternative measure of differentiation [12, 56]

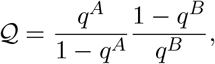

such that ln (𝒬) = logit (*q*^*A*^) − logit (*q*^*B*^). Hence, when the variant frequency is the same in both populations (no differentiation), 𝒬 = 1 and ln (𝒬) = 0. Under the assumption of weak migration, ln (𝒬) can be approximated by (see **SI Appendix §S4.2.3**)

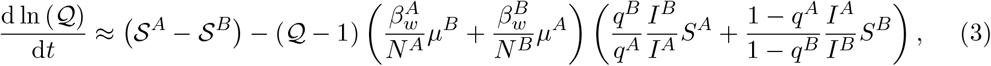

along with the initial condition 𝒬 (*t* = 0) = 𝒬_0_. The above dynamical equation shows that the spatial differentiation is driven by two evolutionary forces. The first, 𝒮^*A*^ − 𝒮^*B*^, is the difference in selection between populations *A* and *B*. The second is a decay term that tends to homogenize the frequency of the variant across populations. This second term depends on the current level of differentiation (𝒬 − 1) and on the amount of migration. In the neutral case (i.e., no selection, 𝒮^*A*^ = 𝒮^*B*^ = 0), 𝒬 converges towards unity (no differentiation). Next, to better understand the interplay between these two terms we examine the dynamics of differentiation under different scenarios.

First, let’s assume for simplicity that selection is homogeneous across space (i.e., 𝒮^*A*^ = 𝒮^*B*^). As in **Fig. 1** and **Fig. 3**, an initial spatial differentiation 𝒬_0_ can be due to different times of introduction of the variant in each population. This can happen for instance when two populations, only one harboring the variant, were previously isolated (e.g., lockdown, travel ban), and/or because of the inherent stochasticity of the introduction and emergence of a rare variant. In particular, longer delays between different introductions of the variant will result in a larger initial spatial differentiation 𝒬_0_. The spatial differentiation can only persist over time in the absence of migration between the two populations (**Fig. S5-A**). However, as soon as there is some migration, differentiation decays with time and eventually vanishes (**Fig. S5-C** and **E**).

Second, if selection varies across space it may counteract the homogenizing effect of migration. First, note that selection can vary across space even when the phenotypic differences are the same in both populations (Δ*β*^*A*^ = Δ*β*^*B*^ = Δ*β* and Δ*γ*^*A*^ = Δ*γ*^*B*^ = Δ*γ*). In particular, when Δ*β*≠ 0, a variation in the prevalence or in the availability of susceptible hosts between the two populations can generate a transient dynamics of 𝒬, even without pre-existing spatial differentiation (ln (𝒬_0_) = 0) (**Fig. S6**). Second, selection can vary across space when the effect of the mutation vary across populations (i.e., Δ*β*^*A*^ ≠ Δ*β*^*B*^ and/or Δ*γ*^*A*^≠ Δ*γ*^*B*^). In this scenario, selection can generate and maintain the differentiation 𝒬, whose dynamics depends on the balance between selection and migration (**Fig. S5**, second column). In particular, 𝒬 may reach a quasi-equilibrium value when migration is large enough relative to selection (see **Fig. S5-F** and **SI Appendix §S4.2.3**). If the variant is always selected for, it will eventually be fixed in both populations. In contrast, if the variant is adapted in one population but maladapted in the other (i.e., local adaptation), a stable level of differentiation can be maintained in the long term when migration is not too strong as it erodes local adaptation [57–59] (**Fig. S7**). The variant may thus persist at an intermediate frequency without going to fixation and coexist with the wildtype (polymorphism maintenance).

## 3 Migration can bias estimates of selection

We conduct a simulation study to illustrate and quantify the extent to which ignoring migration can bias estimates of selection. In **Fig. 4**, we simulate trajectories of the variant logit-frequency in a two-patch host metapopulation using a selection coefficient 𝒮^*A*^ = 𝒮^*B*^ = 0.1 day^−1^, which is around the value we estimated for Delta at the national level (**Fig. 1-D**). As in **Fig. 3**, the variant is initially introduced in population *A* (initially epidemiologically isolated from population *B*). At *t* = 0, the two populations become connected and the variant is introduced in population *B*. We vary the migration probability *µ* (assuming for simplicity *µ*^*A*^ = *µ*^*B*^ = *µ*) and the population sizes *N*^*A*^ and *N*^*B*^. We also vary the initial value of the spatial log-differentiation ln (𝒬_0_) from 0 to 3 (similar to the range observed for Alpha and Delta in **Fig. 5**). For each set of parameters, we track the frequency of the variant in the metapopulation (*q*) and in each population (*q*^*A*^ and *q*^*B*^) and generate synthetic data assuming observation errors to be normally distributed with mean 0 and standard deviation 0.25. We then estimate the relative fitness of the variant from the observed change in the variant frequency using linear regressions and compute the corresponding relative error with respect to its true value (**Fig. 4**).

**Figure 4:**
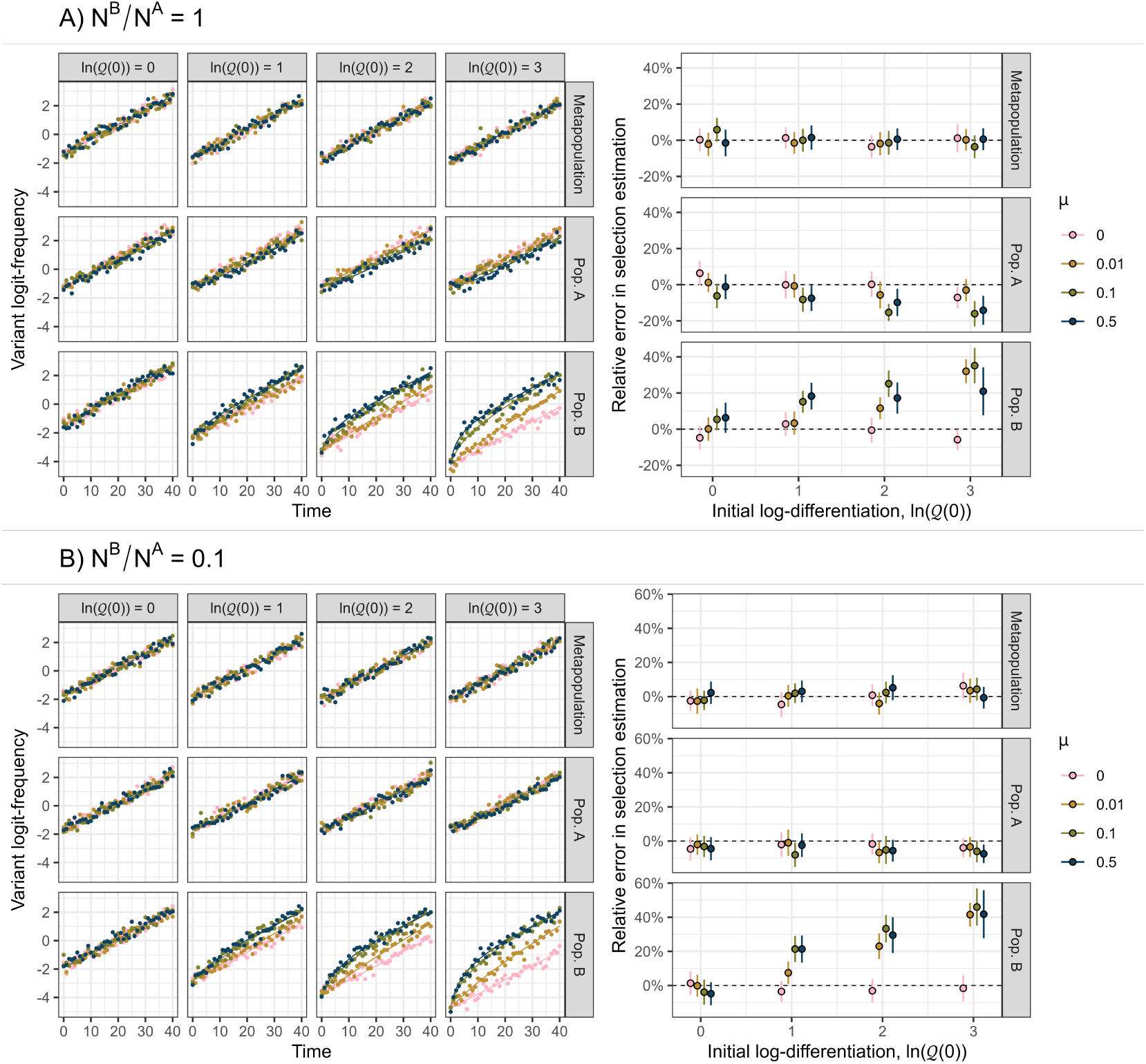
Migration can bias local estimates of selection. For the sake of simplicity, we assume that both strains share the same transmissibility and only differ in terms of recovery rates, so that we can set aside the dynamical effects in the selection coefficient. We further assume that phenotypic traits do not vary across populations. The relative fitness of the variant reduces thus to − Δ*γ*. We simulate the model with (A) identical population sizes (*N*^*A*^ = *N*^*B*^ = 100) or (B) asymmetry in population size (*N*^*A*^ = 100, *N*^*B*^ = 10), varying the initial value of the log-differentiation ln (𝒬 (0)) from 0 to 3 and the migration probability *µ* (assuming *µ*^*A*^ = *µ*^*B*^ = *µ*) from 0 to 0.5. For all simulations, we take: *β*_*w*_ = *β*_*m*_ = 0.15, *γ*_*w*_ = 0.2, *γ*_*m*_ = 0.1 (+100% duration of infectiousness) and *ω* = 0. At *t* = 0, the variant represents about 16% of infections in pop. *A*. From the simulated trajectories of the variant logit-frequency (lines), we generated simulated data (points) by adding Gaussian noise with standard deviation 0.25. For each simulated dataset, we estimate the selection coefficient by fitting a linear regression and compute the relative error of the estimated slope with respect to the true selection coefficient 𝒮^*i*^ = −Δ*γ* = 0.1. Note how local estimates of selection can differ from the true coefficient of selection. In contrast, the estimates based on the mean change of frequency is not biased. *q*

**Figure 5:**
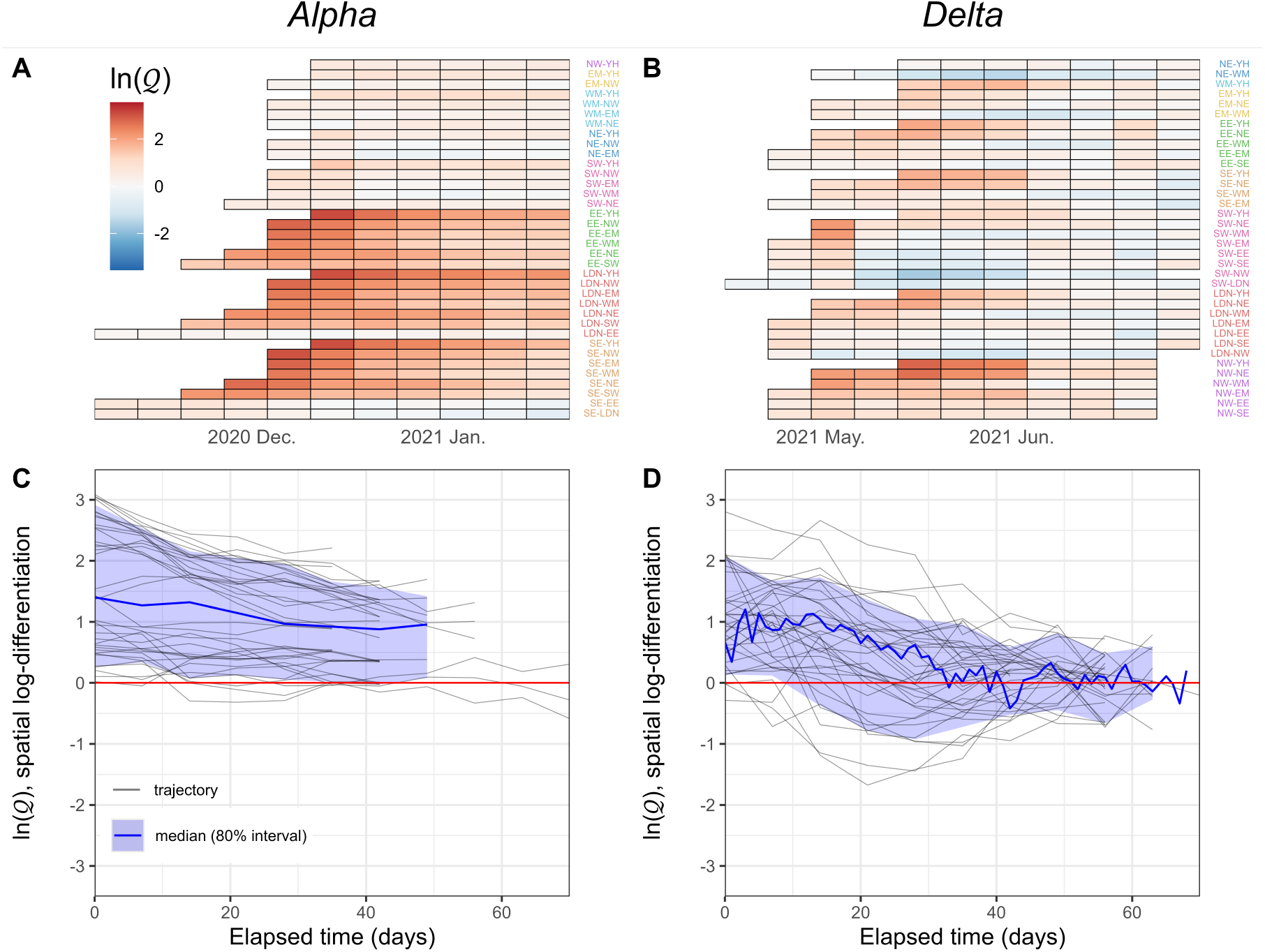
The decay of spatial differentiation over time for the Alpha and Delta variants in England. Using the regional time series of the Alpha and Delta variant logit-frequency in the 9 regions of England presented in **Fig. 1**, we compute the 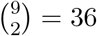 pairwise regional differences for each available time point (log-differentiation ln (𝒬) = logit (*q*^A^) *−* logit (*q*^*B*^)). We choose the pairs so that the early values of ln (𝒬) are positive (see **Fig. S8**). Computed values are plotted (A-B) by date and region pair (colored according to the first region) and (C-D) against the time elapsed since the first value of each region pair. The thick black line represents the mean and the envelopes the empirical interval between the 10% and 90% quantiles (only computed with at least 10 values). The horizontal red line indicates the absence of differentiation (ln (𝒬) = 0). For better visualization, we use weekly data for the Delta variant. Initial spatial differentiation values are usually higher for the Alpha variant and, for both variants, the dynamics of the spatial log-differentiation seems to be converging towards 0, suggesting no or little spatial heterogeneity in selection. Abbreviations: East Midlands (EM), East of England (EE), London (LDN), North East (NE), North West (NW), South East (SE), South West (SW), West Midlands (WM) and Yorkshire and Humber (YH).

As explained above, the change in frequency can be affected by the spatial structure and can bias the estimated relative fitness of the new variant. For instance, with *µ* = 0.1, the magnitude of the error can reach 40-50% when ln (𝒬_0_) = 3. Relative errors are either similar or smaller with a 10-fold decrease in *µ* and are relatively similar with a 5-fold increase in *µ*. Note that the bias can be even larger if the focal populations is coupled with larger (or many) populations. Importantly, starting from lower levels of spatial differentiation helps to mitigate this bias with almost no relative errors higher than ≈ 20% for ln (𝒬 (0)) ≤ 1; and the estimation of the relative fitness at the metapopulation level using the average frequency *q* yields here unbiased estimates of the selection coefficient.

## 4 Discussion

Gene flow is a potent evolutionary force that tends to homogenize genetic variability across space via migration. It usually slows down adaptation because it counteracts the effect of local selection, but it may also promote adaptation when it introduces adaptive mutations [60]. Unraveling the tangled contributions of different evolutionary forces to the changes in frequency can be carried out using genomic time series [61]. Here we study the transient evolutionary dynamics of a pathogen population in a host metapopulation to understand the interplay between selection and migration on the dynamics of different strains of pathogens across time and space.

### 4.1 Migration and selection jointly shape the dynamics of new variants

Neglecting spatial heterogeneity can bias the estimation of key epidemiological parameters in monomorphic pathogen populations, such as the basic reproduction number [62–65]. Here we show that in a polymorphic pathogen population migration can also interfere with the effect of selection, thereby introducing a bias in the estimation of the fitness of a new variant. In particular, the slope of the logit-frequency of a new variant (used as an estimation of its selection coefficient) can become locally very steep during the emergence of a variant because of the effect of migration from another population. This bias can be mitigated when coupled populations are not or weakly differentiated or when frequency data are aggregated at the metapopulation level, which requires in both cases some prior knowledge of the underlying population-interaction network (who is connected to whom). While focusing on the metapopulation level seem to be an effective approach, it may also not be relevant if selection is heterogeneous across space or in the presence of immigration events from outside the focal metapopulation. This highlights the need to identify the appropriate spatial scales (e.g., cities, regions, countries) for estimating selection, which in practice can only be determined on a case-by-case basis, depending on the available data and the questions of interest.

### 4.2 Insights from the spread of the Alpha and Delta variants in England

Understanding the interplay between selection and migration yields some important insights on the spread of SARS-CoV-2 variants. In particular, the analysis of the dynamics of the spatial differentiation can help explain the difference between the Alpha and the Delta variants in England (**Fig. 1**).

First, the computation of pairwise measures of differentiation across the regions of England yields higher initial values for the Alpha variant than for the Delta variant (**Fig. 5**). As discussed above, higher levels of initial differentiation 𝒬_0_ could be due to longer delays between successive introductions for Alpha (**Fig. S1**), which can be induced by a lower degree of connectivity between regions, i.e., fewer migration events (see also [66]). Second, for both variants, spatial differentiation seems to converge towards 0 (log scale) over time, suggesting no or little spatial heterogeneity in selection (as in **Fig. S5-C** and **E**). Third, the speed of decay of spatial differentiation with time may also reveal the amount of connectivity among regions. We show in **Fig. S8-A** and **B** that the early rate at which the differentiation ln (𝒬) decays depends both on the initial amount of differentiation and the amount of migration. We apply linear regressions to the beginning of the dynamics of the spatial differentiation shown in **Fig. 5** to compare initial differentiation values (intercept) and early decay rates (slope) of both variants. Interestingly, the patterns we obtain for SARS-CoV-2 are consistent with the hypothesis that migration was higher during the sweep of Delta because, for similar initial values, the drop in differentiation tends to be steeper than for the Alpha variant (**Fig. S8-C**).

The hypothesis that contrasted amounts of migration may drive the different patterns observed for the two variants is further supported by the increase in human mobility during the sweep of Delta compared to Alpha according to Google mobility reports [67] (**Fig. S9**). Although Google mobility reports do not explicitly capture inter-region mobility, they reflect the progressive relaxation of NPIs throughout the pandemic. Additionally, the analysis of neutral allele frequency time series have also been used very recently to estimate inter-community transmission rates, confirming the higher mixing rates during the spread of the Delta variant than during the spread of the Alpha variant [68]. This analysis further shows that the reproductive values (i.e. the relative contribution to the future of the pathogen population) vary among regions and across time. From the pathogen point of view, these reproductive values measure the relative “quality” of the different regions [55, 69]. Weighting the change in variant frequency by the reproductive values of different regions may thus provide an alternative way to model the heterogeneous and time-varying contribution of the different regions (i.e., source-sink dynamics) to the evolution of the pathogen [55, 70].

### 4.3 Limitations, perspectives and conclusion

Our model relies on numerous simplifying assumptions. For example, we assume that all hosts from the same population can commute with the same probability, regardless of whether they are infected or not. We thus neglect the potential influence the pathogen, or each strain, may have on the mobility of its host – e.g., if hosts tend to limit their movements when they present symptoms or are aware of being infected. In **SI Appendix §S6.2** and **Fig. S10**, we relax the assumption that infected hosts can commute with the same probability as non-infected hosts. We show that in most cases this assumption has little impact on the evolutionary dynamics of the variant, in particular as the infected fraction of the population is here very small compared to the fraction of susceptibles. This effect could be even less pronounced in the presence of asymptomatic transmissions, like for SARS-CoV-2 [71]. We also model host mobility as instantaneous to-and-fro host movements, which is a crude approximation and may not reflect realistic travel flows.

While our model is a first step to take spatial structure into account, it remains an overly simplified representation of real-world spatial and social networks, which are characterized by complex and time-varying interaction processes [72]. Continuous-space models (e.g., reaction-diffusion models [73, 74]) could for example capture finer spatial heterogeneity and diffusion dynamics (more realistic for systems without clear patch boundaries); they could thus offer complementary insights and help assess the robustness of our findings. More sophisticated models usually offer a better depiction of the reality, but general conclusions are also more difficult to draw, as they are also inherently less tractable [29]. Nonetheless, our parsimonious framework can be extended to an arbitrary number of populations (see **SI Appendix §S6.1** and **Fig. S2**), and migration probabilities can be calibrated to capture more complex scenarios – such as in, e.g., gravity models (from transportation theory [75]), which assume transmissions to be correlated positively with population sizes and negatively with separating distances [76–78].

In our model, transmission rates depend on the population where the infection occurs, not on the population to which the infected host belongs. This is suitable to model scenarios with, e.g., differences in control strategies or behavioral differences where hosts align with local habits, but not for scenarios where transmission is an intrinsic property of the host (e.g., transmission-blocking vaccine). In contrast, population differences in recovery rates can be due to, e.g., differences in treatment/vaccination which affect the infection clearance. Note that spatial heterogeneity in host immunity profiles (natural or vaccine-induced) can also induce a heterogeneous selection on other phenotypic traits and favor immune-escape variants [14, 15, 54, 79, 80]. In **SI Appendix §S6.3**, we extend our model to account for the ability of the variant to partially escape the immunity provided by the wildtype. This extension affects the selection component 𝒮^*i*^ and, as expected, the local selection of the variant increases since it has access to a larger proportion of the host population than the wildtype.

Though beyond the scope of this paper, demographic stochasticity can also affect the dynamics of pathogen variants. For instance, there was no clear consensus early in the COVID-19 pandemic on whether the global increase in the frequency of SARS-CoV-2 substitution D614G was due to positive selection or to a random founder effect [7, 8, 81–83], underscoring the complexity of disentangling the effects of selection from genetic drift. Our deterministic model is not meant to capture the very early stage of the invasion of a new variant because the finite number of hosts infected by the new variant is likely to lead to the extinction of the variant. Stochastic models are needed to quantify the probability of emergence during the onset in the invasion of a new variant [84, 85].

Last, although we use empirical time series from SARS-CoV-2 as a motivation and illustration of this work, we did not statistically fit the model to the data. This is a challenging task that would require additional assumptions and information, such as regional incidence or prevalence data to account for the spatial distribution of susceptible and infected hosts. We would also need to incorporate the matrix of (potentially time-varying) migration probabilities to account for host mobility. Human traveling behavior can be quantified with increasingly accuracy at different scales – e.g., travel history data [86, 87], air-traffic data [31, 88], cell-phone mobility data [34, 89] –, yielding independent measures of the intensity of host mobility. Alternatively, the analysis of neutral allele frequency time series can be used to estimate the proportion of infections imported from another population [68]. And phylogeographic analyses, by reconstructing the ancestral geographic locations of a viral phylogenetic tree, can also be used for the inference of migration events independently of natural selection (under the assumption that the studied genetic markers are neutral). More broadly, phylogenetic epidemiology (phylogeographic and phylodynamic approaches) enables to estimate epidemiological and evolutionary parameters from genomic data, such as pathogen reproduction numbers, growth rates and spatial dispersal (see e.g., [8, 86, 89–93]). These recent inference methods rely on the reconstruction of viral phylogenies from sampled sequences to infer the underlying population processes (and can also be extended to integrate auxiliary sources of information like mobility data [86]). Our mechanistic framework thus provides a complementary tree-free approach to data-driven phylogenetic analyses.

Our analysis aims to untangle how migration and selection jointly shape the spread of an emerging variant. We show that the dynamics of variant frequency and spatial differentiation contain valuable information about pathogen evolution. More generally, this underscores the importance of the distribution of variant frequency among different “classes” of hosts that may belong to different regions or to other types of class-structure (e.g., age or social group, vaccination status). Hence, it would be interesting to explore further how such stratified data may yield more quantitative approaches to disentangle the effect of selection from the effects of migration among different classes of hosts, and thus refine the estimation of variant fitness.

## Data Availability

Data that were used in this study, along with the scripts for the analyses (in R), are available in this GitHub repository: https://github.com/WakinyanB/OMEGA.

## Supplementary material

**Supplementary Figures** (PDF)

**Supplementary Information (SI Appendix)** (PDF)

## Data and code accessibility

Data that were used in this study, along with the scripts for the analyses (in R), are available in this GitHub repository: https://github.com/WakinyanB/OMEGA. Numerical simulations were performed using R [94] version 4.4.2 (2024-10-31) and ordinary differential equations were numerically solved with the function lsoda from the package deSolve [95].

## Acknowledgments

We thank Sébastien Lion for numerous inspiring discussions on the analysis of class-structured models. We also thank Erik Volz for sharing the data underlying the invasion of the SARS-CoV-2 Delta variant.

## Supplementary Figures

**List of supplementary figures** (pp. 2-11)

**Figure S1:**
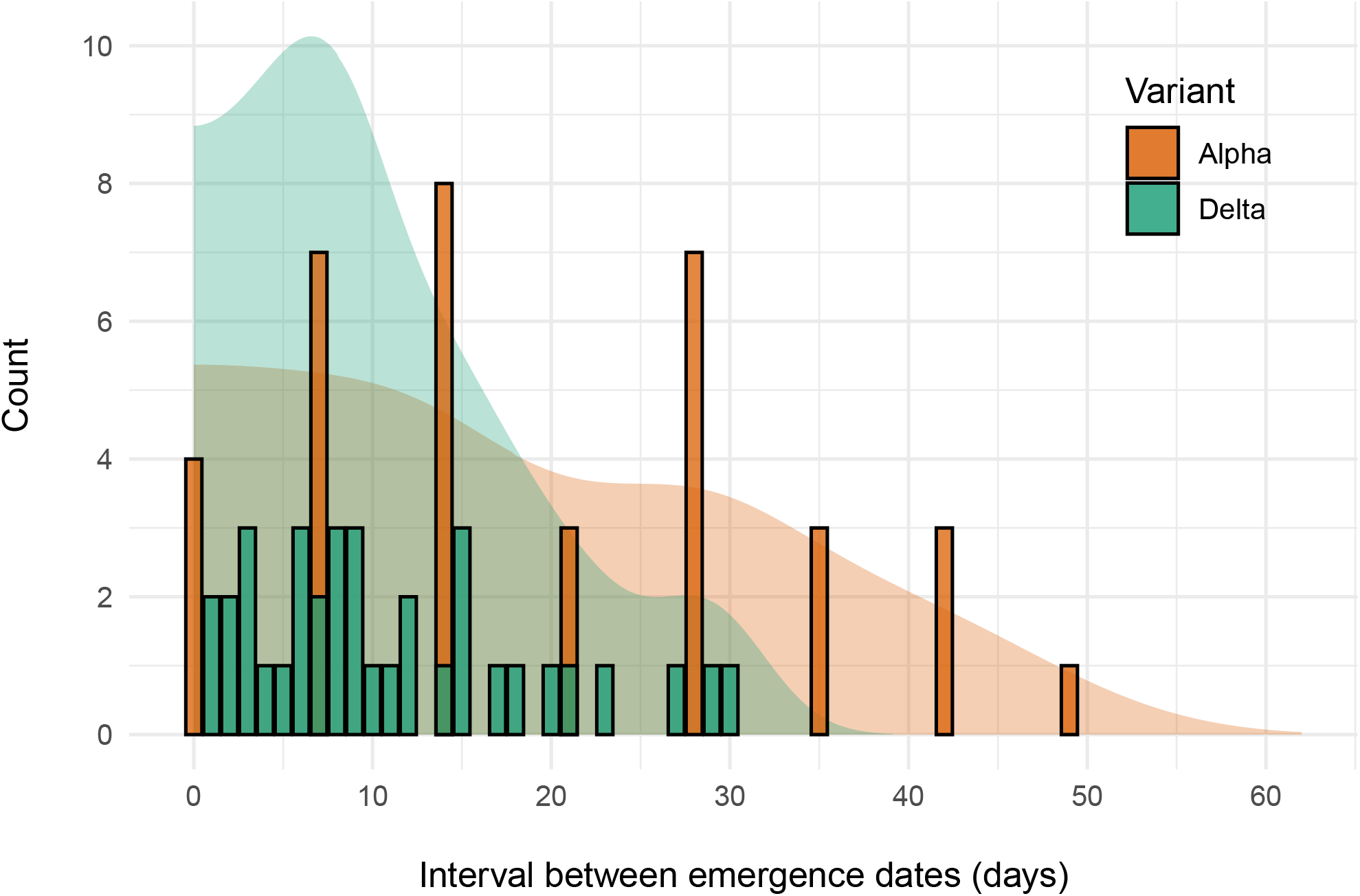
Delays between emergence of the SARS-CoV-2 Alpha and Delta variants across regions of England. We compute pairwise time differences between emergence dates from **Fig. 1** and plot the corresponding distributions (count and density) for both variants. In each case, the time of introduction of the new variant differs substantially among regions, but delays between successive introductions are longer on average for Alpha. The latter pattern might be induced by a weaker interregional connectivity (i.e., fewer migration events between regions).

**Figure S2:**
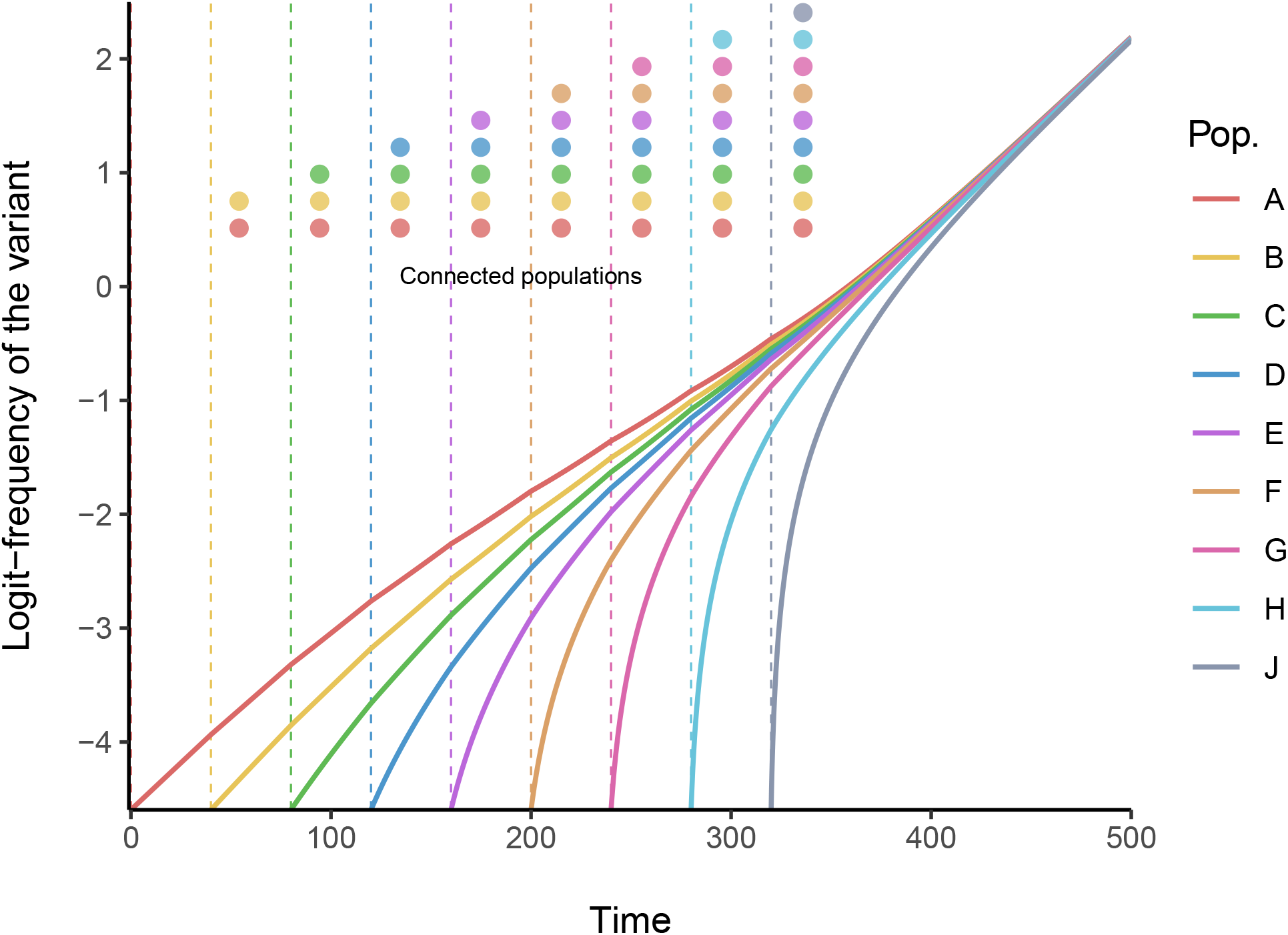
Evolutionary dynamics of an emerging variant in a nine-patch host metapopulation. We extend in **SI Appendix §S6.1** the model presented in the main text to any number of populations. Here, the host metapopulation is divided in nine populations (such as the nine regions of England). For the sake of simplicity, the variant has only a transmission advantage and the phenotypes of both strains do not vary across populations: *β*_*w*_ = 0.11, *β*_*m*_ = 0.1265 (+15% transmission advantage), *γ*_*w*_ = *γ*_*m*_ = 0.1 and *ω* = 0. We also assume same total density for each population (100) and same probability of migration (0.01). At *t* = 0, the wildtype is introduced at low density (10^−7^) in all populations, but the variant is only introduced in population *A* at a lower density (1% of infected hosts *I*^*A*^) – all populations are otherwise fully susceptible. All populations are first isolated from each other. Populations are then sequentially connected by migration, starting from population *A* and *B*. Every time a population is connected (as indicated by the vertical dashed lines), we introduce the variant such that it represents 1% of the infected hosts in that population.

**Figure S3:**
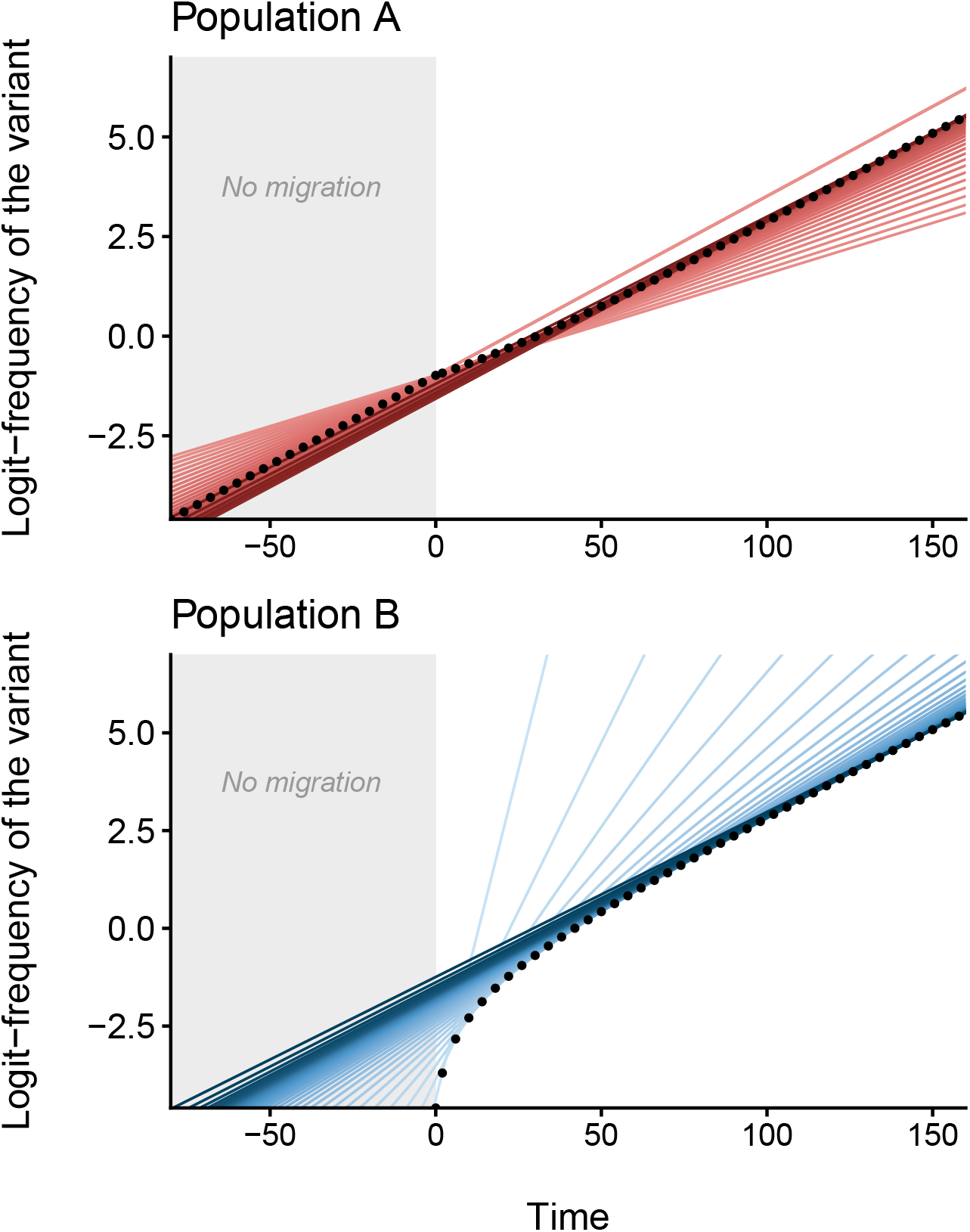
Migration can steepen the change of local frequency of an emerging variant. This figure is built upon the simulation presented in the main text in **Fig. 1**. The host metapopulation is divided in two populations, *A* (top) and *B* (bottom). The variant has a +30% transmission rate selective advantage over the wildtype. We simulate the model with identical total population densities (*N*^*A*^ = *N*^*B*^ = 100), and parameter values (same for both populations): *β*_*w*_ = 0.15, *β*_*m*_ = 0.195, *γ*_*w*_ = *γ*_*m*_ = 0.1 and *ω* = 0. The wildtype is initially introduced at very low density (10^−7^) in both populations and the variant is only introduced in population *A* such that it represents 1% of the infected hosts *I*^*A*^ – both populations are otherwise fully susceptible. Before *t* = 0, the two populations are isolated (*µ*^*A*^ = *µ*^*B*^ = 0, gray background). At *t* = 0, the variant is introduced in population *B* (1% of *I*^*B*^) and, from that time point onward, hosts from population *B* may visit population *A* (*µ*^*B*^ = 0.1). Tangents (red and blue lines) to each logit-frequency values (black points) indicate the corresponding current rate of change. Tangent slopes are approximated using equations (1)-(2) from the main text; color shades (from lighter to darker) represent the course of time.

**Figure S4:**
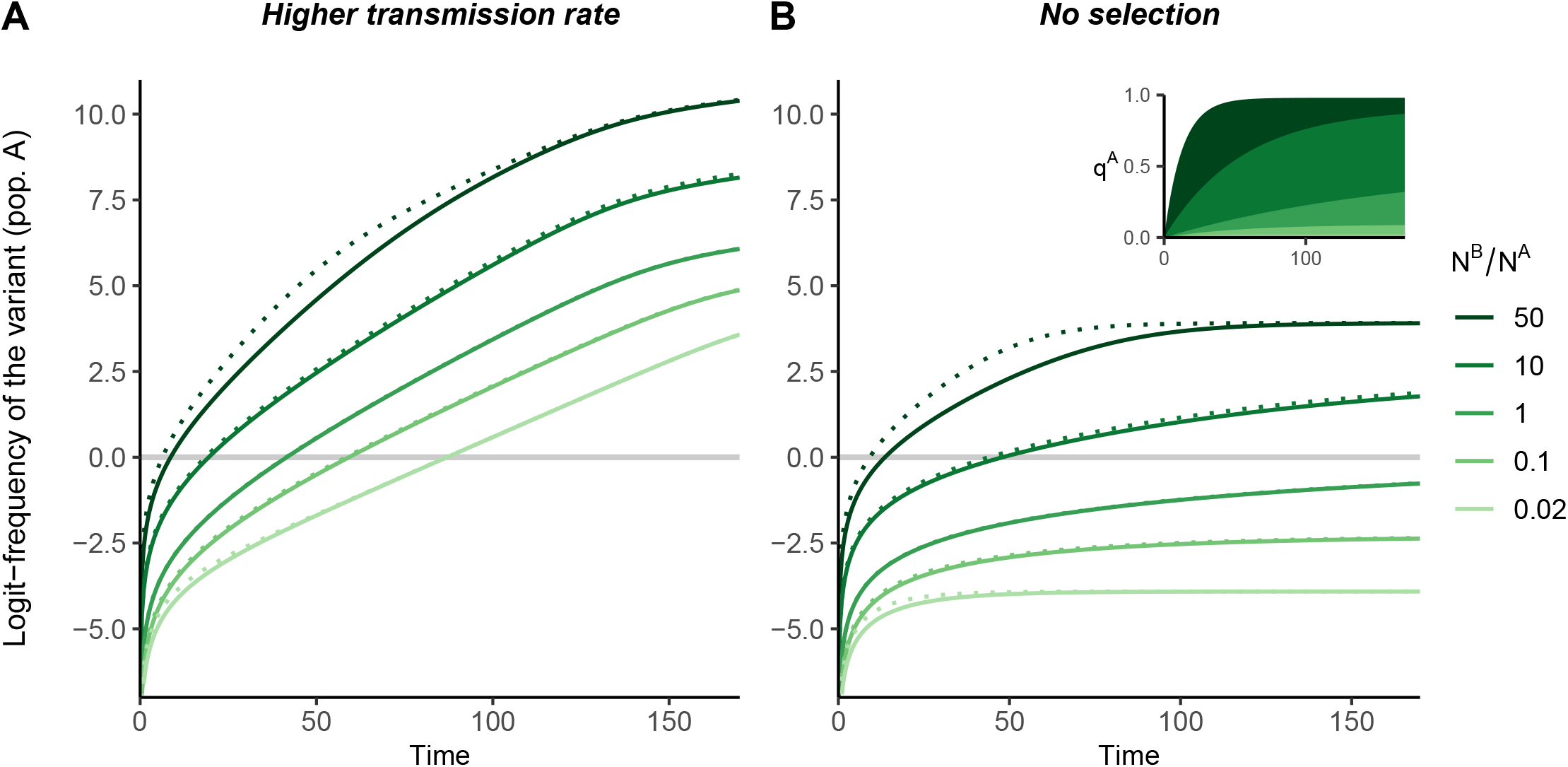
The effect of population size asymmetry on the dynamics of the variant frequency in the focal population. The host metapopulation is divided in two populations, *A* and *B*. Varying the total population density ratio *N*^*B*^*/N*^*A*^ (in all cases, *N*^*A*^ = 100), we simulate our full model (**SI Appendix §S2.2**, system (S5), solid lines) along with our first-order approximation under weak migration (dotted lines). Trajectories of the logit-frequency of a variant under: (A) positive selection (+30% transmission rate, *β*_*m*_ *> β*_*w*_ ⇒ Δ*β >* 0), (B) neutral evolution (*β*_*m*_ = *β*_*w*_ ⇒ Δ*β* = 0). In all simulations, we take (same for both populations): *β*_*w*_ = 0.15, *γ*_*w*_ = *γ*_*m*_ = 0.1, *µ*^*A*^ = *µ*^*B*^ = 10^−2^ and *ω* = 0. At *t* = 0, the wildtype is introduced at very low prevalence in population A(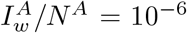 and 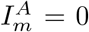) and the variant is introduced at the same prevalence in population B(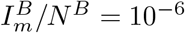 and 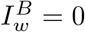) – both populations are otherwise fully susceptible.

**Figure S5:**
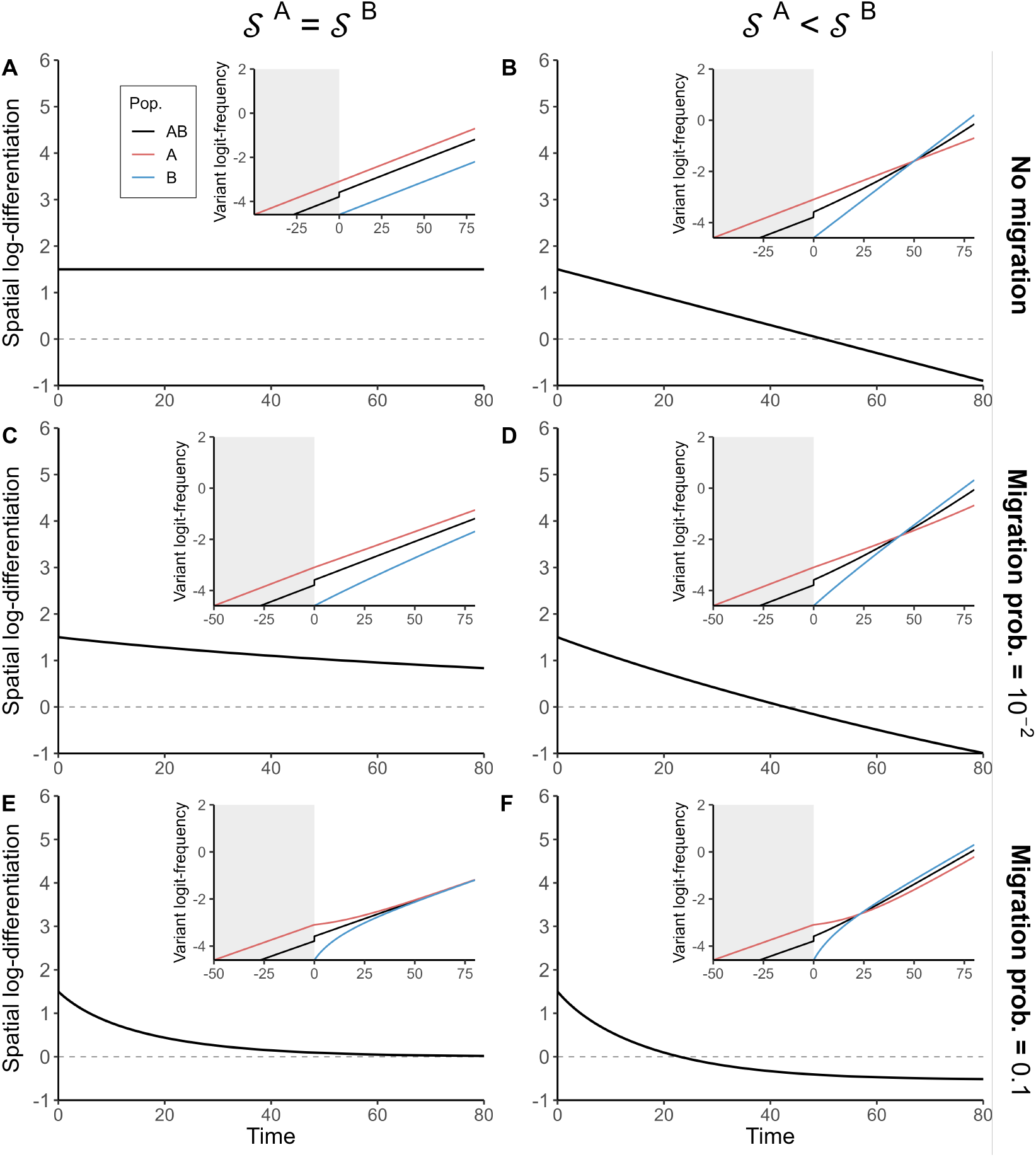
Short-term dynamics of spatial differentiation and the migration–selection balance. For the sake of simplicity, we assume that both strains share the same transmissibility. This allows us to set aside the dynamical effects in the selection coefficient. Fixed parameter values are: *ω* = 0 and ∀*i* ∈ {*A, B*}, *N*^*i*^ = 100, 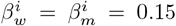and 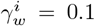 The recovery rate of the variant do not vary across populations (homogeneous selection, 𝒮^*A*^ = 𝒮^*B*^): 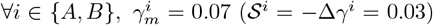. (B-F) The variant is more adapted in population *B* (heterogeneous selection, 𝒮^*A*^ < 𝒮^*B*^): 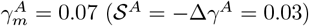 and 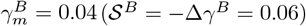. The wildtype is initially introduced in both populations at the same density (10^−4^) and the variant is only introduced in population *A* such that it represents 1% of infected hosts *I*^*A*^ – both populations are otherwise fully susceptible. Until *t* = 0, the two populations are isolated (*µ*^*A*^ = *µ*^*B*^ = 0) and the variant is only present in population *A* (gray background). At *t* = 0, the variant is introduced in population *B* at low density (1% of *I*^*B*^) and the two populations (A-B) stay isolated, or are coupled with (C-D) low or (E-F) higher levels of migration. Note that epidemiological dynamics exhibit only little variations over the simulation period.

**Figure S6:**
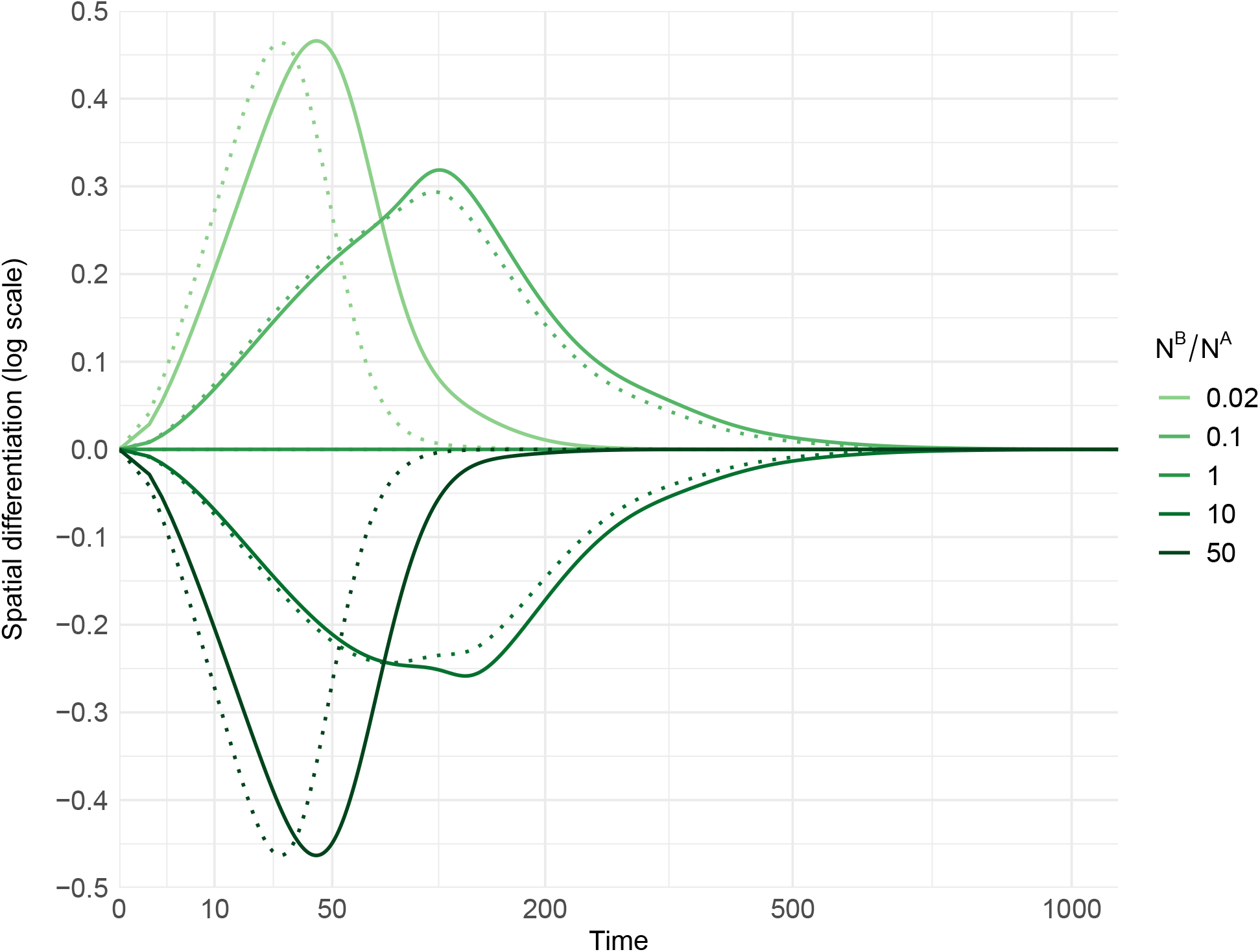
The transient disruption of spatial differentiation. The host metapopulation is divided in two populations, *A* and *B*. Varying the total population density ratio *N*^*B*^*/N*^*A*^ (in all cases, *N*^*A*^ = 100), we simulate our full model (**SI Appendix §S2.2**, system (S5), solid lines) with parameter values (same for both populations): *β*_*w*_ = 0.15, *β*_*m*_ = 0.195 (+30% transmission rate, Δ*β* = 0.045), *γ*_*w*_ = *γ*_*m*_ = 0.1 (Δ*γ* = 0), *ω* = 0.01 and *µ*^*A*^ = *µ*^*B*^ = 10^−2^. We also run these simulations using our first-order approximation under weak migration (dotted lines). At *t* = 0, the wildtype and the variant are introduced at the same low *density* in both populations 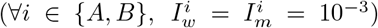 both populations are otherwise fully susceptible. The log-differentiation ln (𝒬) is then calculated as ln (𝒬) = logit (*q*^*A*^) − logit (*q*^*B*^). Although there is no spatial differentiation initially (ln (𝒬) = 0), the spatial differentiation is then transiently disrupted as soon as *N*^*A*^ ≠ *N*^*B*^ because the initial differential in the proportions of infected and susceptible hosts results in a differential in selection when Δ*β* ≠ 0.

**Figure S7:**
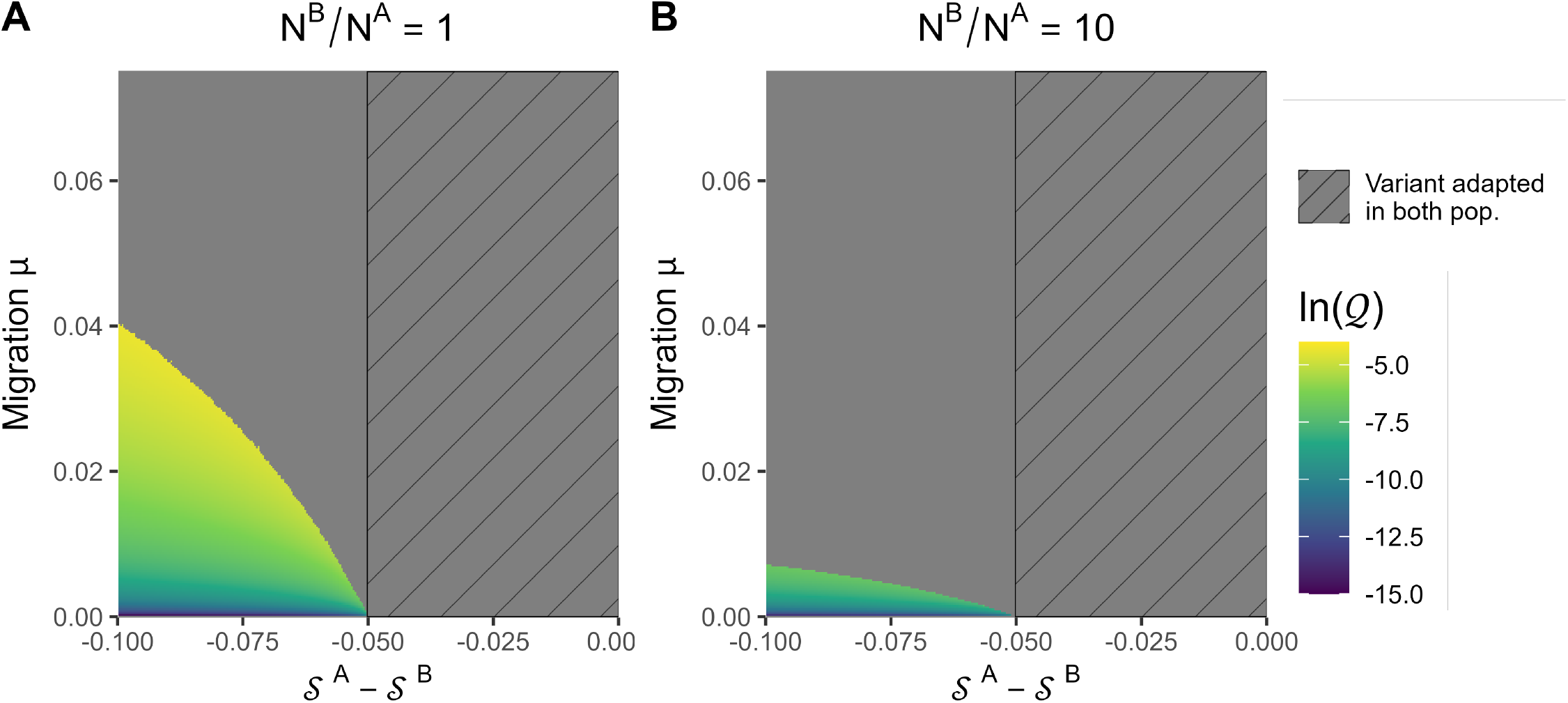
Long-term spatial differentiation and local adaptation. For the sake of simplicity, we assume that both strains share the same transmissibility and only differ in terms of recovery rates. This allows us to set aside the dynamical effects in the selection coefficients such that the difference 𝒮^*A*^ *− 𝒮*^*B*^ reduces to −Δ*γ*^*A*^ + Δ*γ*^*B*^. Parameter values shared by both populations are: *β*_*w*_ = *β*_*m*_ = 0.15, *γ*_*w*_ = 0.1, 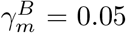 and *ω* = 0.01; the variant is thus selected for in population *B* (𝒮^*B*^ = *−* Δ*γ*^*B*^ *>* 0). We compute the long-term spatial differentiation by varying the parameter value of 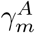 from 0.05 to 0.15 and of the migration probability *µ* = *µ*^*A*^ = *µ*^*B*^ from 0 to 0.075. In population *A*, depending on the value of 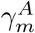, the variant is either selected for (hatching background, 𝒮^*A*^ = −Δ*γ*^*A*^ *>* 0) or against (𝒮 ^*A*^ = − Δ*γ*^*A*^ *<* 0). Simulations were performed with (A) *N*^*A*^ = *N*^*B*^ = 100 or (B) *N*^*A*^ = 100 and *N*^*B*^ = 1000. The gray color corresponds to cases where a strain has numerically reached fixation in a population.

**Figure S8:**
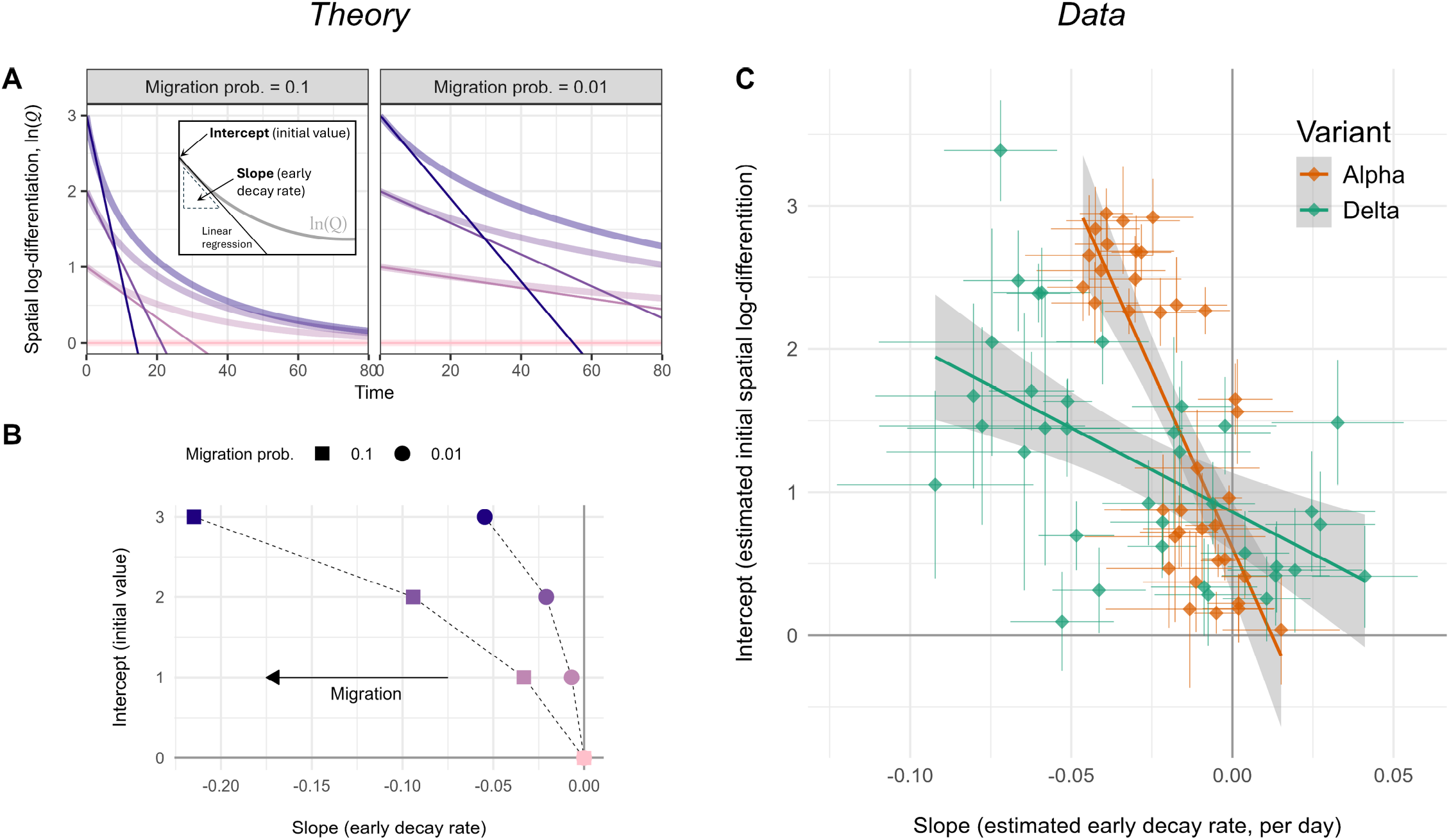
Approximations of the early dynamics of the spatial differentiation. (A-B) ‘Theory’ panel. According to equation (3), the dynamics of the spatial differentiation 𝒬 depends on the differential in selection between the two populations, the current level of differentiation and the intensity of migration. In the following we assume that selection is homogeneous across populations and we only focus on the last two. (A) Dynamics of the spatial log-differentiation ln (𝒬) = logit (*q*^*A*^) − logit (*q*^*B*^) starting from different initial conditions and with two level of migration (*µ*^*A*^ = *µ*^*B*^ = 0.01 or 0.1) – parameter values (same for all populations): *β*_*w*_ = *β*_*m*_ = 0.15, *γ*_*w*_ = 0.1 and *γ*_*m*_ = 0.05. Straight lines represent linear approximations of the early dynamics of ln (𝒬) and we reported the corresponding slopes and intercepts in panel B. (C) ‘Data’ panel. We estimated the early dynamics of the spatial log-differentiation (slope and intercept) of the Alpha and Delta variants using simple linear regressions. We used the data and the pairs of regions presented in **Fig. 4-C** and **D** (using daily data for Delta) for which elapsed time did not exceed 35 days. We oriented each pair so that the point estimate of the intercept was always positive. Interestingly, the patterns we obtain for SARS-CoV-2 are consistent with the hypothesis that migration was higher during the sweep of Delta because the drop in differentiation is steeper than for the Alpha variant (straight lines and gray envelopes show a linear regression on point estimates).

**Figure S9:**
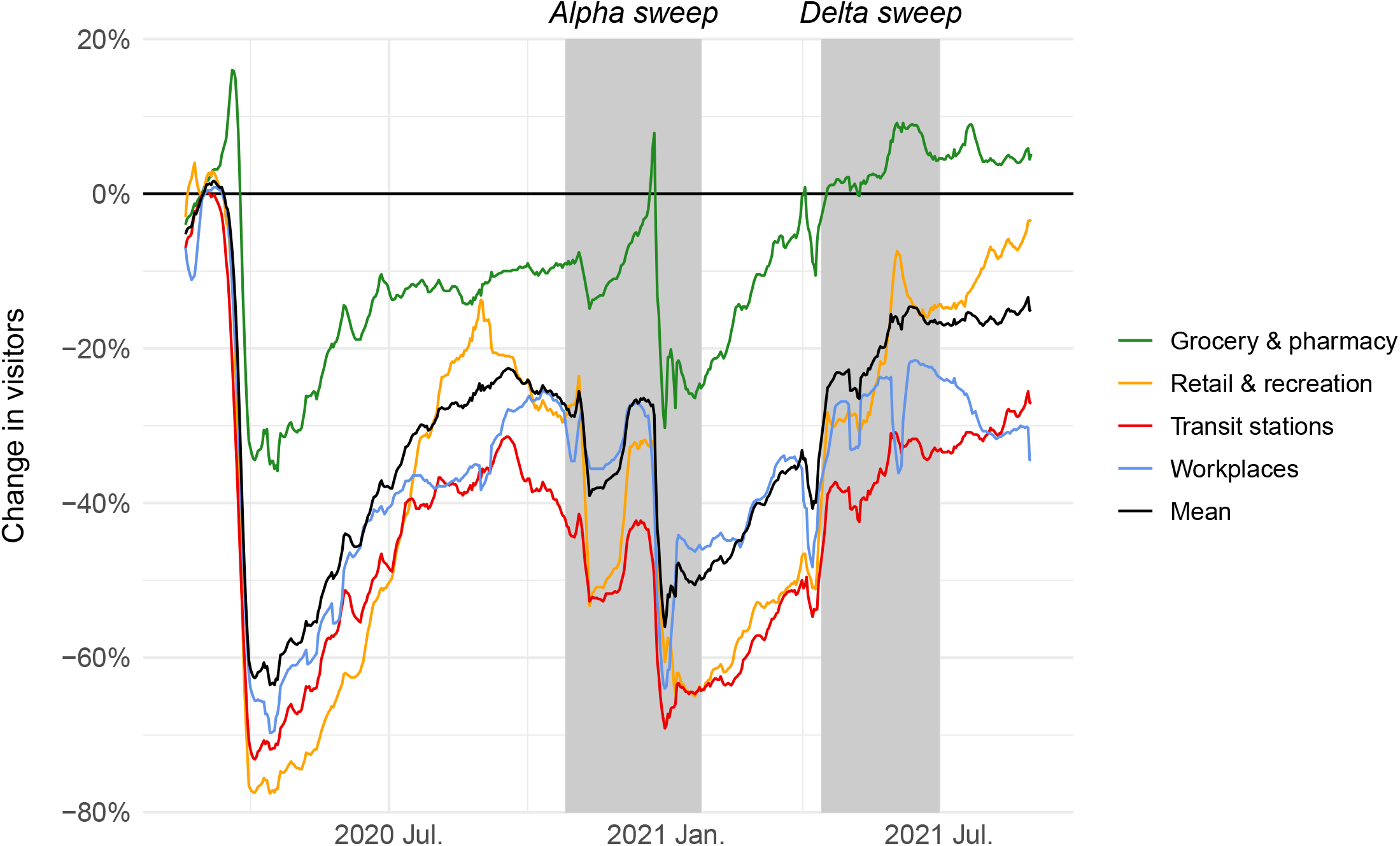
Change in visitors in the United Kingdom (Google mobility reports). Changes in the number of visitors compared to baseline days (median value from the 5-week period from Jan. 3 to Feb. 6, 2020). Mobility data were downloaded from the website *Our World in Data* (https://ourworldindata.org/covid-mobility-trends, last visited 2025-10-10) and we only focused on the following locations: grocery & pharmacy, retail & recreation, transit stations and workplaces (the mean across these four place categories is shown in black). The gray rectangles indicate the period of the sweep of the Alpha and Delta variants.

**Figure S10:**
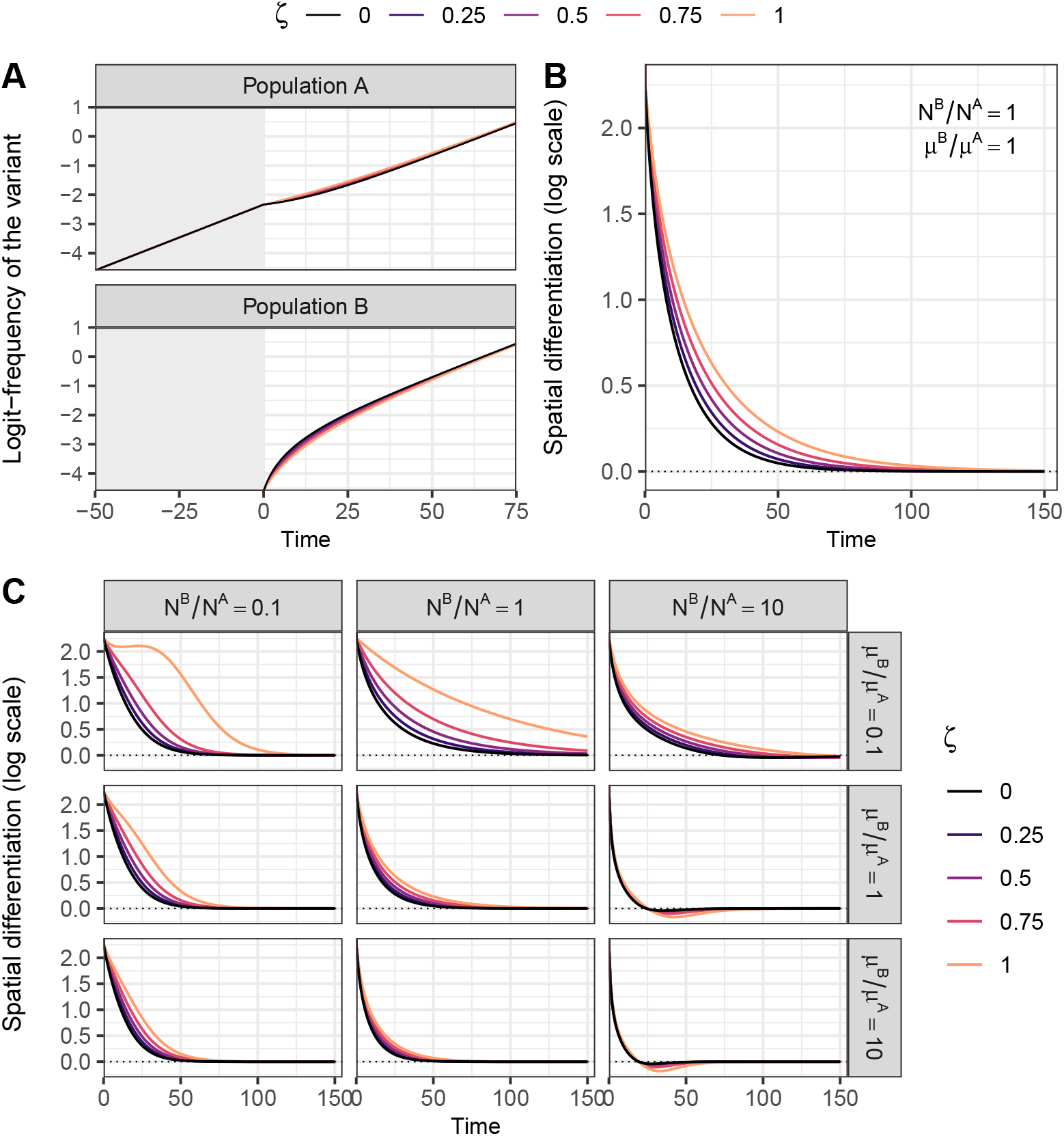
The effect of reduced mobility in infected hosts on the evolutionary dynamics. The host metapopulation is divided in two populations, *A* and *B*. The variant has a +30% transmission rate selective advantage over the wildtype. We simulate the model with identical total population densities (*N*^*A*^ = *N*^*B*^ = 100), and parameter values (same for both populations): *β*_*w*_ = 0.15, *β*_*m*_ = 0.195, *γ*_*w*_ = *γ*_*m*_ = 0.1 and *ω* = 0. The wildtype is initially introduced at very low density (10^−7^) in both populations and the variant is only introduced in population *A* such that it represents 1% of the infected hosts *I*^*A*^ – both populations are otherwise fully susceptible. Before *t* = 0, the two populations are isolated (*µ*^*A*^ = *µ*^*B*^ = 0, gray background). At *t* = 0, the variant is introduced in population *B* (1% of *I*^*B*^) and, from that time point onward, hosts may visit the other population (*µ*^*A*^ = *µ*^*B*^ = 0.1). In **SI Appendix §S6.2**, we relax the assumption that infected hosts (*I*) commute with the same probability that non-infected hosts (*S* and *R*), such that ζ represents the reduction in host mobility due to the disease, ranging from 0 (no impact, same as in the main text) to 1 (no mobility of infected hosts). Varying the value of ζ, we plot the temporal dynamics of (A) the variant logit-frequency and of (B) the spatial log-differentiation ln (𝒬) = logit (*q*^*A*^) − logit (*q*^*B*^). In (C), we focus on the dynamics of ln (𝒬), varying also population size and migration asymmetries between the two populations.

## Supplementary Information (SI Appendix)

### S1 A two-patch host metapopulation coupling epidemiology and evolution

We model the dynamics of a directly and horizontally transmitted infectious disease. To do so, we rely on a classical susceptible-infected-recovered-susceptible (SIRS) models. Hosts are either susceptible (*S*), infected/infectious (*I*) or recovered (*R*). For each compartment “*X*”, we denote *X* its density at the current time *t* (we drop the time dependence in our notations for compactness and readability). For the sake of simplicity, we neglect host birth and death processes and assume that the total host population density *N* = *S* + *I* + *R* remains constant over time. Let *β* be the *per capita* transmission rate – which captures both the host contact rate and the probability of transmission per contact with an infected host –, and *γ*, the *per capita* recovery rate (infection clearance). We assume that there is no pathogen virulence (i.e., disease-induced mortality).

We focus on a scenario where two strains (genotypes) of the pathogen are spreading in the host population and we neglect the appearance of new mutations. Without loss of generality, let us call them the wildtype (*w*) and the mutant strain (*m*), or variant. Throughout, we use the subscripts *w* and *m* to refer to each strain. In addition, we assume that a host can only be infected once, i.e., we assume no super-infection (including co-infections by both strains) and that, after recovery, infection provides perfect immunity to both strains (full cross-immunity) – recovered individuals are however assumed to return to the susceptible compartment at a *per capita* rate *ω* (waning immunity). The total density of infected hosts *I* is thus *I* = *I*_*w*_ + *I*_*m*_. Crucially, the phenotype of the variant may differ from the wildtype in terms of transmission rate (*β*_*m*_ = *β*_*w*_ + Δ*β*) and/or recovery rate (*γ*_*m*_ = *γ*_*w*_ + Δ*γ*).

We also consider a two-patch metapopulation, that is, two host populations, labeled *A* and *B*, coupled by migration (we extend this model to *n* ≥ 2 populations in the section **§S6.1**). Throughout, we use superscripts to distinguish the two populations. We denote *N*^*A*^ and *N*^*B*^, the (constant) total densities of populations *A* and *B*, respectively, such that *N*^*A*^ + *N*^*B*^ = *N*. Now define *µ*^*i*^, the probability for a host from the focal population *i* ∈ {*A, B*} to visit the non-focal population. For the sake of simplicity, we assume that *µ*^*i*^ is independent of both the host’s epidemiological status (*S, I* or *R*), and the pathogen’s strain (*w, m*). We thus neglect the potential influence the pathogen may have on the mobility of its host – e.g., if hosts tend to limit their movements when they present symptoms or are aware of being infected – (we relax this assumption in the last section **§S6.2**), and we assume that migration is not itself under selection. Visits are considered to be only (instantaneous) to-and-fro host movements, such as commuting [1], neglecting the impact of longer or permanent stays. Strictly speaking, we thus model the migration of the disease, rather than that of hosts [2]. Finally, we assume that, due to some spatial heterogeneities between the two populations (e.g., non-pharmaceutical interventions, behaviors, treatments) phenotypic traits *β* and *γ* can vary between populations, so that the phenotypic traits of the variant in population *i* is given by

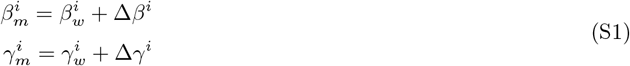

For example, if the efficacy of control measures implemented to reduce transmission in the focal population *i* is denoted *c*^*i*^, then we would have 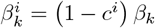, with *β*_*k*_, the baseline transmissibility of strain *k*.

### S2 Epidemiology

#### S2.1 At the metapopulation level

Epidemiological dynamics at the metapopulation level are given by the following system of ordinary differential equations (ODEs)

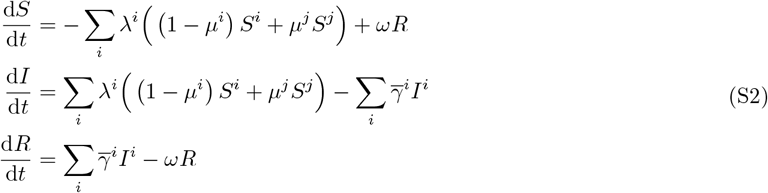

with 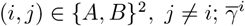 is the mean recovery rate in population *i* (averaged across pathogen genotypes)

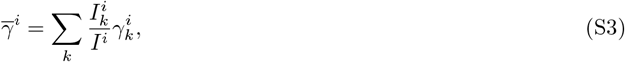

with *k* ∈ {*w, m*}, and *λ*^*i*^ represents the force of infection (i.e., the *per capita* infection rate) experienced in population *i*. Assuming transmission to be frequency-dependent [3], we have

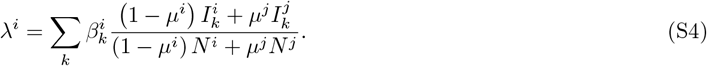

The total force of infection is therefore the sum of the contributions of four types of intra or inter-community interactions [4]. The intra-community contributions correspond to transmissions between a susceptible and an infected host who belong to the same population, which can occur either (i) in the local population or (ii) in the visited population (visitor-to-visitor infection). In contrast, the inter-community contributions correspond to transmissions between a susceptible and an infected host who do not belong to the same population, which can occur (i) when a susceptible visitor gets infected by a native or (ii) when a susceptible native gets infected by a visitor.

#### S2.2 At the level of the focal population

Epidemiological dynamics for the focal population *i* ∈ {*A, B*} (accordingly, the non-focal population is denoted by _*i*_ *j*) are given by the following system of ODEs

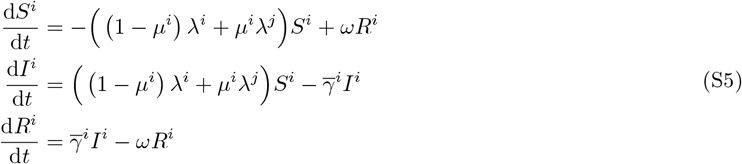

with *k* ∈ {*w, m*} and with *λ*^*i*^ and *λ*^*j*^, the forces of infection experienced in the focal and non-focal population, respectively. Focusing only on the epidemiological dynamics of hosts from the focal population *i* who are infected by the variant, we have

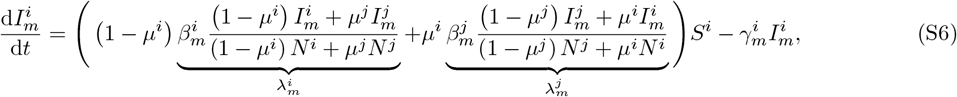

with 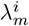 and 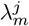, the force of infection of the variant in population *i* and *j*, respectively.

### S3 Evolution

Alongside epidemiological dynamics, we use a population genetics approach and track the evolution of the system over time (strain frequency and spatial differentiation).

#### S3.1 Frequencies of the variant

First, at the level of the focal population *i* ∈ {*A, B*}, define

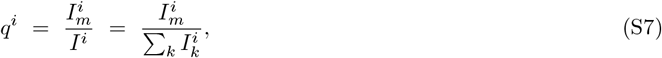

with *k* ∈ {*w, m*}, the frequency of the variant in the focal population *i* at the current time *t*. The temporal dynamics of the variant in the focal population *i* is given by the derivative of *q*^*i*^ with respect to time, that is,

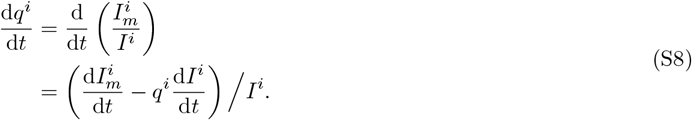

Second, at the metapopulation level, let *q* be the overall frequency of the variant

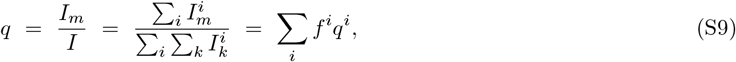

where *f*^*i*^ = *I*^*i*^*/I* is the frequency of infected hosts that belong to population *i* ∈ {*A, B*}. Likewise, the temporal dynamics of *q* is given by

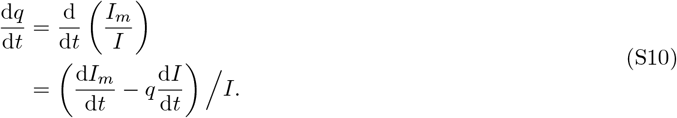

#### S3.2 Spatial differentiation

We now introduce the following measure of the genetic differentiation between population *A* and *B* (hereafter referred to as spatial differentiation)

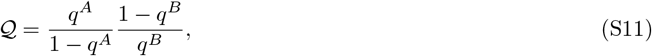

such that

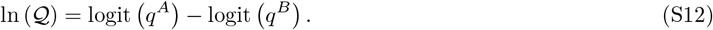

Note that ln (𝒬) is only defined when (*q*^*A*^, *q*^*B*^ ) ∈]0, 1[^2^ (i.e., neither strain has reached fixation). When the variant is more frequent in population *A* than in population *B*, 𝒬 *>* 1 and ln (𝒬) *>* 0. When the variant is found in equal frequency in both populations, 𝒬 = 1 and ln (𝒬) = 0 (no differentiation). We used similar quantities in previous work to track the differentiation across pathogen life stages [5, 6], rather than across space, but the principle remains exactly the same. Though the spatial differentiation would be more classically expressed on the natural scale as *q*^*A*^ − *q*^*B*^, the dynamics of this measure is more difficult to interpret because it depends on the genetic variances which are also dynamical variables (see **§S4.2.3**). We thus focus more conveniently on 𝒬. Using equation (S12), the temporal dynamics of the log-differentiation ln (𝒬) is given by

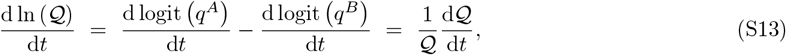

#### S3.3 Relation between local and global variables

Using either the pair of variables *q*^*A*^ and *q*^*B*^ (“local” variables) or the pair *q* and 𝒬 (“global” variables) are sufficient to fully describe the evolutionary dynamics of the system. The definitions of *q* and 𝒬 are based on *q*^*A*^ and *q*^*B*^ and, the other way around, *q*^*A*^ and *q*^*B*^ can be expressed as functions of *q* and 𝒬

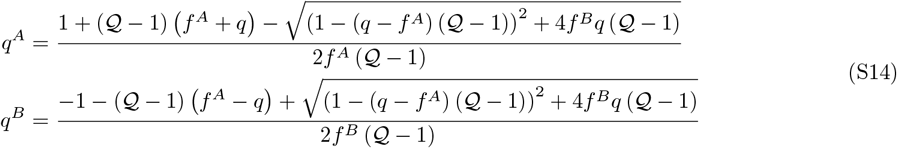

The case 𝒬 = 1 is a removable discontinuity for which *q*^*A*^ = *q*^*B*^ = *q*. When the differentiation 𝒬 is not too far from 1 (small differentiation), a Taylor expansion about the case 𝒬 = 1 yields the approximations

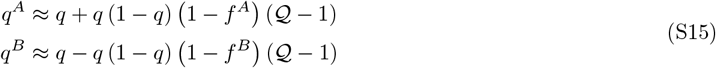

This also implies that

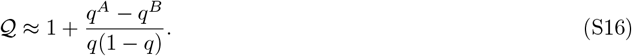

### S4 Weak migration approximation

We now assume that the amount of host mobility is small, i.e., hosts interact much more locally rather than with other populations. In the following, we treat *µ*^*i*^ as *ϵ*_ℳ_*M*^*i*^ and *µ*^*j*^ as *ϵ*_ℳ_*M*^*j*^ to emphasize that the migration probabilities of the hosts are small, of order *ϵ*_ℳ_ ≪ 1. We referred to this approximation in the main text as the weak migration assumption.

#### S4.1 Epidemiological dynamics

A Taylor expansion of the force of infection *λ*^*i*^ (S4) about *ϵ*_ℳ_ = 0 yields

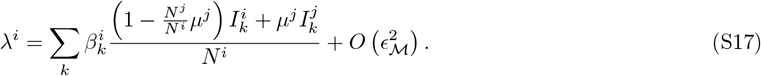

Note that the term *O* (*ϵ*_ℳ_) depends on *µ*^*j*^ but not on *µ*^*i*^ anymore. Likewise, a Taylor expansion of the ODE system (S5) about *ϵ*_ℳ_ = 0 yields the following epidemiological dynamics for the infected hosts from population *i*

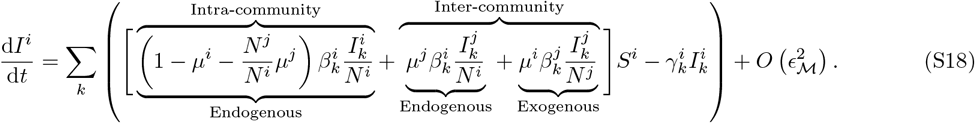

Here, “endogenous” (resp. “exogenous”) refers to infections that occur within (resp. outside) the focal population, regardless of the population to which the infector belongs. Note that all the visitor-to-visitor contribution to the force of infection (intra-community but exogenous) is now included in the term 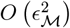.

#### S4.2 Evolutionary dynamics

##### S4.2.1 Dynamics of the variant frequency at the population level

Using equations (S8) and (S18), the dynamics of the variant frequency *q*^*i*^ is given after some rearrangements by

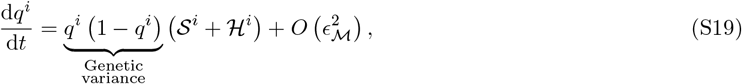

with

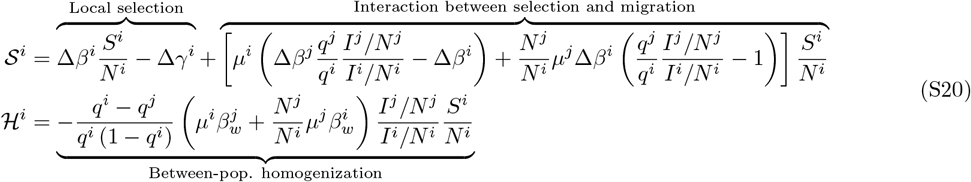

Both 𝒮^*i*^ and ℋ^*i*^ are first-order approximations in *ϵ*_ℳ_. The component 𝒮^*i*^ collects all the terms that depend on the phenotypic differences between the two strains and refer to the effects of local selection on the focal population as well as the interaction between selection and migration between populations. The component ℋ^*i*^ is a homogenization force that affects the local dynamics of the variant as soon as the variant frequency varies between the two connected populations (i.e., *q*^*i*^≠ *q*^*j*^). We obtain a similar partitioning following an approach analogous to [7] (**see details in §S5.2.2**).

The selection coefficient of the variant is typically quantified by estimating the regression slope of its frequency changes on the logit scale [8, 9]. We thus focus instead on the dynamics of the logit-frequency of the variant which is given by

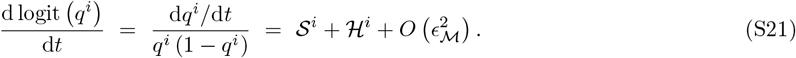

##### S4.2.2 Dynamics of the variant frequency at the metapopulation level

Likewise, the temporal dynamics of the variant logit-frequency at the metapopulation level (S10) is given under the weak migration assumption by

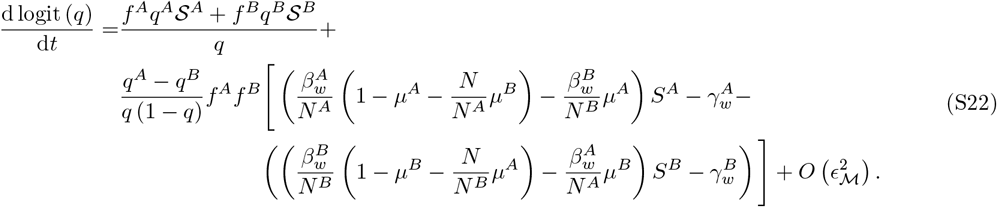

If selection is homogeneous across space (i.e., 𝒮^*A*^ = 𝒮^*B*^ = 𝒮), then (*f* ^*A*^*q*^*A*^𝒮^*A*^ + *f* ^*B*^*q*^*B*^𝒮^*B*^*) /q* is simply 𝒮. Besides, in the absence of host mobility (i.e., *µ*^*A*^ = *µ*^*B*^ = 0), the previous equation reduces to

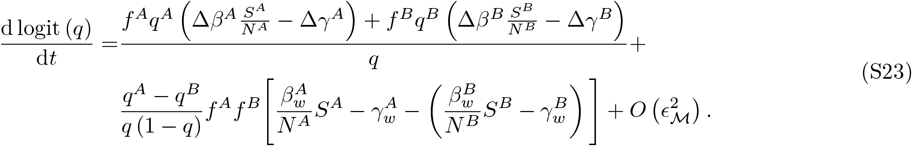

Following an approach similar to [7], we show in **§S5.2.1** how to tease out the effects of selection on the change in frequency.

##### S4.2.3 Dynamics of the spatial differentiation

Using (S19), the dynamics of the difference *q*^*A*^ − *q*^*B*^ is

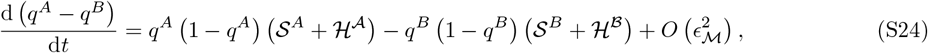

which depends on the genetic variance *q*^*i*^ (1 − *q*^*i*^) in each population *i*. Alternatively, we can use the measure of spatial differentiation 𝒬 (as defined in (S11)), whose dynamics on the log scale (S13) can be expressed in a simpler form using equation (S21)

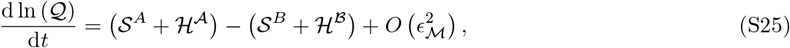

which gives after some rearrangements of ℋ^*A*^ − ℋ^*B*^

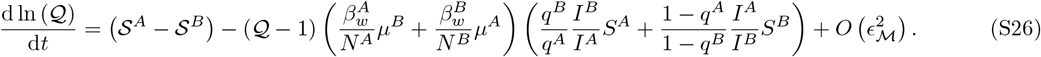

The differentiation 𝒬 may rapidly reach a quasi-equilibrium value 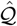 when migration is large relative to selection. This quasi-equilibrium can be obtained by setting the right-hand side of (S26) to 0 and solving for 𝒬, which yields

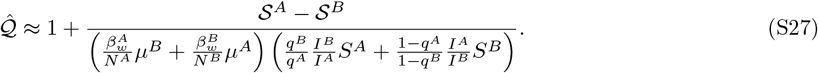

This quasi-equilibrium can be viewed as a classical balance between the effects of selection (numerator) and the effects of migration (denominator), and can only be different from 1 when selection is heterogeneous across populations (𝒮^*A*^≠ 𝒮^*B*^).

If the variant is adapted in one population but maladapted in the other, the variant may persist at an intermediate frequency without going to fixation (i.e., polymorphism maintenance) when migration is not too strong (**Fig. S7**). In this case, (S27) is the long-term equilibrium value of 𝒬.

### S5 Relation with Priklopil & Lehmann (2024), *American Naturalist*

In this section, we follow the approach used in [7] to disentangle the effects of “*natural selection*” from the effects of “*class transmission*” on the change in frequency of pathogen variants. We keep the term “*class transmission*”, defined in [7] as “*nonheritable fitness differences*”, “*which also [affect] allele-frequency change even in the absence of selection*”. While the partitioning in [7] is based on a discrete model, we use here a continuous version instead in order to match the framework used in the main text. Similarly to [7], we first start with a general model to describe the dynamics of a class-structure population. Second, we derive the dynamics of the global frequency of a variant, for which we partition selection from class transmission, and apply the obtained decomposition on the model presented in the main text. Last, we do the same but at the level of a single class (i.e., local variant frequency). In the latter, we show that this approach yields results identical to those presented in the main text.

#### S5.1 A general model to describe the dynamics of a class-structure population

We consider here a host population divided between *n* ≥ 2 classes, labeled from 1 to *n* – e.g., in the main text, these classes would represent different populations within a host metapopulation. Let 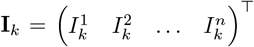 be the vector collecting the densities of hosts infected by strain *k* ∈ {*w, m*}. We model the temporal dynamics of **I**_*k*_ by the following ODE

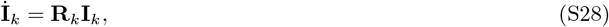

where **R**_*k*_ is the matrix of transmission/transition rates

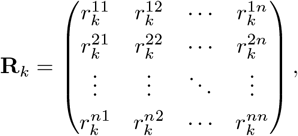

so that 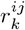 is the growth rate of *i*-class individual per *j*-class individual

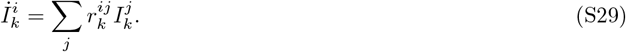

This equation is the continuous counterpart of equation (1) in [7]. We then also define 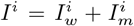, the total density of infected hosts in population *i*, 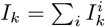, the total density of hosts infected by strain *k*, and 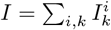, the total density of infected hosts in the metapopulation. Therefore

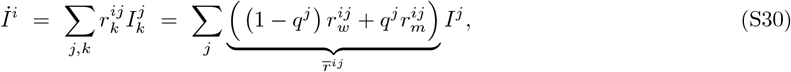

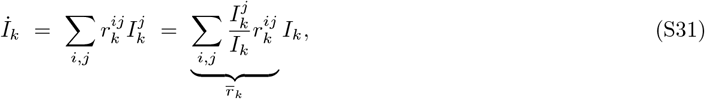

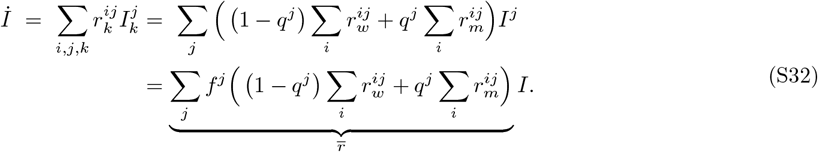

In the previous equations, 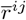 represent the pathogen growth rate (or fitness) in population *i* due to *j*-class individual, 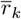, the fitness of strain *k* and 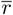, the overall fitness of the pathogen; *f*^*j*^ = *I*^*j*^*/I* is the proportion of infected hosts in population *j*.

#### S5.2 Partitioning selection and class transmission

##### S5.2.1 Global frequency of the variant

The dynamics of the (global) frequency of the variant *q* = *I*_*m*_*/I* is given by the following ODE

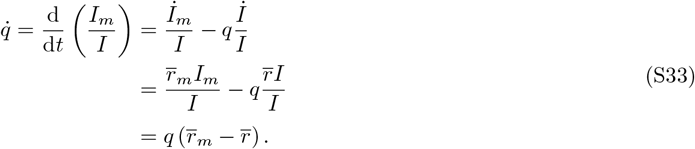

This equation is equivalent to the equation (2) in [7]. After some rearrangements, equation (S33) can also be written as

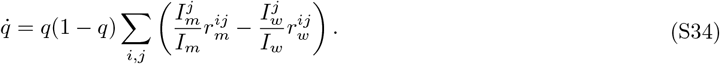

Note that, in the absence of selection, we have 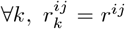, so that 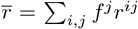 and 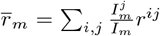 which, applied to equation (S33), yields a neutral process where

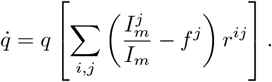

Similarly to equation (3) in [7], this equation shows that, even in the absence of fitness difference (and thus of selection), class transmissions can still affect the dynamics of the variant frequency *q*; these non-selective changes only arise when there is an asymmetric distribution of the pathogen strains across classes, and stop as soon as 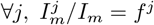.

To disentangle the effects of selection from those of class transmission, we use the approach presented in [7] and decompose the dynamics of hosts infected by strain *k* as follows

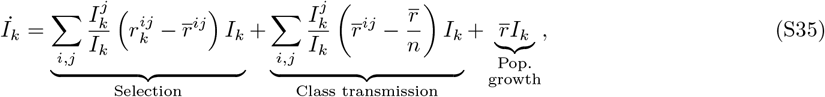

where the mean 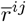 represents a reference measure of fitness. Equation (S35) is equivalent to equation (S6) in [7]. Plugging (S35) into (S33) yields

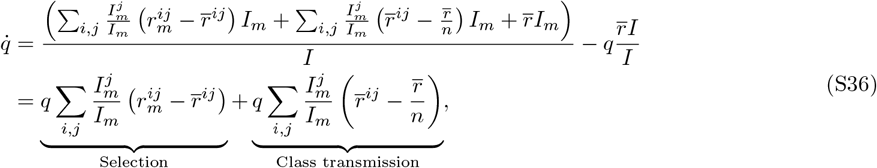

which is similar to equation (4) in [7]. Likewise, note that the last term in (S35) (population growth) cancels out in (S36) and thus does not contribute to the dynamics of *q*. Summing over all strains of the pathogen, we can verify that natural selection is indeed a conservative evolutionary force (i.e., if one strain increases in frequency, the other must decrease)

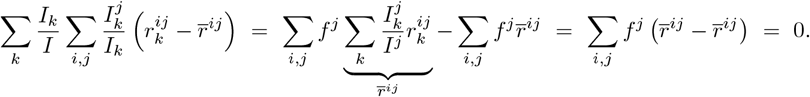

We then apply the partitioning (S36) to the model presented in the main text, for which we have ∀(*i, j*) ∈ {*A, B*}^2^, *j*≠ *i*

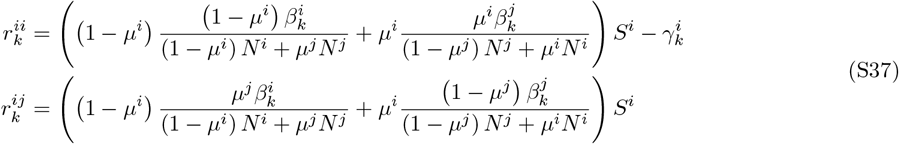

Under the assumption of weak migration, a Taylor expansion yields

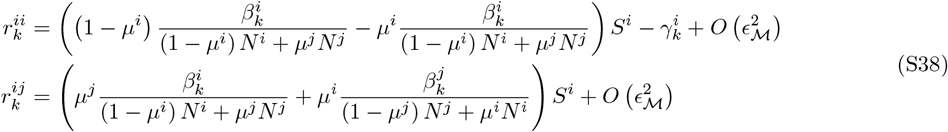

Using 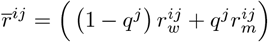 and plugging (S38) into (S36) yields the selection component

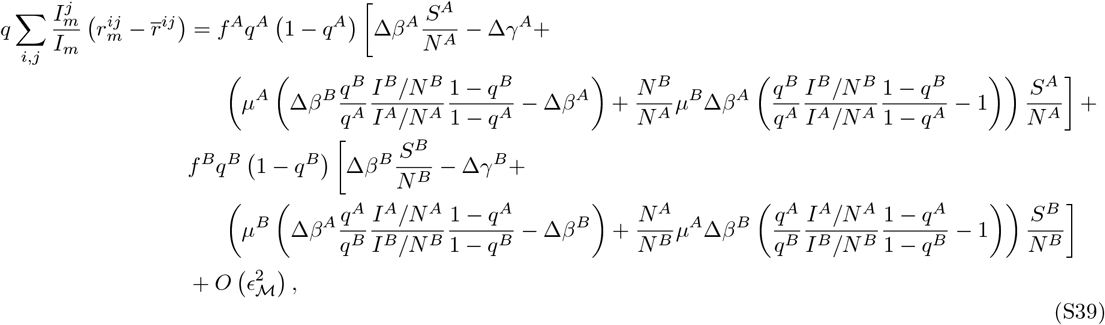

and the class-transmission component

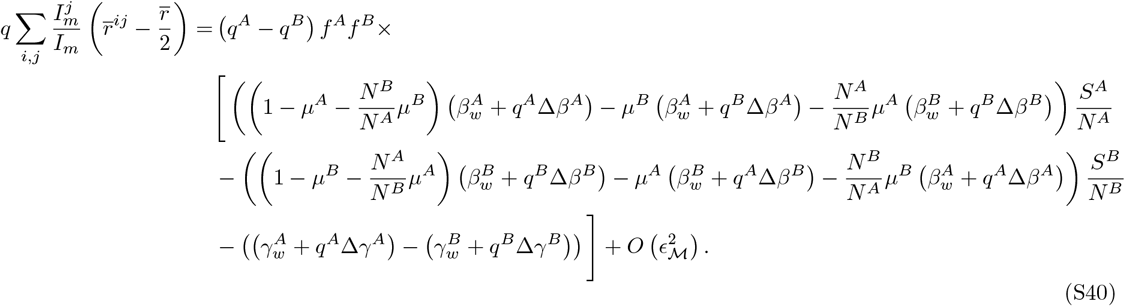

Note that the terms

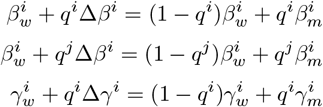

correspond to mean trait values. For the transmission rate *β*, these mean trait values can be averaged across the genotype distribution of the pathogen in either population (*i* or *j*) depending on where the infection occurs.

##### S5.2.2 Local frequency of the variant

We now focus on the local variant frequency, i.e., at the level of a single class *i*. The dynamics of the local frequency of the variant 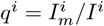 is given by the following ODE

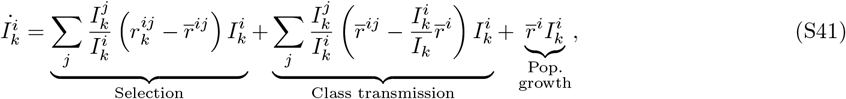

with

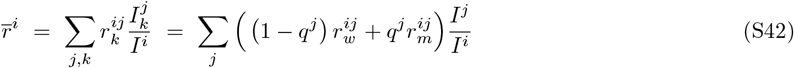

and where, again, the mean 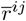 represents a reference measure of fitness. Thus

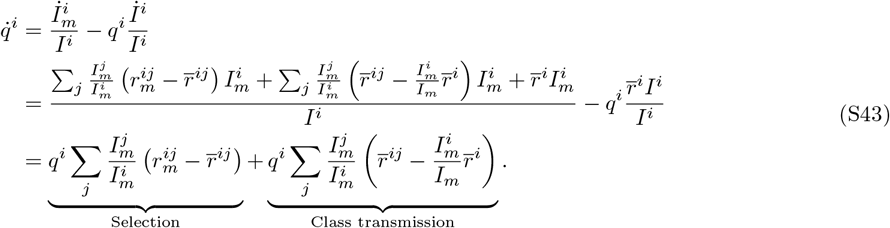

Again, the last term in (S41) (population growth) cancels out in (S44) and we can also verify that the effects of natural selection, summed over all strains of the pathogen, is null

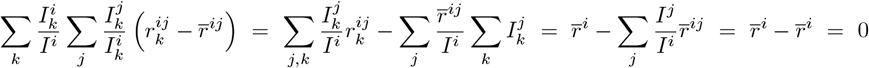

After some rearrangements, we have

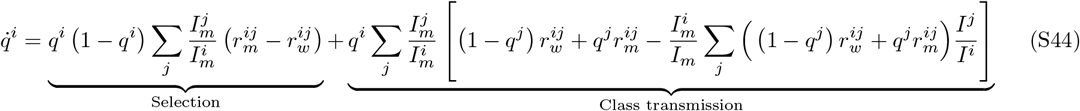

We then apply the partitioning (S44) to the model presented in the main text. Using the growth rates given in equations (S38), we have

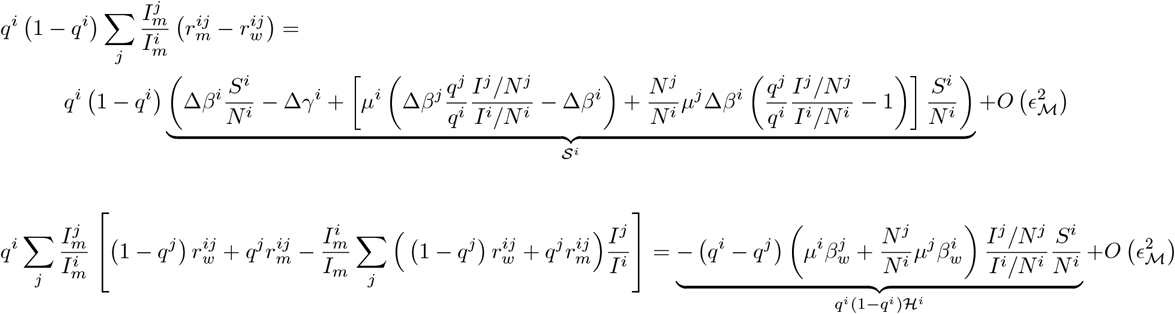

We thus recover the results derived in the main text (the only difference being that our formulation scales each component by the genetic variance *q*^*i*^ (1 − *q*^*i*^))

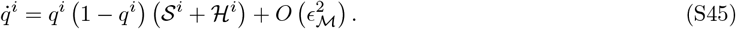

### S6 Model extensions

#### S6.1 Extension to an arbitrary number of populations

More generally, we can extend the previous model to *n* ≥ 2 populations.

##### S6.1.1 Epidemiological dynamics

At the level of the focal population *i*, the dynamics of the density of infected hosts in model (S5) becomes

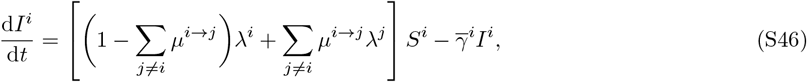

with *k* ∈ {*w, m*} and where *j* refers to the index of the *j*th population and *µ*^*i*→*j*^ (resp. *µ*^*j*→*i*^), to the probability of migration from the focal population *i* (resp. from the non-focal population *j*) to population *j* (resp. *i*). As before, *λ*^*i*^ and *λ*^*j*^ are the forces of infection experienced in population *i* and *j*, respectively, such that

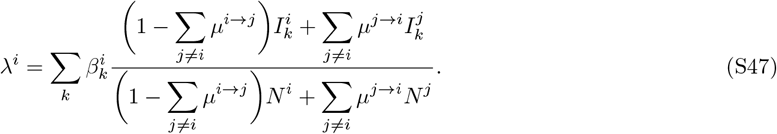

Under the weak migration assumption, a Taylor expansion of the previous equation about *ϵ*_ℳ_ = 0 (see details in **§S4**) yields

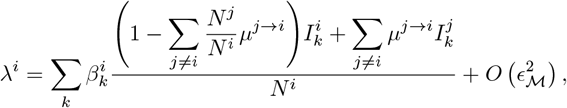

and, similarly, the dynamics of the density of infected hosts (S46) is now given by

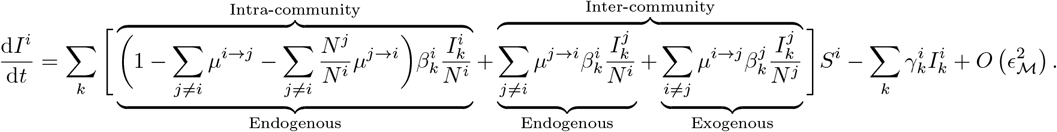

##### S6.1.2 Evolutionary dynamics

At the level of the focal population *i*, the dynamics of the variant logit-frequency 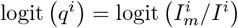 can still be expressed as (S21), but now with

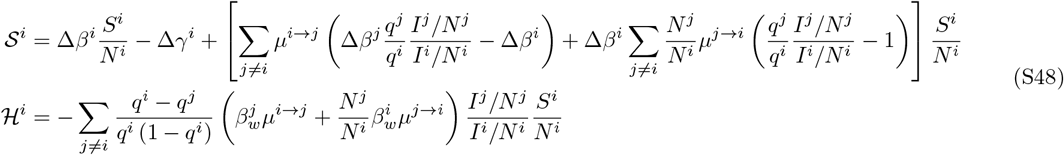

We illustrate the dynamics of the logit-frequency of an emerging variant in a nine-patch host metapopulation (such as the nine regions of England) in **Fig. S3**.

We can also define the genetic differentiation between any pair of populations, say *i* and *l*≠ *i*, as

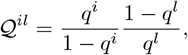

such that ln 𝒬^*il*^ = logit (*q*^*i*^ ) logit (*q*^*l*^ ). Again, under the assumption of weak migration, the dynamics of the spatial log-differentiation ln (𝒬^*il*^) is given by

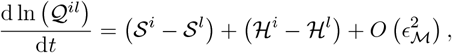

which, after some rearrangements, yields

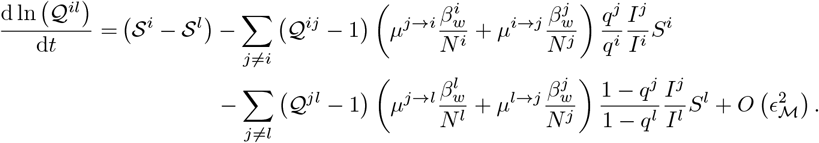

This expression can be difficult to use in practice as the dynamics of the spatial differentiation between two populations now depends not only on itself, but on the whole spatial differentiation structure in which populations *i* and *l* are embedded, including all the corresponding pairwise differentiation values. The weight of each corresponding pair of populations can either speed up or slow down the overall dynamics of the pairwise spatial differentiation.

#### S6.2 Extension to reduced mobility in infected hosts

In this section, we relax the assumption that infected hosts (*I*) commute with the same probability that non-infected hosts (*S* and *R*). To do so, we assume that infected individuals from the focal population *i* can commute to the non-focal population with probability (1 − ζ)*µ*^*i*^, where ζ ∈ [0, 1] represents the reduction in host mobility due to the disease. For the sake of simplicity, ζ is assumed to be the same for both populations and for both strains. Note that the case ζ = 0 corresponds thus to the scenario presented in the main text, in which the infection does not affect host mobility.

##### S6.2.1 Epidemiological dynamics

In these conditions, the ODE systems (S2) and (S5) are unchanged but the force of infection in the focal population *i* is now given by

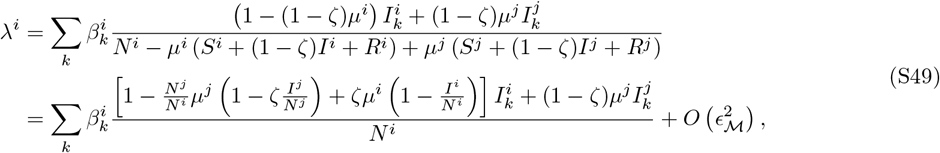

where the second line is a Taylor expansion of the first line under the assumption of the weak migration (see details in **§S4**). Plugging equation (S49) into the ODE system (S5), the dynamics of the density of infected hosts thus becomes

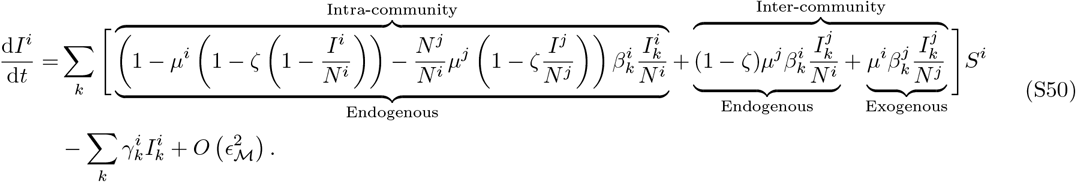

Note that, if ζ = 1 – i.e., infection prevents infected hosts from commuting between populations –, the endogenous inter-community term disappears because inter-community transmissions would solely occur through the commuting of susceptible hosts (infection in the non-focal population and pathogen importation in the focal population).

##### S6.2.2 Evolutionary dynamics

At the level of the focal population *i*, the dynamics of the variant logit-frequency can still be expressed as (S21), but now with

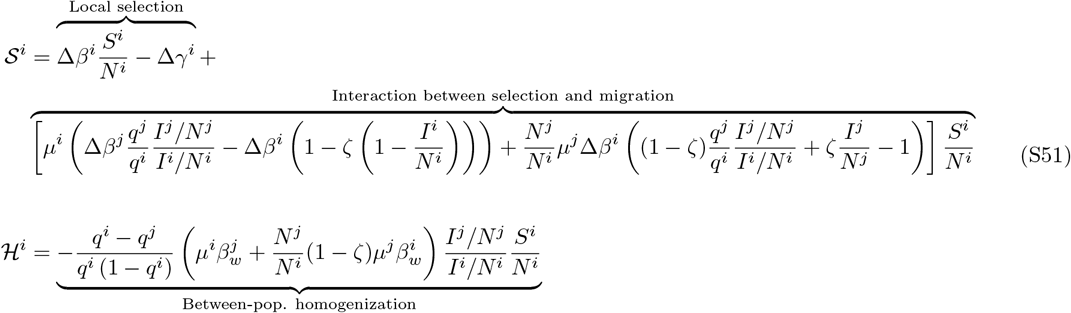

When ζ = 0 (no reduced mobility in infected hosts), we recover equations (S20), as presented in the main text. At the other end of the spectrum, when ζ = 1, equation (S51) reduces to

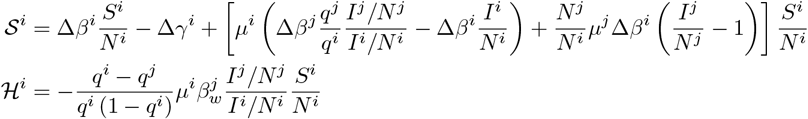

Again, the homogenization component ℋ^*i*^ now only depends on *µ*^*i*^ because inter-community transmissions can only be achieved through the mobility of susceptible hosts. In **Fig. S9-A**, we plot some examples for the dynamics of the variant logit-frequency by varying the value of ζ, from 0 to 1. Simulated trajectories are extremely similar, in particular because the proportion of infected hosts remains really small compared to the proportion of susceptibles.

Besides, the dynamics of the spatial log-differentiation ln (𝒬) = logit(*q*^*A*^ )− logit (*q*^*B*^) is now given by

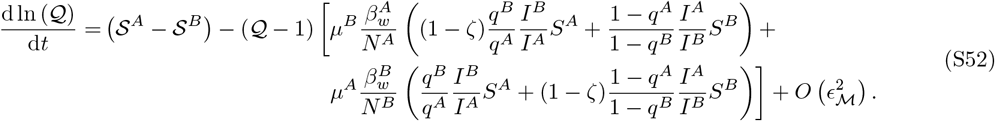

Again, when ζ = 0, we recover equations (S26). We show in **Fig. S9-B** some simulated dynamics for the spatial log-differentiation by varying the value of ζ, from 0 to 1. As expected, lower amounts of migration, due to reduced mobility rates in infected hosts (ζ → 1), slow down the homogenization process between the two populations; note however that, again, this effect is small because the proportion of infected hosts is small.

#### S6.3 Extension to partial cross-immunity

In this section, we extend the model (S5)-(S6) to account for partial cross-immunity. In the previous model, we assume that immunity provided by one strain also provides full protection against the other (full cross-immunity). By relaxing this assumption, we now need to take into account possible reinfections of recovered individuals by another strain. Because increasing the number of infected classes makes theoretical derivations much more complicated and difficult to compare with our previous model, we assume for simplicity that only the variant can partially escape the immunity provided by the wildtype. Define 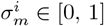 as the ability of the variant to infect hosts from population *i* who acquired immunity to the wildtype (denoted by 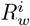), where 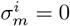 indicates full cross-immunity (as assumed in the main text) and 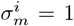 indicates no-cross immunity. Note that, since we assume 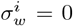, the phenotypic difference 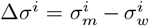 reduces to 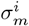 in this very special case. The dynamics of 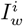 thus remains unchanged but that of 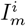 becomes

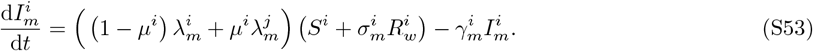

Then, the selection component 𝒮^*i*^ of the dynamics of the variant logit-frequency logit (*q*^*i*^ ) is given by

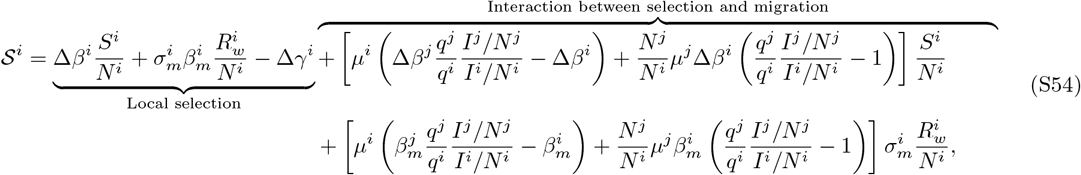

while the homogenization component ℋ^*i*^ remains the same as in (S20) (and, therefore, the dynamics of the spatial differentiation can still be written as (S26)).

## References

1. Fisher, R. A. The genetical theory of natural selection: a complete variorum edition (Oxford University Press, 1999).

2. Chevin, L.-M. On measuring selection in experimental evolution. Biology letters 7, 210–213. doi:10.1098/rsbl.2010.0580 (2011).

3. Boyle, L., Hletko, S., Huang, J., Lee, J., Pallod, G., Tung, H.-R. & Durrett, R. Selective sweeps in SARS-CoV-2 variant competition. Proceedings of the National Academy of Sciences 119, e2213879119. doi:10.1073/pnas.2213879119 (2022).

4. Volz, E. Fitness, growth and transmissibility of SARS-CoV-2 genetic variants. Nature Reviews Genetics 24, 724–734. doi:10.1038/s41576-023-00610-z (2023).

5. Donker, T., Papathanassopoulos, A., Ghosh, H., Kociurzynski, R., Felder, M., Grundmann, H. & Reuter, S. Estimation of SARS-CoV-2 fitness gains from genomic surveillance data without prior lineage classification. Proceedings of the National Academy of Sciences 121, e2314262121. doi:10.1073/pnas.2314262121 (2024).

6. Lythgoe, K. A., Golubchik, T., Hall, M., House, T., Cahuantzi, R., MacIntyre-Cockett, G., Fryer, H., Thomson, L., Nurtay, A., Ghafani, M., et al. Lineage replacement and evolution captured by 3 years of the United Kingdom Coronavirus (COVID-19) Infection Survey. Proceedings of the Royal Society B 290, 20231284. doi:10.1098/rspb.2023.1284 (2023).

7. Grubaugh, N. D., Hanage, W. P. & Rasmussen, A. L. Making sense of mutation: what D614G means for the COVID-19 pandemic remains unclear. Cell 182, 794–795. doi:10.1016/j.cell.2020.06.040 (2020).

8. Volz, E., Hill, V., McCrone, J. T., Price, A., Jorgensen, D., O’Toole, Á., Southgate, J., Johnson, R., Jackson, B., Nascimento, F. F., et al. Evaluating the effects of SARS-CoV-2 spike mutation D614G on transmissibility and pathogenicity. Cell 184, 64–75. doi:10.1016/j.cell.2020.11.020 (2021).

9. Yu, Q., Ascensao, J. A., Okada, T., COVID-19 Genomics UK (COG-UK) Consortium, Boyd, O., Volz, E. & Hallatschek, O. Lineage frequency time series reveal elevated levels of genetic drift in SARS-CoV-2 transmission in England. PLOS Pathogens 20, e1012090. doi:10.1371/journal.ppat.1012090 (2024).

10. Day, T., Gandon, S., Lion, S. & Otto, S. P. On the evolutionary epidemiology of SARS-CoV-2. Current Biology 30, R849–R857. doi:10.1016/j.cub.2020.06.031 (2020).

11. Otto, S. P., Day, T., Arino, J., Colijn, C., Dushoff, J., Li, M., Mechai, S., Van Domselaar, G., Wu, J., Earn, D. J., et al. The origins and potential future of SARS-CoV-2 variants of concern in the evolving COVID-19 pandemic. Current Biology 31, R918–R929. doi:10.1016/j.cub.2021.06.049 (2021).

12. Benhamou, W., Lion, S., Choquet, R. & Gandon, S. Phenotypic evolution of SARS-CoV-2: a statistical inference approach. Evolution 77, 2213–2223. doi:10.1093/evolut/qpad133 (2023).

13. Walter, A. & Lion, S. Epidemiological and evolutionary consequences of periodicity in treatment coverage. Proceedings of the Royal Society B 288, 20203007. doi:10.1098/rspb.2020.3007 (2021).

14. Day, T., Kennedy, D. A., Read, A. F. & Gandon, S. Pathogen evolution during vaccination campaigns. PLOS biology 20, e3001804. doi:10.1371/journal.pbio.3001804 (2022).

15. Walter, A., Gandon, S. & Lion, S. Effect of unequal vaccination coverage and migration on long-term pathogen evolution in a metapopulation. Journal of Evolutionary Biology 37, 189–200. doi:10.1093/jeb/voad016 (2024).

16. Kermack, W. O. & McKendrick, A. G. A contribution to the mathematical theory of epidemics. Proceedings of the royal society of london. Series A, Containing papers of a mathematical and physical character 115, 700–721. doi:10.1098/rspa.1927.0118 (1927).

17. Anderson, R. M. & May, R. M. Infectious diseases of humans: dynamics and control (Oxford University Press, 1991).

18. Webb, S. D., Keeling, M. J. & Boots, M. Host–parasite interactions between the local and the mean-field: How and when does spatial population structure matter? Journal of Theoretical Biology 249, 140–152. doi:10.1016/j.jtbi.2007.06.013 (2007).

19. Lipshtat, A., Alimi, R. & Ben-Horin, Y. Commuting in metapopulation epidemic modeling. Scientific Reports 11, 15198. doi:10.1038/s41598-021-94672-w (2021).

20. Ball, F., Mollison, D. & Scalia-Tomba, G. Epidemics with two levels of mixing. The Annals of Applied Probability, 46–89. doi:10.1214/aoap/1034625252 (1997).

21. Ferguson, N. M., Galvani, A. P. & Bush, R. M. Ecological and immunological determinants of influenza evolution. Nature 422, 428–433. doi:10.1038/nature01509 (2003).

22. Grenfell, B. & Harwood, J. (Meta)population dynamics of infectious diseases. Trends in ecology & evolution 12, 395–399. doi:10.1016/S0169-5347(97)01174-9 (1997).

23. Arino, J. in Modeling and dynamics of infectious diseases 64–122 (World Scientific, 2009). doi:10.1142/9789814261265_0003.

24. Lajmanovich, A. & Yorke, J. A. A deterministic model for gonorrhea in a nonhomogeneous population. Mathematical Biosciences 28, 221–236. doi:10.1016/0025-5564(76)90125-5 (1976).

25. Hethcote, H. W. An immunization model for a heterogeneous population. Theoretical population biology 14, 338–349. doi:10.1016/0040-5809(78)90011-4 (1978).

26. Post, W., DeAngelis, D. & Travis, C. Endemic disease in environments with spatially heterogeneous host populations. Mathematical Biosciences 63, 289–302. doi:10.1016/0025-5564(82)90044-X (1983).

27. May, R. M. & Anderson, R. M. Spatial heterogeneity and the design of immunization programs. Mathematical Biosciences 72, 83–111. doi:10.1016/0025-5564(84)90063-4 (1984).

28. Bolker, B. & Grenfell, B. T. Space, persistence and dynamics of measles epidemics. Philosophical Transactions of the Royal Society of London. Series B: Biological Sciences 348, 309–320. doi:10.1098/rstb.1995.0070 (1995).

29. Watts, D. J., Muhamad, R., Medina, D. C. & Dodds, P. S. Multiscale, resurgent epidemics in a hierarchical metapopulation model. Proceedings of the National Academy of Sciences 102, 11157–11162. doi:10.1073/pnas.0501226102 (2005).

30. Colizza, V. & Vespignani, A. Epidemic modeling in metapopulation systems with heterogeneous coupling pattern: Theory and simulations. Journal of theoretical biology 251, 450–467. doi:10.1016/j.jtbi.2007.11.028 (2008).

31. Brockmann, D. & Helbing, D. The hidden geometry of complex, network-driven contagion phenomena. science 342, 1337–1342. doi:10.1126/science.1245200 (2013).

32. Yuksel, M. K., Remien, C. H., Karki, B., Bull, J. J. & Krone, S. M. Vector dynamics influence spatially imperfect genetic interventions against disease. Evolution, Medicine, and Public Health 9, 1–10. doi:10.1093/emph/eoaa035 (2021).

33. Roques, L., Bonnefon, O., Baudrot, V., Soubeyrand, S. & Berestycki, H. A parsimonious approach for spatial transmission and heterogeneity in the COVID-19 propagation. Royal Society Open Science 7, 201382. doi:10.1098/rsos.201382 (2020).

34. Le Treut, G., Huber, G., Kamb, M., Kawagoe, K., McGeever, A., Miller, J., Pnini, R., Veytsman, B. & Yllanes, D. A high-resolution flux-matrix model describes the spread of diseases in a spatial network and the effect of mitigation strategies. Scientific Reports 12, 15946. doi:10.1038/s41598-022-19931-w (2022).

35. Haldane, J. B. S. The theory of a cline. Journal of genetics 48, 277–284. doi:10.1007/BF02986626 (1948).

36. Nagylaki, T. Conditions for the existence of clines. Genetics 80, 595–615. doi:10.1093/genetics/80.3.595 (1975).

37. Slatkin, M. Gene flow and selection in a cline. Genetics 75, 733–756. doi:10.1093/genetics/75.4.733 (1973).

38. Débarre, F., Bonhoeffer, S. & Regoes, R. R. The effect of population structure on the emergence of drug resistance during influenza pandemics. Journal of the Royal Society Interface 4, 893–906. doi:10.1098/rsif.2007.1126 (2007).

39. Freire, T. F. A., Hu, Z., Wood, K. B. & Gjini, E. Modeling spatial evolution of multidrug resistance under drug environmental gradients. PLOS Computational Biology 20, e1012098. doi:10.1371/journal.pcbi.1012098 (2024).

40. Boots, M. & Sasaki, A. ‘Small worlds’ and the evolution of virulence: infection occurs locally and at a distance. Proceedings of the Royal Society of London. Series B: Biological Sciences 266, 1933–1938. doi:10.1098/rspb.1999.0869 (1999).

41. Boots, M., Hudson, P. J. & Sasaki, A. Large shifts in pathogen virulence relate to host population structure. Science 303, 842–844. doi:10.1126/science.1088542 (2004).

42. Lion, S. & Boots, M. Are parasites “prudent” in space? Ecology Letters 13, 1245–1255. doi:10.1111/j.1461-0248.2010.01516.x (2010).

43. Osnas, E. E., Hurtado, P. J. & Dobson, A. P. Evolution of pathogen virulence across space during an epidemic. The American Naturalist 185, 332–342. doi:10.1086/679734 (2015).

44. Berngruber, T. W., Lion, S. & Gandon, S. Spatial structure, transmission modes and the evolution of viral exploitation strategies. PLOS Pathogens 11, e1004810. doi:10.1371/journal.ppat.1004810 (2015).

45. Griette, Q., Raoul, G. & Gandon, S. Virulence evolution at the front line of spreading epidemics. Evolution 69, 2810–2819. doi:10.1111/evo.12781 (2015).

46. Zurita-Gutiérrez, Y. H. & Lion, S. Spatial structure, host heterogeneity and parasite virulence: implications for vaccine-driven evolution. Ecology letters 18, 779–789. doi:10.1111/ele.12455 (2015).

47. Lion, S. & Gandon, S. Evolution of spatially structured host–parasite interactions. Journal of Evolutionary Biology 28, 10–28. doi:10.1111/jeb.12551 (2015).

48. Lion, S. & Gandon, S. Spatial evolutionary epidemiology of spreading epidemics. Proceedings of the Royal Society B 283, 20161170. doi:10.1098/rspb.2016.1170 (2016).

49. Griette, Q., Alfaro, M., Raoul, G. & Gandon, S. Evolution and spread of multiadapted pathogens in a spatially heterogeneous environment. Evolution Letters 8, 427–436. doi:10.1093/evlett/qrad073 (2024).

50. Lion, S., Sasaki, A. & Boots, M. Extending eco-evolutionary theory with oligomorphic dynamics. Ecology Letters 26, S22–S46. doi:10.1111/ele.14183 (2023).

51. Priklopil, T. & Lehmann, L. On the interpretation of the operation of natural selection in class-structured populations. The American Naturalist 203, 292–304. doi:10.1086/727970 (2024).

52. Day, T. & Gandon, S. Insights from Price’s equation into evolutionary epidemiology. Disease evolution: models, concepts, and data analyses 71, 23–44. doi:10.1090/dimacs/071/02 (2006).

53. Day, T. & Gandon, S. Applying population-genetic models in theoretical evolutionary epidemiology. Ecology Letters 10, 876–888. doi:10.1111/j.1461-0248.2007.01091.x (2007).

54. Gandon, S. & Day, T. The evolutionary epidemiology of vaccination. Journal of the Royal Society Interface 4, 803–817. doi:10.1098/rsif.2006.0207 (2007).

55. Lion, S. & Gandon, S. Evolution of class-structured populations in periodic environments. Evolution 76, 1674–1688. doi:10.1111/evo.14522 (2022).

56. Benhamou, W., Blanquart, F., Choisy, M., Berngruber, T. W., Choquet, R. & Gandon, S. Evolution of virulence in emerging epidemics: from theory to experimental evolution and back. Virus Evolution 10, veae069. doi:10.1093/ve/veae069 (2024).

57. Blanquart, F., Gandon, S. & Nuismer, S. The effects of migration and drift on local adaptation to a heterogeneous environment. Journal of evolutionary biology 25, 1351–1363. doi:10.1111/j.1420-9101.2012.02524.x (2012).

58. Blanquart, F., Kaltz, O., Nuismer, S. L. & Gandon, S. A practical guide to measuring local adaptation. Ecology letters 16, 1195–1205. doi:10.1111/ele.12150 (2013).

59. Mirrahimi, S. & Gandon, S. Evolution of specialization in heterogeneous environments: equilibrium between selection, mutation and migration. Genetics 214, 479–491. doi:10.1534/genetics.119.302868 (2020).

60. Slatkin, M. Gene flow and the geographic structure of natural populations. Science 236, 787–792. doi:10.1126/science.3576198 (1987).

61. Simon, A. & Coop, G. The contribution of gene flow, selection, and genetic drift to five thousand years of human allele frequency change. Proceedings of the National Academy of Sciences 121, e2312377121. doi:10.1073/pnas.2312377121 (2024).

62. Getz, W. M., Salter, R. & Mgbara, W. Adequacy of SEIR models when epidemics have spatial structure: Ebola in Sierra Leone. Philosophical Transactions of the Royal Society B 374, 20180282. doi:10.1098/rstb.2018.0282 (2019).

63. Krishnan, R., Cenci, S. & Bourouiba, L. Mitigating bias in estimating epidemic severity due to heterogeneity of epidemic onset and data aggregation. Annals of epidemiology 65, 1–14. doi:10.1016/j.annepidem.2021.07.008 (2022).

64. Zachreson, C., Chang, S., Harding, N. & Prokopenko, M. The effects of local homogeneity assumptions in metapopulation models of infectious disease. Royal Society Open Science 9, 211919. doi:10.1098/rsos.211919 (2022).

65. Shen, N. & Bourouiba, L. Assessing bias in susceptible–infected–recovered estimation from aggregated epidemic data. Royal Society Open Science 12, 240526. doi:10.1098/rsos.240526 (2025).

66. Tegally, H., Wilkinson, E., Tsui, J. L.-H., Moir, M., Martin, D., Brito, A. F., Giovanetti, M., Khan, K., Huber, C., Bogoch, I. I., et al. Dispersal patterns and influence of air travel during the global expansion of SARS-CoV-2 variants of concern. Cell 186, 3277–3290. doi:10.1016/j.cell.2023.06.001 (2023).

67. Google LLC. Google COVID-19 Community Mobility Reports https://www.google.com/covid19/mobility/.

68. Okada, T., Isacchini, G., Yu, Q. & Hallatschek, O. Uncovering heterogeneous intercommunity disease transmission from neutral allele frequency time series. medRxiv, 2024–12. doi:10.1101/2024.12.02.24318370 (2024).

69. Gandon, S. Evolution of multihost parasites. Evolution 58, 455–469 (2004).

70. Lion, S. Class structure, demography, and selection: reproductive-value weighting in nonequilibrium, polymorphic populations. The American Naturalist 191, 620–637. doi:10.1086/696976 (2018).

71. Sah, P., Fitzpatrick, M. C., Zimmer, C. F., Abdollahi, E., Juden-Kelly, L., Moghadas, S. M., Singer, B. H. & Galvani, A. P. Asymptomatic SARS-CoV-2 infection: A systematic review and meta-analysis. Proceedings of the National Academy of Sciences 118, e2109229118. doi:10.1073/pnas.2109229118 (2021).

72. Pastor-Satorras, R., Castellano, C., Van Mieghem, P. & Vespignani, A. Epidemic processes in complex networks. Reviews of modern physics 87, 925–979. doi:10.1103/RevModPhys.87.925 (2015).

73. Murray, J. D. Mathematical biology: II: spatial models and biomedical applications (Springer, 2003).

74. Postnikov, E. B. & Sokolov, I. M. Continuum description of a contact infection spread in a SIR model. Mathematical biosciences 208, 205–215. doi:10.1016/j.mbs.2006.10.004 (2007).

75. Erlander, S. & Stewart, N. F. The gravity model in transportation analysis: theory and extensions (Vsp, 1990).

76. Murray, G. D. & Cliff, A. D. A stochastic model for measles epidemics in a multi-region setting. Transactions of the Institute of British Geographers, 158–174. doi:10.2307/621855 (1977).

77. Xia, Y., Bjørnstad, O. N. & Grenfell, B. T. Measles metapopulation dynamics: a gravity model for epidemiological coupling and dynamics. The American Naturalist 164, 267–281. doi:10.1086/422341 (2004).

78. Balcan, D., Colizza, V., Gonçalves, B., Hu, H., Ramasco, J. J. & Vespignani, A. Multiscale mobility networks and the spatial spreading of infectious diseases. Proceedings of the National Academy of Sciences 106, 21484–21489. doi:10.1073/pnas.0906910106 (2009).

79. McLeod, D. V. & Gandon, S. Effects of epistasis and recombination between vaccineescape and virulence alleles on the dynamics of pathogen adaptation. Nature Ecology & Evolution 6, 786–793. doi:10.1038/s41559-022-01709-y (2022).

80. Gog, J. R., Hill, E. M., Danon, L. & Thompson, R. N. Vaccine escape in a heterogeneous population: insights for SARS-CoV-2 from a simple model. Royal Society Open Science 8, 210530 (2021).

81. Hodcroft, E. B., Zuber, M., Nadeau, S., Vaughan, T. G., Crawford, K. H., Althaus, C. L., Reichmuth, M. L., Bowen, J. E., Walls, A. C., Corti, D., et al. Spread of a SARS-CoV-2 variant through Europe in the summer of 2020. Nature 595, 707–712. doi:10.1038/s41586-021-03677-y (2021).

82. Korber, B., Fischer, W. M., Gnanakaran, S., Yoon, H., Theiler, J., Abfalterer, W., Hengartner, N., Giorgi, E. E., Bhattacharya, T., Foley, B., et al. Tracking changes in SARS-CoV-2 spike: evidence that D614G increases infectivity of the COVID-19 virus. Cell 182, 812–827. doi:10.1016/j.cell.2020.06.043 (2020).

83. Plante, J. A., Liu, Y., Liu, J., Xia, H., Johnson, B. A., Lokugamage, K. G., Zhang, X., Muruato, A. E., Zou, J., Fontes-Garfias, C. R., et al. Spike mutation D614G alters SARS-CoV-2 fitness. Nature 592, 116–121. doi:10.1038/s41586-020-2895-3 (2021).

84. Gandon, S., Hochberg, M. E., Holt, R. D. & Day, T. What limits the evolutionary emergence of pathogens? Philosophical transactions of the Royal Society B: biological sciences 368, 20120086. doi:10.1098/rstb.2012.0086 (2013).

85. Gandon, S., Lambert, A., Voinson, M., Day, T. & Parsons, T. L. The speed of vaccination rollout and the risk of pathogen adaptation. Journal of the Royal Society Interface 22, 20250060. doi:10.1098/rsif.2025.0060 (2025).

86. Lemey, P., Hong, S. L., Hill, V., Baele, G., Poletto, C., Colizza, V., O’toole, Á., Mc-Crone, J. T., Andersen, K. G., Worobey, M., Nelson, M. I., Rambaut, A. & Suchard, M. A. Accommodating individual travel history and unsampled diversity in Bayesian phylogeographic inference of SARS-CoV-2. Nature Communications 11, 5110. doi:10.1038/s41467-020-18877-9 (2020).

87. Butera, Y., Mukantwari, E., Artesi, M., Umuringa, J. d., O’Toole, Á. N., Hill, V., Rooke, S., Hong, S. L., Dellicour, S., Majyambere, O., et al. Genomic sequencing of SARS-CoV-2 in Rwanda reveals the importance of incoming travelers on lineage diversity. Nature Communications 12, 5705. doi:10.1038/s41467-021-25985-7 (2021).

88. Hufnagel, L., Brockmann, D. & Geisel, T. Forecast and control of epidemics in a globalized world. Proceedings of the National Academy of Sciences 101, 15124–15129. doi:10.1073/pnas.0308344101 (2004).

89. Kraemer, M. U., Hill, V., Ruis, C., Dellicour, S., Bajaj, S., McCrone, J. T., Baele, G., Parag, K. V., Battle, A. L., Gutierrez, B., et al. Spatiotemporal invasion dynamics of SARS-CoV-2 lineage B.1.1.7 emergence. Science 373, 889–895. doi:10.1126/science.abj0113 (2021).

90. Volz, E. M., Koelle, K. & Bedford, T. Viral phylodynamics. PLoS computational biology 9, e1002947. doi:10.1371/journal.pcbi.1002947 (2013).

91. Volz, E., Mishra, S., Chand, M., Barrett, J. C., Johnson, R., Geidelberg, L., Hinsley, W. R., Laydon, D. J., Dabrera, G., O’Toole, Á., et al. Assessing transmissibility of SARS-CoV-2 lineage B.1.1.7 in England. Nature 593, 266–269. doi:10.1038/s41586-021-03470-x (2021).

92. Lemey, P., Ruktanonchai, N., Hong, S. L., Colizza, V., Poletto, C., Van den Broeck, F., Gill, M. S., Ji, X., Levasseur, A., Oude Munnink, B. B., et al. Untangling introductions and persistence in COVID-19 resurgence in Europe. Nature 595, 713–717. doi:10.1038/s41586-021-03754-2 (2021).

93. McCrone, J. T., Hill, V., Bajaj, S., Pena, R. E., Lambert, B. C., Inward, R., Bhatt, S., Volz, E., Ruis, C., Dellicour, S., et al. Context-specific emergence and growth of the SARS-CoV-2 Delta variant. Nature 610, 154–160. doi:10.1038/s41586-022-05200-3 (2022).

94. R Core Team. R: A Language and Environment for Statistical Computing R Foundation for Statistical Computing (Vienna, Austria, 2024). https://www.R-project.org/.

95. Soetaert, K., Petzoldt, T. & Setzer, R. W. Solving differential equations in R: package deSolve. Journal of statistical software 33, 1–25. doi:10.18637/jss.v033.i09 (2010).

96. Public Health England. Investigation of novel SARS-COV-2 variant 202012/01: technical briefing 5 tech. rep. (2020). https://assets.publishing.service.gov.uk/government/uploads/system/uploads/attachment_data/file/959426/Variant_of_Concern_VOC_202012_01_Technical_Briefing_5.pdf.

## Supplementary references

1. Lipshtat, A., Alimi, R. & Ben-Horin, Y. Commuting in metapopulation epidemic modeling. Scientific Reports 11, 15198 (2021).

2. Post, W., DeAngelis, D. & Travis, C. Endemic disease in environments with spatially heterogeneous host populations. Mathematical Biosciences 63, 289–302 (1983).

3. McCallum, H., Barlow, N. & Hone, J. How should pathogen transmission be modelled? Trends in ecology & evolution 16, 295–300 (2001).

4. Le Treut, G. et al. A high-resolution flux-matrix model describes the spread of diseases in a spatial network and the effect of mitigation strategies. Scientific Reports 12, 15946 (2022).

5. Benhamou, W., Lion, S., Choquet, R. & Gandon, S. Phenotypic evolution of SARS-CoV-2: a statistical inference approach. Evolution 77, 2213–2223 (2023).

6. Benhamou, W. et al. Evolution of virulence in emerging epidemics: from theory to experimental evolution and back. Virus Evolution 10, veae069 (2024).

7. Priklopil, T. & Lehmann, L. On the interpretation of the operation of natural selection in class-structured populations. The American Naturalist 203, 292–304 (2024).

8. Fisher, R. A. The genetical theory of natural selection: a complete variorum edition (Oxford University Press, 1999).

9. Chevin, L.-M. On measuring selection in experimental evolution. Biology letters 7, 210–213 (2011)

