## Supplementary material for "The interplay between migration and selection on the dynamics of pathogen variants": SI Appendix

$$\begin{aligned} \beta_m^i &= \beta_w^i + \Delta\beta^i \\ \gamma_m^i &= \gamma_w^i + \Delta\gamma^i \end{aligned} \tag{S1}$$

For example, if the efficacy of control measures implemented to reduce transmission in the focal population $i$ is denoted $c^i$, then we would have $\beta_k^i = \left(1 - c^i\right)\beta_k$, with $\beta_k$, the baseline transmissibility of strain $k$.

### S2 Epidemiology

#### S2.1 At the metapopulation level

Epidemiological dynamics at the metapopulation level are given by the following system of ordinary differential equations (ODEs)

$$\begin{aligned}
\frac{\mathrm{d}S}{\mathrm{d}t} &= -\sum_i \lambda^i \Big( \left(1-\mu^i\right) S^i + \mu^j S^j \Big) + \omega R \\
\frac{\mathrm{d}I}{\mathrm{d}t} &= \sum_i \lambda^i \Big( \left(1-\mu^i\right) S^i + \mu^j S^j \Big) - \sum_i \overline{\gamma}^i I^i \\
\frac{\mathrm{d}R}{\mathrm{d}t} &= \sum_i \overline{\gamma}^i I^i - \omega R
\end{aligned} \tag{S2}$$

with $(i, j) \in \{A, B\}^2,\ j \neq i$; $\overline{\gamma}^i$ is the mean recovery rate in population $i$ (averaged across pathogen genotypes)

$$\overline{\gamma}^i = \sum_k \frac{I_k^i}{I^i} \gamma_k^i, \tag{S3}$$

with $k \in \{w, m\}$, and $\lambda^i$ represents the force of infection (i.e., the *per capita* infection rate) experienced in population $i$. Assuming transmission to be frequency-dependent [3], we have

$$\lambda^i = \sum_k \beta_k^i \frac{\left(1-\mu^i\right) I_k^i + \mu^j I_k^j}{\left(1-\mu^i\right) N^i + \mu^j N^j}. \tag{S4}$$

The total force of infection is therefore the sum of the contributions of four types of intra or inter-community interactions [4]. The intra-community contributions correspond to transmissions between a susceptible and an infected host who belong to the same population, which can occur either (i) in the local population or (ii) in the visited population (visitor-to-visitor infection). In contrast, the inter-community contributions correspond to transmissions between a susceptible and an infected host who do not belong to the same population, which can occur (i) when a susceptible visitor gets infected by a native or (ii) when a susceptible native gets infected by a visitor.

#### S2.2 At the level of the focal population

Epidemiological dynamics for the focal population $i \in \{A, B\}$ (accordingly, the non-focal population is denoted by $j$) are given by the following system of ODEs

$$\begin{aligned}
\frac{\mathrm{d}S^i}{\mathrm{d}t} &= -\Big( \left(1-\mu^i\right) \lambda^i + \mu^i \lambda^j \Big) S^i + \omega R^i \\
\frac{\mathrm{d}I^i}{\mathrm{d}t} &= \Big( \left(1-\mu^i\right) \lambda^i + \mu^i \lambda^j \Big) S^i - \overline{\gamma}^i I^i \\
\frac{\mathrm{d}R^i}{\mathrm{d}t} &= \overline{\gamma}^i I^i - \omega R^i
\end{aligned} \tag{S5}$$

with $k \in \{w, m\}$ and with $\lambda^i$ and $\lambda^j$, the forces of infection experienced in the focal and non-focal population, respectively. Focusing only on the epidemiological dynamics of hosts from the focal population $i$ who are infected by the variant, we have

$$\frac{\mathrm{d}I_m^i}{\mathrm{d}t} = \left( \left(1-\mu^i\right) \underbrace{\beta_m^i \frac{\left(1-\mu^i\right) I_m^i + \mu^j I_m^j}{\left(1-\mu^i\right) N^i + \mu^j N^j}}_{\lambda_m^i} + \mu^i \underbrace{\beta_m^j \frac{\left(1-\mu^j\right) I_m^j + \mu^i I_m^i}{\left(1-\mu^j\right) N^j + \mu^i N^i}}_{\lambda_m^j} \right) S^i - \gamma_m^i I_m^i, \tag{S6}$$

with $\lambda_m^i$ and $\lambda_m^j$, the force of infection of the variant in population $i$ and $j$, respectively.

### S3 Evolution

Alongside epidemiological dynamics, we use a population genetics approach and track the evolution of the system over time (strain frequency and spatial differentiation).

#### S3.1 Frequencies of the variant

First, at the level of the focal population $i \in \{A, B\}$, define

$$q^i = \frac{I_m^i}{I^i} = \frac{I_m^i}{\sum_k I_k^i}, \tag{S7}$$

with $k \in \{w, m\}$, the frequency of the variant in the focal population $i$ at the current time $t$. The temporal dynamics of the variant in the focal population $i$ is given by the derivative of $q^i$ with respect to time, that is,

$$\begin{aligned} \frac{\mathrm{d}q^i}{\mathrm{d}t} &= \frac{\mathrm{d}}{\mathrm{d}t}\left(\frac{I_m^i}{I^i}\right) \\ &= \left(\frac{\mathrm{d}I_m^i}{\mathrm{d}t} - q^i \frac{\mathrm{d}I^i}{\mathrm{d}t}\right) \Big/ I^i. \end{aligned} \tag{S8}$$

Second, at the metapopulation level, let $q$ be the overall frequency of the variant

$$q = \frac{I_m}{I} = \frac{\sum_i I_m^i}{\sum_i \sum_k I_k^i} = \sum_i f^i q^i, \tag{S9}$$

where $f^i = I^i/I$ is the frequency of infected hosts that belong to population $i \in \{A, B\}$. Likewise, the temporal dynamics of $q$ is given by

$$\begin{aligned} \frac{\mathrm{d}q}{\mathrm{d}t} &= \frac{\mathrm{d}}{\mathrm{d}t}\left(\frac{I_m}{I}\right) \\ &= \left(\frac{\mathrm{d}I_m}{\mathrm{d}t} - q \frac{\mathrm{d}I}{\mathrm{d}t}\right) \Big/ I. \end{aligned} \tag{S10}$$

#### S3.2 Spatial differentiation

We now introduce the following measure of the genetic differentiation between population $A$ and $B$ (hereafter referred to as spatial differentiation)

$$\mathcal{Q} = \frac{q^A}{1-q^A} \frac{1-q^B}{q^B}, \tag{S11}$$

such that

$$\ln\left(\mathcal{Q}\right) = \mathrm{logit}\left(q^A\right) - \mathrm{logit}\left(q^B\right). \tag{S12}$$

Note that $\ln(\mathcal{Q})$ is only defined when $\left(q^A,\, q^B\right) \in \,]0,\, 1[^2$ (i.e., neither strain has reached fixation). When the variant is more frequent in population $A$ than in population $B$, $\mathcal{Q} > 1$ and $\ln(\mathcal{Q}) > 0$. When the variant is found in equal frequency in both populations, $\mathcal{Q} = 1$ and $\ln(\mathcal{Q}) = 0$ (no differentiation). We used similar quantities in previous work to track the differentiation across pathogen life stages [5, 6], rather than across space, but the principle remains exactly the same. Though the spatial differentiation would be more classically expressed on the natural scale as $q^A - q^B$, the dynamics of this measure is more difficult to interpret because it depends on the genetic variances which are also dynamical variables (see **§S4.2.3**). We thus focus more conveniently on $\mathcal{Q}$. Using equation (S12), the temporal dynamics of the log-differentiation $\ln(\mathcal{Q})$ is given by

$$\frac{\mathrm{d}\ln(\mathcal{Q})}{\mathrm{d}t} = \frac{\mathrm{d}\,\mathrm{logit}\left(q^A\right)}{\mathrm{d}t} - \frac{\mathrm{d}\,\mathrm{logit}\left(q^B\right)}{\mathrm{d}t} = \frac{1}{\mathcal{Q}}\frac{\mathrm{d}\mathcal{Q}}{\mathrm{d}t}, \tag{S13}$$

##### S3.3 Relation between local and global variables

Using either the pair of variables $q^A$ and $q^B$ ("local" variables) or the pair $q$ and $\mathcal{Q}$ ("global" variables) are sufficient to fully describe the evolutionary dynamics of the system. The definitions of $q$ and $\mathcal{Q}$ are based on $q^A$ and $q^B$ and, the other way around, $q^A$ and $q^B$ can be expressed as functions of $q$ and $\mathcal{Q}$

$$\begin{aligned} q^A &= \frac{1 + (\mathcal{Q}-1)\left(f^A + q\right) - \sqrt{\left(1 - \left(q - f^A\right)(\mathcal{Q}-1)\right)^2 + 4f^B q\,(\mathcal{Q}-1)}}{2f^A\,(\mathcal{Q}-1)} \\ q^B &= \frac{-1 - (\mathcal{Q}-1)\left(f^A - q\right) + \sqrt{\left(1 - \left(q - f^A\right)(\mathcal{Q}-1)\right)^2 + 4f^B q\,(\mathcal{Q}-1)}}{2f^B\,(\mathcal{Q}-1)} \end{aligned} \tag{S14}$$

The case $\mathcal{Q} = 1$ is a removable discontinuity for which $q^A = q^B = q$. When the differentiation $\mathcal{Q}$ is not too far from 1 (small differentiation), a Taylor expansion about the case $\mathcal{Q} = 1$ yields the approximations

$$\begin{aligned} q^A &\approx q + q\,(1-q)\left(1 - f^A\right)(\mathcal{Q}-1) \\ q^B &\approx q - q\,(1-q)\left(1 - f^B\right)(\mathcal{Q}-1) \end{aligned} \tag{S15}$$

This also implies that

$$\mathcal{Q} \approx 1 + \frac{q^A - q^B}{q(1-q)}. \tag{S16}$$

#### S4 Weak migration approximation

We now assume that the amount of host mobility is small, i.e., hosts interact much more locally rather than with other populations. In the following, we treat $\mu^i$ as $\epsilon_{\mathcal{M}} M^i$ and $\mu^j$ as $\epsilon_{\mathcal{M}} M^j$ to emphasize that the migration probabilities of the hosts are small, of order $\epsilon_{\mathcal{M}} \ll 1$. We referred to this approximation in the main text as the weak migration assumption.

##### S4.1 Epidemiological dynamics

A Taylor expansion of the force of infection $\lambda^i$ (S4) about $\epsilon_{\mathcal{M}} = 0$ yields

$$\lambda^i = \sum_k \beta_k^i \frac{\left(1 - \frac{N^j}{N^i}\mu^j\right) I_k^i + \mu^j I_k^j}{N^i} + O\left(\epsilon_{\mathcal{M}}^2\right). \tag{S17}$$

Note that the term $O\left(\epsilon_{\mathcal{M}}\right)$ depends on $\mu^j$ but not on $\mu^i$ anymore. Likewise, a Taylor expansion of the ODE system (S5) about $\epsilon_{\mathcal{M}}=0$ yields the following epidemiological dynamics for the infected hosts from population $i$

$$\frac{\mathrm{d}I^i}{\mathrm{d}t}=\sum_k\left(\Bigg[\overbrace{\underbrace{\left(1-\mu^i-\frac{N^j}{N^i}\mu^j\right)\beta_k^i\frac{I_k^i}{N^i}}_{\text{Endogenous}}}^{\text{Intra-community}}+\overbrace{\underbrace{\mu^j\beta_k^i\frac{I_k^j}{N^i}}_{\text{Endogenous}}+\underbrace{\mu^i\beta_k^j\frac{I_k^j}{N^j}}_{\text{Exogenous}}}^{\text{Inter-community}}\Bigg]S^i-\gamma_k^i I_k^i\right)+O\left(\epsilon_{\mathcal{M}}^2\right). \tag{S18}$$

Here, "endogenous" (resp. "exogenous") refers to infections that occur within (resp. outside) the focal population, regardless of the population to which the infector belongs. Note that all the visitor-to-visitor contribution to the force of infection (intra-community but exogenous) is now included in the term $O\left(\epsilon_{\mathcal{M}}^2\right)$.

#### S4.2 Evolutionary dynamics

##### S4.2.1 Dynamics of the variant frequency at the population level

Using equations (S8) and (S18), the dynamics of the variant frequency $q^i$ is given after some rearrangements by

$$\frac{\mathrm{d}q^i}{\mathrm{d}t}=\underbrace{q^i\left(1-q^i\right)}_{\substack{\text{Genetic}\\\text{variance}}}\left(\mathcal{S}^i+\mathcal{H}^i\right)+O\left(\epsilon_{\mathcal{M}}^2\right), \tag{S19}$$

with

$$\begin{aligned}\mathcal{S}^i&=\overbrace{\Delta\beta^i\frac{S^i}{N^i}-\Delta\gamma^i}^{\text{Local selection}}+\overbrace{\left[\mu^i\left(\Delta\beta^j\frac{q^j}{q^i}\frac{I^j/N^j}{I^i/N^i}-\Delta\beta^i\right)+\frac{N^j}{N^i}\mu^j\Delta\beta^i\left(\frac{q^j}{q^i}\frac{I^j/N^j}{I^i/N^i}-1\right)\right]\frac{S^i}{N^i}}^{\text{Interaction between selection and migration}}\\\mathcal{H}^i&=\underbrace{-\frac{q^i-q^j}{q^i\left(1-q^i\right)}\left(\mu^i\beta_w^j+\frac{N^j}{N^i}\mu^j\beta_w^i\right)\frac{I^j/N^j}{I^i/N^i}\frac{S^i}{N^i}}_{\text{Between-pop. homogenization}}\end{aligned} \tag{S20}$$

Both $\mathcal{S}^i$ and $\mathcal{H}^i$ are first-order approximations in $\epsilon_{\mathcal{M}}$. The component $\mathcal{S}^i$ collects all the terms that depend on the phenotypic differences between the two strains and refer to the effects of local selection on the focal population as well as the interaction between selection and migration between populations. The component $\mathcal{H}^i$ is a homogenization force that affects the local dynamics of the variant as soon as the variant frequency varies between the two connected populations (i.e., $q^i\neq q^j$). We obtain a similar partitioning following an approach analogous to [7] (**see details in §S5.2.2**).

The selection coefficient of the variant is typically quantified by estimating the regression slope of its frequency changes on the logit scale [8, 9]. We thus focus instead on the dynamics of the logit-frequency of the variant which is given by

$$\frac{\mathrm{d}\,\mathrm{logit}\left(q^i\right)}{\mathrm{d}t}=\frac{\mathrm{d}q^i/\mathrm{d}t}{q^i\left(1-q^i\right)}=\mathcal{S}^i+\mathcal{H}^i+O\left(\epsilon_{\mathcal{M}}^2\right). \tag{S21}$$

##### S4.2.2 Dynamics of the variant frequency at the metapopulation level

Likewise, the temporal dynamics of the variant logit-frequency at the metapopulation level (S10) is given under the weak migration assumption by

$$\begin{aligned}\frac{\mathrm{d}\,\mathrm{logit}\,(q)}{\mathrm{d}t} =& \frac{f^A q^A \mathcal{S}^A + f^B q^B \mathcal{S}^B}{q} + \\ & \frac{q^A - q^B}{q\,(1-q)} f^A f^B \Bigg[ \left( \frac{\beta_w^A}{N^A} \left( 1 - \mu^A - \frac{N}{N^A}\mu^B \right) - \frac{\beta_w^B}{N^B}\mu^A \right) S^A - \gamma_w^A - \\ & \left( \left( \frac{\beta_w^B}{N^B} \left( 1 - \mu^B - \frac{N}{N^B}\mu^A \right) - \frac{\beta_w^A}{N^A}\mu^B \right) S^B - \gamma_w^B \right) \Bigg] + O\left(\epsilon_{\mathcal{M}}^2\right).\end{aligned} \tag{S22}$$

If selection is homogeneous across space (i.e., $\mathcal{S}^A = \mathcal{S}^B = \mathcal{S}$), then $\left(f^A q^A \mathcal{S}^A + f^B q^B \mathcal{S}^B\right)/q$ is simply $\mathcal{S}$. Besides, in the absence of host mobility (i.e., $\mu^A = \mu^B = 0$), the previous equation reduces to

$$\begin{aligned}\frac{\mathrm{d}\,\mathrm{logit}\,(q)}{\mathrm{d}t} =& \frac{f^A q^A \left( \Delta\beta^A \frac{S^A}{N^A} - \Delta\gamma^A \right) + f^B q^B \left( \Delta\beta^B \frac{S^B}{N^B} - \Delta\gamma^B \right)}{q} + \\ & \frac{q^A - q^B}{q\,(1-q)} f^A f^B \left[ \frac{\beta_w^A}{N^A} S^A - \gamma_w^A - \left( \frac{\beta_w^B}{N^B} S^B - \gamma_w^B \right) \right] + O\left(\epsilon_{\mathcal{M}}^2\right).\end{aligned} \tag{S23}$$

Following an approach similar to [7], we show in **§S5.2.1** how to tease out the effects of selection on the change in frequency.

##### S4.2.3 Dynamics of the spatial differentiation

Using (S19), the dynamics of the difference $q^A - q^B$ is

$$\frac{\mathrm{d}\left(q^A - q^B\right)}{\mathrm{d}t} = q^A\left(1-q^A\right)\left(\mathcal{S}^A + \mathcal{H}^{\mathcal{A}}\right) - q^B\left(1-q^B\right)\left(\mathcal{S}^B + \mathcal{H}^{\mathcal{B}}\right) + O\left(\epsilon_{\mathcal{M}}^2\right), \tag{S24}$$

which depends on the genetic variance $q^i\left(1-q^i\right)$ in each population $i$. Alternatively, we can use the measure of spatial differentiation $\mathcal{Q}$ (as defined in (S11)), whose dynamics on the log scale (S13) can be expressed in a simpler form using equation (S21)

$$\frac{\mathrm{d}\ln\left(\mathcal{Q}\right)}{\mathrm{d}t} = \left(\mathcal{S}^A + \mathcal{H}^{\mathcal{A}}\right) - \left(\mathcal{S}^B + \mathcal{H}^{\mathcal{B}}\right) + O\left(\epsilon_{\mathcal{M}}^2\right), \tag{S25}$$

which gives after some rearrangements of $\mathcal{H}^A - \mathcal{H}^B$

$$\frac{\mathrm{d}\ln\left(\mathcal{Q}\right)}{\mathrm{d}t} = \left(\mathcal{S}^A - \mathcal{S}^B\right) - \left(\mathcal{Q} - 1\right)\left(\frac{\beta_w^A}{N^A}\mu^B + \frac{\beta_w^B}{N^B}\mu^A\right)\left(\frac{q^B}{q^A}\frac{I^B}{I^A}S^A + \frac{1-q^A}{1-q^B}\frac{I^A}{I^B}S^B\right) + O\left(\epsilon_{\mathcal{M}}^2\right). \tag{S26}$$

The differentiation $\mathcal{Q}$ may rapidly reach a quasi-equilibrium value $\hat{\mathcal{Q}}$ when migration is large relative to selection. This quasi-equilibrium can be obtained by setting the right-hand side of (S26) to 0 and solving for $\mathcal{Q}$, which yields

$$\hat{\mathcal{Q}} \approx 1 + \frac{\mathcal{S}^A - \mathcal{S}^B}{\left(\frac{\beta_w^A}{N^A}\mu^B + \frac{\beta_w^B}{N^B}\mu^A\right)\left(\frac{q^B}{q^A}\frac{I^B}{I^A}S^A + \frac{1-q^A}{1-q^B}\frac{I^A}{I^B}S^B\right)}. \tag{S27}$$

This quasi-equilibrium can be viewed as a classical balance between the effects of selection (numerator) and the effects of migration (denominator), and can only be different from 1 when selection is heterogeneous across populations ($\mathcal{S}^A \neq \mathcal{S}^B$).

If the variant is adapted in one population but maladapted in the other, the variant may persist at an intermediate frequency without going to fixation (i.e., polymorphism maintenance) when migration is not too strong (**Fig. S7**). In this case, (S27) is the long-term equilibrium value of $\mathcal{Q}$.

### S5 Relation with Priklopil & Lehmann (2024), *American Naturalist*

In this section, we follow the approach used in [7] to disentangle the effects of "*natural selection*" from the effects of "*class transmission*" on the change in frequency of pathogen variants. We keep the term "*class transmission*", defined in [7] as "*nonheritable fitness differences*", "*which also [affect] allele-frequency change even in the absence of selection*". While the partitioning in [7] is based on a discrete model, we use here a continuous version instead in order to match the framework used in the main text. Similarly to [7], we first start with a general model to describe the dynamics of a class-structure population. Second, we derive the dynamics of the global frequency of a variant, for which we partition selection from class transmission, and apply the obtained decomposition on the model presented in the main text. Last, we do the same but at the level of a single class (i.e., local variant frequency). In the latter, we show that this approach yields results identical to those presented in the main text.

#### S5.1 A general model to describe the dynamics of a class-structure population

We consider here a host population divided between $n \geq 2$ classes, labeled from 1 to $n$ – e.g., in the main text, these classes would represent different populations within a host metapopulation. Let $\mathbf{I}_k = \begin{pmatrix} I_k^1 & I_k^2 & \dots & I_k^n \end{pmatrix}^\top$ be the vector collecting the densities of hosts infected by strain $k \in \{w, m\}$. We model the temporal dynamics of $\mathbf{I}_k$ by the following ODE

$$\dot{\mathbf{I}}_k = \mathbf{R}_k \mathbf{I}_k, \tag{S28}$$

where $\mathbf{R}_k$ is the matrix of transmission/transition rates

$$\mathbf{R}_k = \begin{pmatrix} r_k^{11} & r_k^{12} & \cdots & r_k^{1n} \\ r_k^{21} & r_k^{22} & \cdots & r_k^{2n} \\ \vdots & \vdots & \ddots & \vdots \\ r_k^{n1} & r_k^{n2} & \cdots & r_k^{nn} \end{pmatrix},$$

so that $r_k^{ij}$ is the growth rate of $i$-class individual per $j$-class individual

$$\dot{I}_k^i = \sum_j r_k^{ij} I_k^j. \tag{S29}$$

This equation is the continuous counterpart of equation (1) in [7]. We then also define $I^i = I_w^i + I_m^i$, the total density of infected hosts in population $i$, $I_k = \sum_i I_k^i$, the total density of hosts infected by strain $k$, and $I = \sum_{i,k} I_k^i$, the total density of infected hosts in the metapopulation. Therefore

$$\dot{I}^i = \sum_{j,k} r_k^{ij} I_k^j = \sum_j \underbrace{\left( \left(1 - q^j\right) r_w^{ij} + q^j r_m^{ij} \right)}_{\overline{r}^{ij}} I^j, \tag{S30}$$

$$\dot{I}_k = \sum_{i,j} r_k^{ij} I_k^j = \underbrace{\sum_{i,j} \frac{I_k^j}{I_k} r_k^{ij}}_{\overline{r}_k} I_k, \tag{S31}$$

$$\begin{aligned}\dot{I} &= \sum_{i,j,k} r_k^{ij} I_k^j = \sum_j \Big( \big(1-q^j\big) \sum_i r_w^{ij} + q^j \sum_i r_m^{ij} \Big) I^j \\ &= \underbrace{\sum_j f^j \Big( \big(1-q^j\big) \sum_i r_w^{ij} + q^j \sum_i r_m^{ij} \Big)}_{\overline{r}} I.\end{aligned} \tag{S32}$$

In the previous equations, $\overline{r}^{ij}$ represent the pathogen growth rate (or fitness) in population $i$ due to $j$-class individual, $\overline{r}_k$, the fitness of strain $k$ and $\overline{r}$, the overall fitness of the pathogen; $f^j = I^j/I$ is the proportion of infected hosts in population $j$.

#### S5.2 Partitioning selection and class transmission

##### S5.2.1 Global frequency of the variant

The dynamics of the (global) frequency of the variant $q = I_m/I$ is given by the following ODE

$$\begin{aligned}\dot{q} = \frac{\mathrm{d}}{\mathrm{d}t}\left(\frac{I_m}{I}\right) &= \frac{\dot{I}_m}{I} - q\frac{\dot{I}}{I} \\ &= \frac{\overline{r}_m I_m}{I} - q\frac{\overline{r} I}{I} \\ &= q\left(\overline{r}_m - \overline{r}\right).\end{aligned} \tag{S33}$$

This equation is equivalent to the equation (2) in [7]. After some rearrangements, equation (S33) can also be written as

$$\dot{q} = q(1-q) \sum_{i,j} \left( \frac{I_m^j}{I_m} r_m^{ij} - \frac{I_w^j}{I_w} r_w^{ij} \right). \tag{S34}$$

Note that, in the absence of selection, we have $\forall k,\ r_k^{ij} = r^{ij}$, so that $\overline{r} = \sum_{i,j} f^j r^{ij}$ and $\overline{r}_m = \sum_{i,j} \frac{I_m^j}{I_m} r^{ij}$, which, applied to equation (S33), yields a neutral process where

$$\dot{q} = q \left[ \sum_{i,j} \left( \frac{I_m^j}{I_m} - f^j \right) r^{ij} \right].$$

Similarly to equation (3) in [7], this equation shows that, even in the absence of fitness difference (and thus of selection), class transmissions can still affect the dynamics of the variant frequency $q$; these non-selective changes only arise when there is an asymmetric distribution of the pathogen strains across classes, and stop as soon as $\forall j,\ I_m^j/I_m = f^j$.

To disentangle the effects of selection from those of class transmission, we use the approach presented in [7] and decompose the dynamics of hosts infected by strain $k$ as follows

$$\dot{I}_k = \underbrace{\sum_{i,j} \frac{I_k^j}{I_k} \left( r_k^{ij} - \overline{r}^{ij} \right) I_k}_{\text{Selection}} + \underbrace{\sum_{i,j} \frac{I_k^j}{I_k} \left( \overline{r}^{ij} - \frac{\overline{r}}{n} \right) I_k}_{\text{Class transmission}} + \underbrace{\overline{r} I_k}_{\substack{\text{Pop.}\\ \text{growth}}}, \tag{S35}$$

where the mean $\overline{r}^{ij}$ represents a reference measure of fitness. Equation (S35) is equivalent to equation (S6) in [7]. Plugging (S35) into (S33) yields

$$\begin{aligned}\dot{q} &= \frac{\left(\sum_{i,j}\frac{I_m^j}{I_m}\left(r_m^{ij}-\overline{r}^{ij}\right)I_m+\sum_{i,j}\frac{I_m^j}{I_m}\left(\overline{r}^{ij}-\frac{\overline{r}}{n}\right)I_m+\overline{r}I_m\right)}{I}-q\frac{\overline{r}I}{I}\\ &= \underbrace{q\sum_{i,j}\frac{I_m^j}{I_m}\left(r_m^{ij}-\overline{r}^{ij}\right)}_{\text{Selection}}+\underbrace{q\sum_{i,j}\frac{I_m^j}{I_m}\left(\overline{r}^{ij}-\frac{\overline{r}}{n}\right)}_{\text{Class transmission}},\end{aligned}\tag{S36}$$

which is similar to equation (4) in [7]. Likewise, note that the last term in (S35) (population growth) cancels out in (S36) and thus does not contribute to the dynamics of $q$. Summing over all strains of the pathogen, we can verify that natural selection is indeed a conservative evolutionary force (i.e., if one strain increases in frequency, the other must decrease)

$$\sum_k\frac{I_k}{I}\sum_{i,j}\frac{I_k^j}{I_k}\left(r_k^{ij}-\overline{r}^{ij}\right)\;=\;\sum_{i,j}f^j\underbrace{\sum_k\frac{I_k^j}{I^j}r_k^{ij}}_{\overline{r}^{ij}}-\sum_{i,j}f^j\overline{r}^{ij}\;=\;\sum_{i,j}f^j\left(\overline{r}^{ij}-\overline{r}^{ij}\right)\;=\;0.$$

We then apply the partitioning (S36) to the model presented in the main text, for which we have $\forall(i,j)\in\{A,B\}^2,\ j\neq i$

$$\begin{aligned}r_k^{ii} &= \left(\left(1-\mu^i\right)\frac{\left(1-\mu^i\right)\beta_k^i}{\left(1-\mu^i\right)N^i+\mu^jN^j}+\mu^i\frac{\mu^i\beta_k^j}{\left(1-\mu^j\right)N^j+\mu^iN^i}\right)S^i-\gamma_k^i\\ r_k^{ij} &= \left(\left(1-\mu^i\right)\frac{\mu^j\beta_k^i}{\left(1-\mu^i\right)N^i+\mu^jN^j}+\mu^i\frac{\left(1-\mu^j\right)\beta_k^j}{\left(1-\mu^j\right)N^j+\mu^iN^i}\right)S^i\end{aligned}\tag{S37}$$

Under the assumption of weak migration, a Taylor expansion yields

$$\begin{aligned}r_k^{ii} &= \left(\left(1-\mu^i\right)\frac{\beta_k^i}{\left(1-\mu^i\right)N^i+\mu^jN^j}-\mu^i\frac{\beta_k^i}{\left(1-\mu^i\right)N^i+\mu^jN^j}\right)S^i-\gamma_k^i+O\left(\epsilon_{\mathcal{M}}^2\right)\\ r_k^{ij} &= \left(\mu^j\frac{\beta_k^i}{\left(1-\mu^i\right)N^i+\mu^jN^j}+\mu^i\frac{\beta_k^j}{\left(1-\mu^j\right)N^j+\mu^iN^i}\right)S^i+O\left(\epsilon_{\mathcal{M}}^2\right)\end{aligned}\tag{S38}$$

Using $\overline{r}^{ij}=\left(\left(1-q^j\right)r_w^{ij}+q^jr_m^{ij}\right)$ and plugging (S38) into (S36) yields the selection component

$$\begin{aligned}q\sum_{i,j}\frac{I_m^j}{I_m}\left(r_m^{ij}-\overline{r}^{ij}\right) &= f^Aq^A\left(1-q^A\right)\Bigg[\Delta\beta^A\frac{S^A}{N^A}-\Delta\gamma^A+\\ &\qquad\left(\mu^A\left(\Delta\beta^B\frac{q^B}{q^A}\frac{I^B/N^B}{I^A/N^A}\frac{1-q^B}{1-q^A}-\Delta\beta^A\right)+\frac{N^B}{N^A}\mu^B\Delta\beta^A\left(\frac{q^B}{q^A}\frac{I^B/N^B}{I^A/N^A}\frac{1-q^B}{1-q^A}-1\right)\right)\frac{S^A}{N^A}\Bigg]+\\ &\quad f^Bq^B\left(1-q^B\right)\Bigg[\Delta\beta^B\frac{S^B}{N^B}-\Delta\gamma^B+\\ &\qquad\left(\mu^B\left(\Delta\beta^A\frac{q^A}{q^B}\frac{I^A/N^A}{I^B/N^B}\frac{1-q^A}{1-q^B}-\Delta\beta^B\right)+\frac{N^A}{N^B}\mu^A\Delta\beta^B\left(\frac{q^A}{q^B}\frac{I^A/N^A}{I^B/N^B}\frac{1-q^A}{1-q^B}-1\right)\right)\frac{S^B}{N^B}\Bigg]\\ &\quad+O\left(\epsilon_{\mathcal{M}}^2\right),\end{aligned}\tag{S39}$$

and the class-transmission component

$$
\begin{aligned}
q\sum_{i,j}\frac{I_m^j}{I_m}\left(\overline{r}^{ij}-\frac{\overline{r}}{2}\right) = & \left(q^A-q^B\right)f^Af^B\times \\
& \Bigg[\left(\left(1-\mu^A-\frac{N^B}{N^A}\mu^B\right)\left(\beta_w^A+q^A\Delta\beta^A\right)-\mu^B\left(\beta_w^A+q^B\Delta\beta^A\right)-\frac{N^A}{N^B}\mu^A\left(\beta_w^B+q^B\Delta\beta^B\right)\right)\frac{S^A}{N^A} \\
& -\left(\left(1-\mu^B-\frac{N^A}{N^B}\mu^A\right)\left(\beta_w^B+q^B\Delta\beta^B\right)-\mu^A\left(\beta_w^B+q^A\Delta\beta^B\right)-\frac{N^B}{N^A}\mu^B\left(\beta_w^A+q^A\Delta\beta^A\right)\right)\frac{S^B}{N^B} \\
& -\left(\left(\gamma_w^A+q^A\Delta\gamma^A\right)-\left(\gamma_w^B+q^B\Delta\gamma^B\right)\right)\Bigg]+O\left(\epsilon_{\mathcal{M}}^2\right).
\end{aligned}
\tag{S40}
$$

Note that the terms

$$
\begin{aligned}
\beta_w^i+q^i\Delta\beta^i &= (1-q^i)\beta_w^i+q^i\beta_m^i \\
\beta_w^i+q^j\Delta\beta^i &= (1-q^j)\beta_w^i+q^j\beta_m^i \\
\gamma_w^i+q^i\Delta\gamma^i &= (1-q^i)\gamma_w^i+q^i\gamma_m^i
\end{aligned}
$$

correspond to mean trait values. For the transmission rate $\beta$, these mean trait values can be averaged across the genotype distribution of the pathogen in either population ($i$ or $j$) depending on where the infection occurs.

##### S5.2.2 Local frequency of the variant

We now focus on the local variant frequency, i.e., at the level of a single class $i$. The dynamics of the local frequency of the variant $q^i = I_m^i/I^i$ is given by the following ODE

$$
\dot{I}_k^i = \underbrace{\sum_j \frac{I_k^j}{I_k^i}\left(r_k^{ij}-\overline{r}^{ij}\right)I_k^i}_{\text{Selection}} + \underbrace{\sum_j \frac{I_k^j}{I_k^i}\left(\overline{r}^{ij}-\frac{I_k^i}{I_k}\overline{r}^i\right)I_k^i}_{\text{Class transmission}} + \underbrace{\overline{r}^iI_k^i}_{\substack{\text{Pop.}\\\text{growth}}},
\tag{S41}
$$

with

$$
\overline{r}^i \;=\; \sum_{j,k} r_k^{ij}\frac{I_k^j}{I^i} \;=\; \sum_j\left(\left(1-q^j\right)r_w^{ij}+q^jr_m^{ij}\right)\frac{I^j}{I^i}
\tag{S42}
$$

and where, again, the mean $\overline{r}^{ij}$ represents a reference measure of fitness. Thus

$$
\begin{aligned}
\dot{q}^i &= \frac{\dot{I}_m^i}{I^i}-q^i\frac{\dot{I}^i}{I^i} \\
&= \frac{\sum_j \frac{I_m^j}{I_m^i}\left(r_m^{ij}-\overline{r}^{ij}\right)I_m^i+\sum_j\frac{I_m^j}{I_m^i}\left(\overline{r}^{ij}-\frac{I_m^i}{I_m}\overline{r}^i\right)I_m^i+\overline{r}^iI_m^i}{I^i}-q^i\frac{\overline{r}^iI^i}{I^i} \\
&= \underbrace{q^i\sum_j\frac{I_m^j}{I_m^i}\left(r_m^{ij}-\overline{r}^{ij}\right)}_{\text{Selection}}+\underbrace{q^i\sum_j\frac{I_m^j}{I_m^i}\left(\overline{r}^{ij}-\frac{I_m^i}{I_m}\overline{r}^i\right)}_{\text{Class transmission}}.
\end{aligned}
\tag{S43}
$$

Again, the last term in (S41) (population growth) cancels out in (S44) and we can also verify that the effects of natural selection, summed over all strains of the pathogen, is null

$$\sum_k \frac{I_k^i}{I^i} \sum_j \frac{I_k^j}{I_k^i} \left( r_k^{ij} - \overline{r}^{ij} \right) \; = \; \sum_{j,k} \frac{I_k^j}{I^i} r_k^{ij} - \sum_j \frac{\overline{r}^{ij}}{I^i} \sum_k I_k^j \; = \; \overline{r}^i - \sum_j \frac{I^j}{I^i} \overline{r}^{ij} \; = \; \overline{r}^i - \overline{r}^i \; = \; 0$$

After some rearrangements, we have

$$\dot{q}^i = \underbrace{q^i \left(1 - q^i\right) \sum_j \frac{I_m^j}{I_m^i} \left( r_m^{ij} - r_w^{ij} \right)}_{\text{Selection}} + \underbrace{q^i \sum_j \frac{I_m^j}{I_m^i} \left[ \left(1 - q^j\right) r_w^{ij} + q^j r_m^{ij} - \frac{I_m^i}{I_m} \sum_j \left( \left(1 - q^j\right) r_w^{ij} + q^j r_m^{ij} \right) \frac{I^j}{I^i} \right]}_{\text{Class transmission}} \tag{S44}$$

We then apply the partitioning (S44) to the model presented in the main text. Using the growth rates given in equations (S38), we have

$$q^i \left(1 - q^i\right) \sum_j \frac{I_m^j}{I_m^i} \left( r_m^{ij} - r_w^{ij} \right) =$$

$$q^i \left(1 - q^i\right) \underbrace{\left( \Delta\beta^i \frac{S^i}{N^i} - \Delta\gamma^i + \left[ \mu^i \left( \Delta\beta^j \frac{q^j}{q^i} \frac{I^j/N^j}{I^i/N^i} - \Delta\beta^i \right) + \frac{N^j}{N^i} \mu^j \Delta\beta^i \left( \frac{q^j}{q^i} \frac{I^j/N^j}{I^i/N^i} - 1 \right) \right] \frac{S^i}{N^i} \right)}_{\mathcal{S}^i} + O\left(\epsilon_{\mathcal{M}}^2\right)$$

$$q^i \sum_j \frac{I_m^j}{I_m^i} \left[ \left(1 - q^j\right) r_w^{ij} + q^j r_m^{ij} - \frac{I_m^i}{I_m} \sum_j \left( \left(1 - q^j\right) r_w^{ij} + q^j r_m^{ij} \right) \frac{I^j}{I^i} \right] = \underbrace{- \left(q^i - q^j\right) \left( \mu^i \beta_w^j + \frac{N^j}{N^i} \mu^j \beta_w^i \right) \frac{I^j/N^j}{I^i/N^i} \frac{S^i}{N^i}}_{q^i(1-q^i)\mathcal{H}^i} + O\left(\epsilon_{\mathcal{M}}^2\right)$$

We thus recover the results derived in the main text (the only difference being that our formulation scales each component by the genetic variance $q^i\left(1 - q^i\right)$)

$$\dot{q}^i = q^i \left(1 - q^i\right) \left(\mathcal{S}^i + \mathcal{H}^i\right) + O\left(\epsilon_{\mathcal{M}}^2\right). \tag{S45}$$

### S6 Model extensions

#### S6.1 Extension to an arbitrary number of populations

More generally, we can extend the previous model to $n \geq 2$ populations.

##### S6.1.1 Epidemiological dynamics

At the level of the focal population $i$, the dynamics of the density of infected hosts in model (S5) becomes

$$\frac{\mathrm{d}I^i}{\mathrm{d}t} = \left[ \left( 1 - \sum_{j \neq i} \mu^{i \to j} \right) \lambda^i + \sum_{j \neq i} \mu^{i \to j} \lambda^j \right] S^i - \overline{\gamma}^i I^i, \tag{S46}$$

with $k \in \{w, m\}$ and where $j$ refers to the index of the $j$th population and $\mu^{i \to j}$ (resp. $\mu^{j \to i}$), to the probability of migration from the focal population $i$ (resp. from the non-focal population $j$) to population $j$ (resp. $i$). As before,

$\lambda^i$ and $\lambda^j$ are the forces of infection experienced in population $i$ and $j$, respectively, such that

$$\lambda^i = \sum_k \beta_k^i \frac{\left(1 - \sum_{j \neq i} \mu^{i \to j}\right) I_k^i + \sum_{j \neq i} \mu^{j \to i} I_k^j}{\left(1 - \sum_{j \neq i} \mu^{i \to j}\right) N^i + \sum_{j \neq i} \mu^{j \to i} N^j}. \tag{S47}$$

Under the weak migration assumption, a Taylor expansion of the previous equation about $\epsilon_{\mathcal{M}} = 0$ (see details in **§S4**) yields

$$\lambda^i = \sum_k \beta_k^i \frac{\left(1 - \sum_{j \neq i} \frac{N^j}{N^i} \mu^{j \to i}\right) I_k^i + \sum_{j \neq i} \mu^{j \to i} I_k^j}{N^i} + O\left(\epsilon_{\mathcal{M}}^2\right),$$

and, similarly, the dynamics of the density of infected hosts (S46) is now given by

$$\frac{\mathrm{d}I^i}{\mathrm{d}t} = \sum_k \left[ \overbrace{\underbrace{\left(1 - \sum_{j \neq i} \mu^{i \to j} - \sum_{j \neq i} \frac{N^j}{N^i} \mu^{j \to i}\right) \beta_k^i \frac{I_k^i}{N^i}}_{\text{Endogenous}}}^{\text{Intra-community}} + \overbrace{\underbrace{\sum_{j \neq i} \mu^{j \to i} \beta_k^i \frac{I_k^j}{N^i}}_{\text{Endogenous}} + \underbrace{\sum_{i \neq j} \mu^{i \to j} \beta_k^j \frac{I_k^i}{N^j}}_{\text{Exogenous}}}^{\text{Inter-community}} \right] S^i - \sum_k \gamma_k^i I_k^i + O\left(\epsilon_{\mathcal{M}}^2\right).$$

##### S6.1.2 Evolutionary dynamics

At the level of the focal population $i$, the dynamics of the variant logit-frequency $\text{logit}\left(q^i\right) = \text{logit}\left(I_m^i / I^i\right)$ can still be expressed as (S21), but now with

$$\begin{aligned} \mathcal{S}^i &= \Delta\beta^i \frac{S^i}{N^i} - \Delta\gamma^i + \left[ \sum_{j \neq i} \mu^{i \to j} \left( \Delta\beta^j \frac{q^j}{q^i} \frac{I^j / N^j}{I^i / N^i} - \Delta\beta^i \right) + \Delta\beta^i \sum_{j \neq i} \frac{N^j}{N^i} \mu^{j \to i} \left( \frac{q^j}{q^i} \frac{I^j / N^j}{I^i / N^i} - 1 \right) \right] \frac{S^i}{N^i} \\ \mathcal{H}^i &= -\sum_{j \neq i} \frac{q^i - q^j}{q^i \left(1 - q^i\right)} \left( \beta_w^j \mu^{i \to j} + \frac{N^j}{N^i} \beta_w^i \mu^{j \to i} \right) \frac{I^j / N^j}{I^i / N^i} \frac{S^i}{N^i} \end{aligned} \tag{S48}$$

We illustrate the dynamics of the logit-frequency of an emerging variant in a nine-patch host metapopulation (such as the nine regions of England) in **Fig. S3**.

We can also define the genetic differentiation between any pair of populations, say $i$ and $l \neq i$, as

$$\mathcal{Q}^{il} = \frac{q^i}{1 - q^i} \frac{1 - q^l}{q^l},$$

such that $\ln\left(\mathcal{Q}^{il}\right) = \text{logit}\left(q^i\right) - \text{logit}\left(q^l\right)$. Again, under the assumption of weak migration, the dynamics of the spatial log-differentiation $\ln\left(\mathcal{Q}^{il}\right)$ is given by

$$\frac{\mathrm{d} \ln\left(\mathcal{Q}^{il}\right)}{\mathrm{d}t} = \left(\mathcal{S}^i - \mathcal{S}^l\right) + \left(\mathcal{H}^i - \mathcal{H}^l\right) + O\left(\epsilon_{\mathcal{M}}^2\right),$$

which, after some rearrangements, yields

$$\frac{\mathrm{d}\ln\left(\mathcal{Q}^{il}\right)}{\mathrm{d}t} = \left(\mathcal{S}^i - \mathcal{S}^l\right) - \sum_{j\neq i}\left(\mathcal{Q}^{ij}-1\right)\left(\mu^{j\to i}\frac{\beta_w^i}{N^i} + \mu^{i\to j}\frac{\beta_w^j}{N^j}\right)\frac{q^j}{q^i}\frac{I^j}{I^i}S^i$$
$$- \sum_{j\neq l}\left(\mathcal{Q}^{jl}-1\right)\left(\mu^{j\to l}\frac{\beta_w^l}{N^l} + \mu^{l\to j}\frac{\beta_w^j}{N^j}\right)\frac{1-q^j}{1-q^l}\frac{I^j}{I^l}S^l + O\left(\epsilon_{\mathcal{M}}^2\right).$$

This expression can be difficult to use in practice as the dynamics of the spatial differentiation between two populations now depends not only on itself, but on the whole spatial differentiation structure in which populations $i$ and $l$ are embedded, including all the corresponding pairwise differentiation values. The weight of each corresponding pair of populations can either speed up or slow down the overall dynamics of the pairwise spatial differentiation.

#### S6.2 Extension to reduced mobility in infected hosts

In this section, we relax the assumption that infected hosts ($I$) commute with the same probability that non-infected hosts ($S$ and $R$). To do so, we assume that infected individuals from the focal population $i$ can commute to the non-focal population with probability $(1-\zeta)\mu^i$, where $\zeta \in [0, 1]$ represents the reduction in host mobility due to the disease. For the sake of simplicity, $\zeta$ is assumed to be the same for both populations and for both strains. Note that the case $\zeta = 0$ corresponds thus to the scenario presented in the main text, in which the infection does not affect host mobility.

##### S6.2.1 Epidemiological dynamics

In these conditions, the ODE systems (S2) and (S5) are unchanged but the force of infection in the focal population $i$ is now given by

$$\begin{aligned}\lambda^i &= \sum_k \beta_k^i \frac{\left(1-(1-\zeta)\mu^i\right)I_k^i + (1-\zeta)\mu^j I_k^j}{N^i - \mu^i\left(S^i + (1-\zeta)I^i + R^i\right) + \mu^j\left(S^j + (1-\zeta)I^j + R^j\right)} \\ &= \sum_k \beta_k^i \frac{\left[1-\frac{N^j}{N^i}\mu^j\left(1-\zeta\frac{I^j}{N^j}\right) + \zeta\mu^i\left(1-\frac{I^i}{N^i}\right)\right]I_k^i + (1-\zeta)\mu^j I_k^j}{N^i} + O\left(\epsilon_{\mathcal{M}}^2\right),\end{aligned} \tag{S49}$$

where the second line is a Taylor expansion of the first line under the assumption of the weak migration (see details in **§S4**). Plugging equation (S49) into the ODE system (S5), the dynamics of the density of infected hosts thus becomes

$$\begin{aligned}\frac{\mathrm{d}I^i}{\mathrm{d}t} =& \sum_k \Bigg[\overbrace{\underbrace{\left(\left(1-\mu^i\left(1-\zeta\left(1-\frac{I^i}{N^i}\right)\right)-\frac{N^j}{N^i}\mu^j\left(1-\zeta\frac{I^j}{N^j}\right)\right)\beta_k^i\frac{I_k^i}{N^i}}_{\text{Endogenous}}}^{\text{Intra-community}} + \overbrace{\underbrace{(1-\zeta)\mu^j\beta_k^i\frac{I_k^j}{N^i}}_{\text{Endogenous}} + \underbrace{\mu^i\beta_k^j\frac{I_k^j}{N^j}}_{\text{Exogenous}}}^{\text{Inter-community}}\Bigg]S^i \\ &- \sum_k \gamma_k^i I_k^i + O\left(\epsilon_{\mathcal{M}}^2\right).\end{aligned} \tag{S50}$$

Note that, if $\zeta = 1$ – i.e., infection prevents infected hosts from commuting between populations –, the endogenous inter-community term disappears because inter-community transmissions would solely occur through the commuting of susceptible hosts (infection in the non-focal population and pathogen importation in the focal population).

###### S6.2.2 Evolutionary dynamics

At the level of the focal population $i$, the dynamics of the variant logit-frequency can still be expressed as (S21), but now with

$$\mathcal{S}^i = \overbrace{\Delta\beta^i \frac{S^i}{N^i} - \Delta\gamma^i}^{\text{Local selection}} + \overbrace{\left[\mu^i \left(\Delta\beta^j \frac{q^j}{q^i}\frac{I^j/N^j}{I^i/N^i} - \Delta\beta^i\left(1-\zeta\left(1-\frac{I^i}{N^i}\right)\right)\right) + \frac{N^j}{N^i}\mu^j\Delta\beta^i\left((1-\zeta)\frac{q^j}{q^i}\frac{I^j/N^j}{I^i/N^i} + \zeta\frac{I^j}{N^j} - 1\right)\right]\frac{S^i}{N^i}}^{\text{Interaction between selection and migration}} \tag{S51}$$

$$\mathcal{H}^i = \underbrace{-\frac{q^i - q^j}{q^i\left(1-q^i\right)}\left(\mu^i\beta_w^j + \frac{N^j}{N^i}(1-\zeta)\mu^j\beta_w^i\right)\frac{I^j/N^j}{I^i/N^i}\frac{S^i}{N^i}}_{\text{Between-pop. homogenization}}$$

When $\zeta = 0$ (no reduced mobility in infected hosts), we recover equations (S20), as presented in the main text. At the other end of the spectrum, when $\zeta = 1$, equation (S51) reduces to

$$\begin{aligned}
\mathcal{S}^i &= \Delta\beta^i \frac{S^i}{N^i} - \Delta\gamma^i + \left[\mu^i\left(\Delta\beta^j\frac{q^j}{q^i}\frac{I^j/N^j}{I^i/N^i} - \Delta\beta^i\frac{I^i}{N^i}\right) + \frac{N^j}{N^i}\mu^j\Delta\beta^i\left(\frac{I^j}{N^j} - 1\right)\right]\frac{S^i}{N^i} \\
\mathcal{H}^i &= -\frac{q^i - q^j}{q^i\left(1-q^i\right)}\mu^i\beta_w^j\frac{I^j/N^j}{I^i/N^i}\frac{S^i}{N^i}
\end{aligned}$$

Again, the homogenization component $\mathcal{H}^i$ now only depends on $\mu^i$ because inter-community transmissions can only be achieved through the mobility of susceptible hosts. In **Fig. S9-A**, we plot some examples for the dynamics of the variant logit-frequency by varying the value of $\zeta$, from 0 to 1. Simulated trajectories are extremely similar, in particular because the proportion of infected hosts remains really small compared to the proportion of susceptibles.

Besides, the dynamics of the spatial log-differentiation $\ln(\mathcal{Q}) = \text{logit}\left(q^A\right) - \text{logit}\left(q^B\right)$ is now given by

$$\begin{aligned}
\frac{\mathrm{d}\ln(\mathcal{Q})}{\mathrm{d}t} = \left(\mathcal{S}^A - \mathcal{S}^B\right) - (\mathcal{Q}-1)\Bigg[&\mu^B\frac{\beta_w^A}{N^A}\left((1-\zeta)\frac{q^B}{q^A}\frac{I^B}{I^A}S^A + \frac{1-q^A}{1-q^B}\frac{I^A}{I^B}S^B\right) + \\
&\mu^A\frac{\beta_w^B}{N^B}\left(\frac{q^B}{q^A}\frac{I^B}{I^A}S^A + (1-\zeta)\frac{1-q^A}{1-q^B}\frac{I^A}{I^B}S^B\right)\Bigg] + O\left(\epsilon_{\mathcal{M}}^2\right).
\end{aligned} \tag{S52}$$

Again, when $\zeta = 0$, we recover equations (S26). We show in **Fig. S9-B** some simulated dynamics for the spatial log-differentiation by varying the value of $\zeta$, from 0 to 1. As expected, lower amounts of migration, due to reduced mobility rates in infected hosts ($\zeta \to 1$), slow down the homogenization process between the two populations; note however that, again, this effect is small because the proportion of infected hosts is small.

##### S6.3 Extension to partial cross-immunity

In this section, we extend the model (S5)-(S6) to account for partial cross-immunity. In the previous model, we assume that immunity provided by one strain also provides full protection against the other (full cross-immunity). By relaxing this assumption, we now need to take into account possible reinfections of recovered individuals by another strain. Because increasing the number of infected classes makes theoretical derivations much more complicated and difficult to compare with our previous model, we assume for simplicity that only the variant can partially escape the immunity provided by the wildtype. Define $\sigma_m^i \in [0,\, 1]$ as the ability of the variant to infect hosts from population $i$ who acquired immunity to the wildtype (denoted by $R_w^i$), where $\sigma_m^i = 0$ indicates full cross-immunity (as assumed in the main text) and $\sigma_m^i = 1$ indicates no-cross immunity. Note that, since we assume $\sigma_w^i = 0$, the phenotypic difference $\Delta\sigma^i = \sigma_m^i - \sigma_w^i$ reduces to $\sigma_m^i$ in this very special case. The dynamics of $I_w^i$ thus remains unchanged but that of $I_m^i$ becomes

$$\frac{\mathrm{d}I_m^i}{\mathrm{d}t} = \Big( \left(1 - \mu^i\right) \lambda_m^i + \mu^i \lambda_m^j \Big) \left(S^i + \sigma_m^i R_w^i\right) - \gamma_m^i I_m^i. \tag{S53}$$

Then, the selection component $\mathcal{S}^i$ of the dynamics of the variant logit-frequency $\text{logit}\left(q^i\right)$ is given by

$$\begin{aligned}\mathcal{S}^i = \underbrace{\Delta\beta^i \frac{S^i}{N^i} + \sigma_m^i \beta_m^i \frac{R_w^i}{N^i} - \Delta\gamma^i}_{\text{Local selection}} & \overbrace{+ \left[\mu^i \left(\Delta\beta^j \frac{q^j}{q^i} \frac{I^j/N^j}{I^i/N^i} - \Delta\beta^i\right) + \frac{N^j}{N^i} \mu^j \Delta\beta^i \left(\frac{q^j}{q^i} \frac{I^j/N^j}{I^i/N^i} - 1\right)\right] \frac{S^i}{N^i}}^{\text{Interaction between selection and migration}} \\ & + \left[\mu^i \left(\beta_m^j \frac{q^j}{q^i} \frac{I^j/N^j}{I^i/N^i} - \beta_m^i\right) + \frac{N^j}{N^i} \mu^j \beta_m^i \left(\frac{q^j}{q^i} \frac{I^j/N^j}{I^i/N^i} - 1\right)\right] \sigma_m^i \frac{R_w^i}{N^i},\end{aligned} \tag{S54}$$

while the homogenization component $\mathcal{H}^i$ remains the same as in (S20) (and, therefore, the dynamics of the spatial differentiation can still be written as (S26)).

### Supplementary references

1. Lipshtat, A., Alimi, R. & Ben-Horin, Y. Commuting in metapopulation epidemic modeling. *Scientific Reports* **11,** 15198 (2021).
2. Post, W., DeAngelis, D. & Travis, C. Endemic disease in environments with spatially heterogeneous host populations. *Mathematical Biosciences* **63,** 289–302 (1983).
3. McCallum, H., Barlow, N. & Hone, J. How should pathogen transmission be modelled? *Trends in ecology & evolution* **16,** 295–300 (2001).
4. Le Treut, G. *et al.* A high-resolution flux-matrix model describes the spread of diseases in a spatial network and the effect of mitigation strategies. *Scientific Reports* **12,** 15946 (2022).
5. Benhamou, W., Lion, S., Choquet, R. & Gandon, S. Phenotypic evolution of SARS-CoV-2: a statistical inference approach. *Evolution* **77,** 2213–2223 (2023).
6. Benhamou, W. *et al.* Evolution of virulence in emerging epidemics: from theory to experimental evolution and back. *Virus Evolution* **10,** veae069 (2024).
7. Priklopil, T. & Lehmann, L. On the interpretation of the operation of natural selection in class-structured populations. *The American Naturalist* **203,** 292–304 (2024).
8. Fisher, R. A. *The genetical theory of natural selection: a complete variorum edition* (Oxford University Press, 1999).
9. Chevin, L.-M. On measuring selection in experimental evolution. *Biology letters* **7,** 210–213 (2011).
