## Supplementary figures for "The interplay between migration and selection on the dynamics of pathogen variants"

November 18, 2025

### List of supplementary figures (pp. 2-11)

– **Figure S1:** Delays between emergence of the SARS-CoV-2 Alpha and Delta variants across regions of England.

– **Figure S2:** Evolutionary dynamics of an emerging variant in a nine-patch host metapopulation.

– **Figure S3:** Migration can steepen the change of local frequency of an emerging variant.

– **Figure S4:** The effect of population size asymmetry on the dynamics of the variant frequency in the focal population.

– **Figure S5:** Short-term dynamics of spatial differentiation and the migration–selection balance.

– **Figure S6:** The transient disruption of spatial differentiation.

– **Figure S7:** Long-term spatial differentiation and local adaptation.

– **Figure S8:** Approximations of the early dynamics of the spatial differentiation.

– **Figure S9:** Change in visitors in the United Kingdom (Google mobility reports).

– **Figure S10:** The effect of reduced mobility in infected hosts on the evolutionary dynamics.

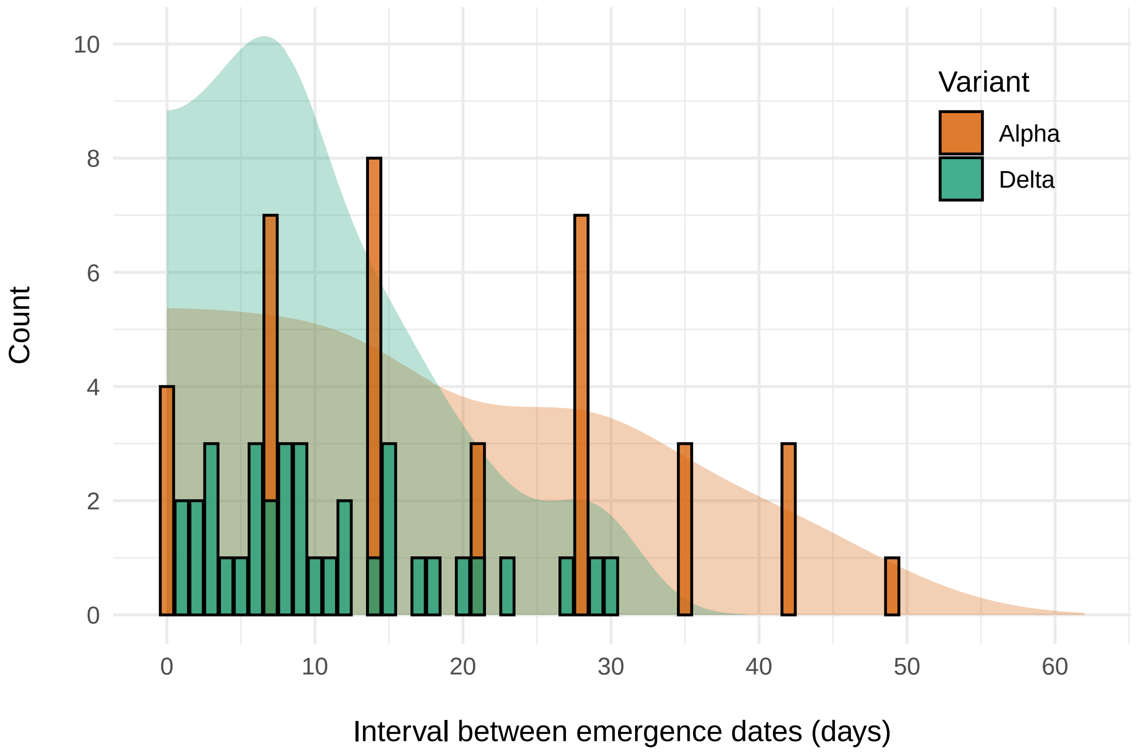


Figure S1: **Delays between emergence of the SARS-CoV-2 Alpha and Delta variants across regions of England.** We compute pairwise time differences between emergence dates from **Fig. 1** and plot the corresponding distributions (count and density) for both variants. In each case, the time of introduction of the new variant differs substantially among regions, but delays between successive introductions are longer on average for Alpha. The latter pattern might be induced by a weaker interregional connectivity (i.e., fewer migration events between regions).

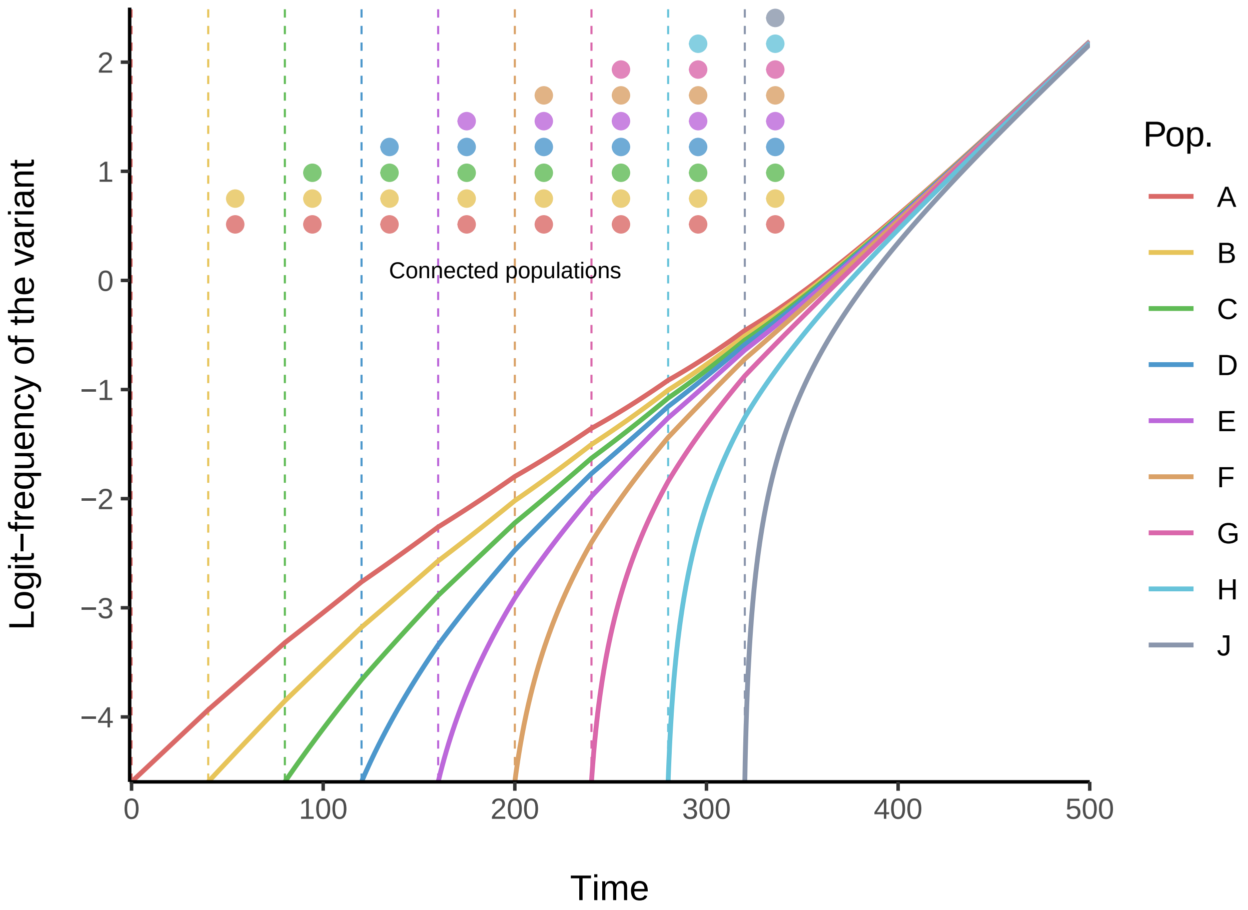


Figure S2: **Evolutionary dynamics of an emerging variant in a nine-patch host metapopulation.** We extend in **SI Appendix §S6.1** the model presented in the main text to any number of populations. Here, the host metapopulation is divided in nine populations (such as the nine regions of England). For the sake of simplicity, the variant has only a transmission advantage and the phenotypes of both strains do not vary across populations: $\beta_w = 0.11$, $\beta_m = 0.1265$ (+15% transmission advantage), $\gamma_w = \gamma_m = 0.1$ and $\omega = 0$. We also assume same total density for each population (100) and same probability of migration (0.01). At $t = 0$, the wildtype is introduced at low density ($10^{-7}$) in all populations, but the variant is only introduced in population $A$ at a lower density (1% of infected hosts $I^A$) – all populations are otherwise fully susceptible. All populations are first isolated from each other. Populations are then sequentially connected by migration, starting from population $A$ and $B$. Every time a population is connected (as indicated by the vertical dashed lines), we introduce the variant such that it represents 1% of the infected hosts in that population.

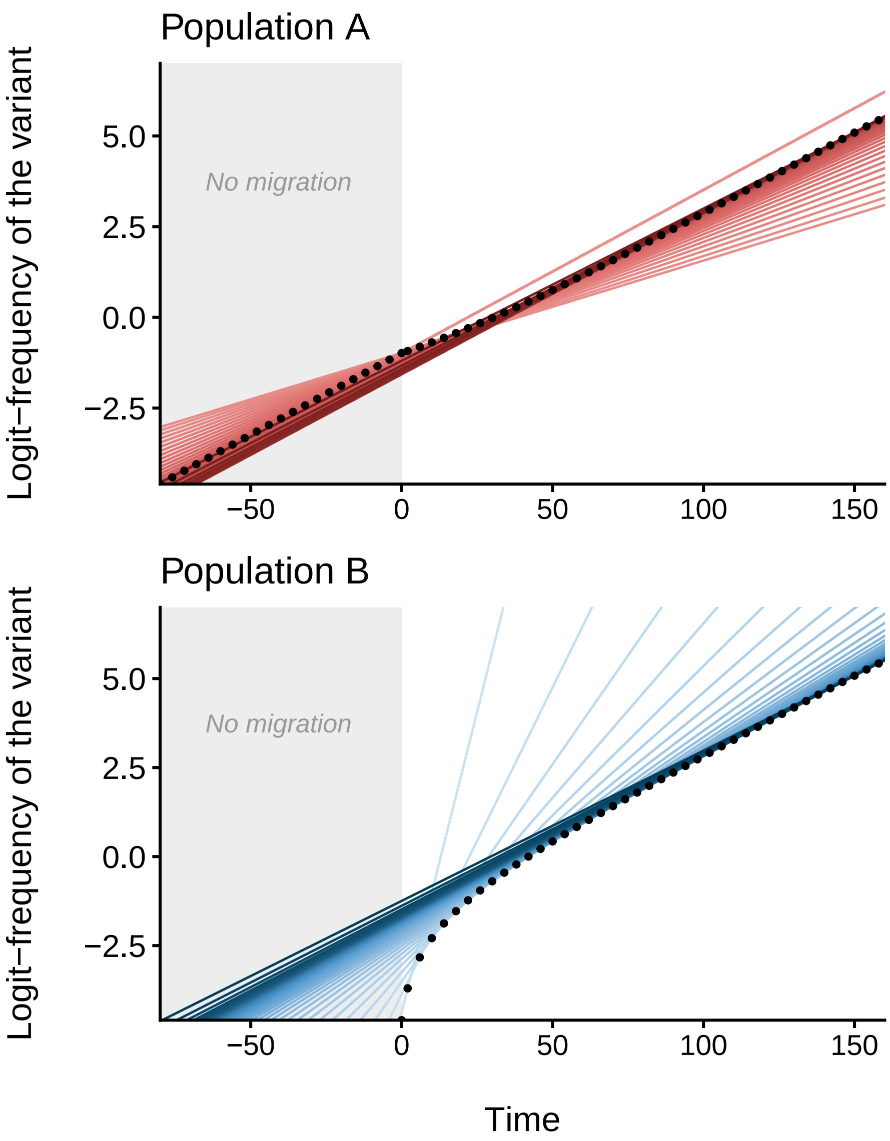


Figure S3: **Migration can steepen the change of local frequency of an emerging variant.** This figure is built upon the simulation presented in the main text in **Fig. 1**. The host metapopulation is divided in two populations, $A$ (top) and $B$ (bottom). The variant has a +30% transmission rate selective advantage over the wildtype. We simulate the model with identical total population densities ($N^A = N^B = 100$), and parameter values (same for both populations): $\beta_w = 0.15$, $\beta_m = 0.195$, $\gamma_w = \gamma_m = 0.1$ and $\omega = 0$. The wildtype is initially introduced at very low density ($10^{-7}$) in both populations and the variant is only introduced in population $A$ such that it represents 1% of the infected hosts $I^A$ – both populations are otherwise fully susceptible. Before $t = 0$, the two populations are isolated ($\mu^A = \mu^B = 0$, gray background). At $t = 0$, the variant is introduced in population $B$ (1% of $I^B$) and, from that time point onward, hosts from population $B$ may visit population $A$ ($\mu^B = 0.1$). Tangents (red and blue lines) to each logit-frequency values (black points) indicate the corresponding current rate of change. Tangent slopes are approximated using equations (1)-(2) from the main text; color shades (from lighter to darker) represent the course of time.

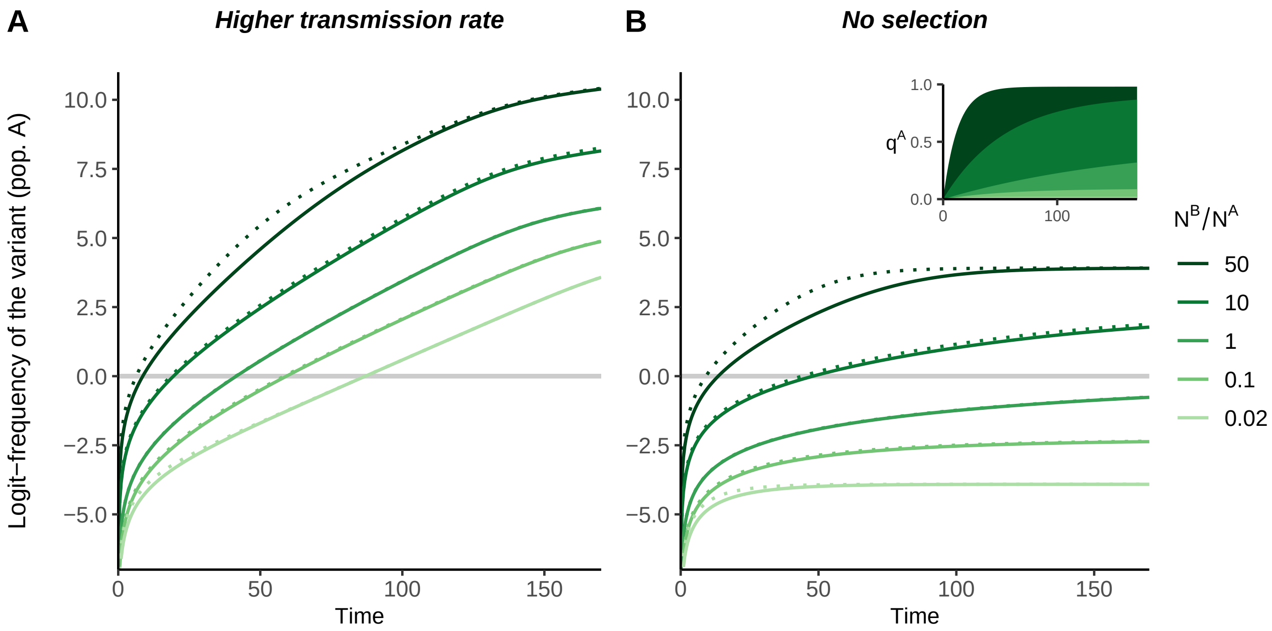


Figure S4: **The effect of population size asymmetry on the dynamics of the variant frequency in the focal population.** The host metapopulation is divided in two populations, $A$ and $B$. Varying the total population density ratio $N^B/N^A$ (in all cases, $N^A = 100$), we simulate our full model (**SI Appendix §S2.2**, system (S5), solid lines) along with our first-order approximation under weak migration (dotted lines). Trajectories of the logit-frequency of a variant under: (A) positive selection (+30% transmission rate, $\beta_m > \beta_w \Rightarrow \Delta\beta > 0$), (B) neutral evolution ($\beta_m = \beta_w \Rightarrow \Delta\beta = 0$). In all simulations, we take (same for both populations): $\beta_w = 0.15$, $\gamma_w = \gamma_m = 0.1$, $\mu^A = \mu^B = 10^{-2}$ and $\omega = 0$. At $t = 0$, the wildtype is introduced at very low prevalence in population $A$ ($I_w^A/N^A = 10^{-6}$ and $I_m^A = 0$) and the variant is introduced at the same prevalence in population $B$ ($I_m^B/N^B = 10^{-6}$ and $I_w^B = 0$) – both populations are otherwise fully susceptible.

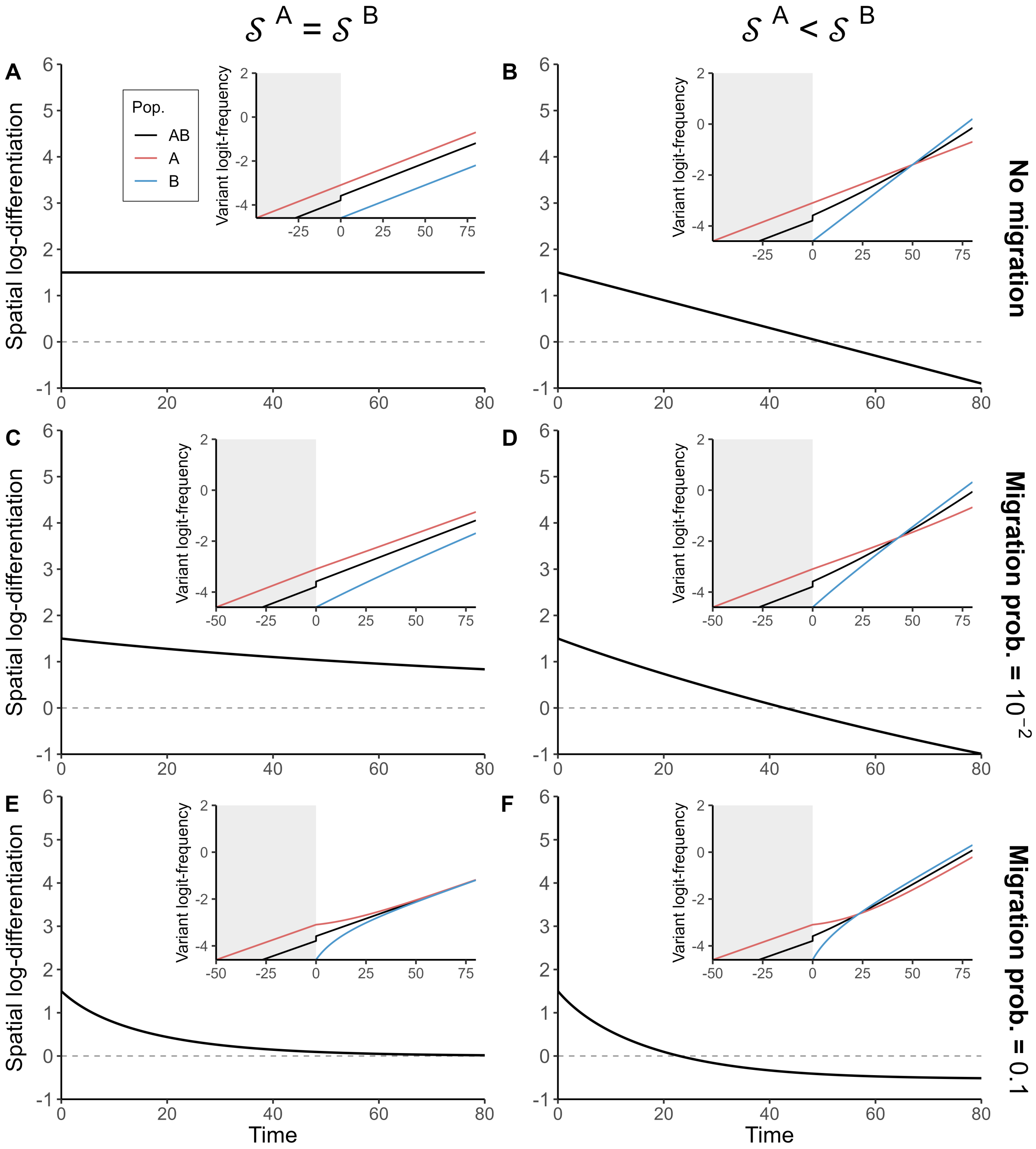


Figure S5: **Short-term dynamics of spatial differentiation and the migration–selection balance.** For the sake of simplicity, we assume that both strains share the same transmissibility. This allows us to set aside the dynamical effects in the selection coefficient. Fixed parameter values are: $\omega = 0$ and $\forall i \in \{A, B\},\ N^i = 100$, $\beta_w^i = \beta_m^i = 0.15$ and $\gamma_w^i = 0.1$. (A-E) The recovery rate of the variant do not vary across populations (homogeneous selection, $\mathcal{S}^A = \mathcal{S}^B$): $\forall i \in \{A, B\},\ \gamma_m^i = 0.07$ ($\mathcal{S}^i = -\Delta\gamma^i = 0.03$). (B-F) The variant is more adapted in population $B$ (heterogeneous selection, $\mathcal{S}^A < \mathcal{S}^B$): $\gamma_m^A = 0.07$ ($\mathcal{S}^A = -\Delta\gamma^A = 0.03$) and $\gamma_m^B = 0.04$ ($\mathcal{S}^B = -\Delta\gamma^B = 0.06$). The wildtype is initially introduced in both populations at the same density ($10^{-4}$) and the variant is only introduced in population $A$ such that it represents 1% of infected hosts $I^A$ – both populations are otherwise fully susceptible. Until $t = 0$, the two populations are isolated ($\mu^A = \mu^B = 0$) and the variant is only present in population $A$ (gray background). At $t = 0$, the variant is introduced in population $B$ at low density (1% of $I^B$) and the two populations (A-B) stay isolated, or are coupled with (C-D) low or (E-F) higher levels of migration. Note that epidemiological dynamics exhibit only little variations over the simulation period.

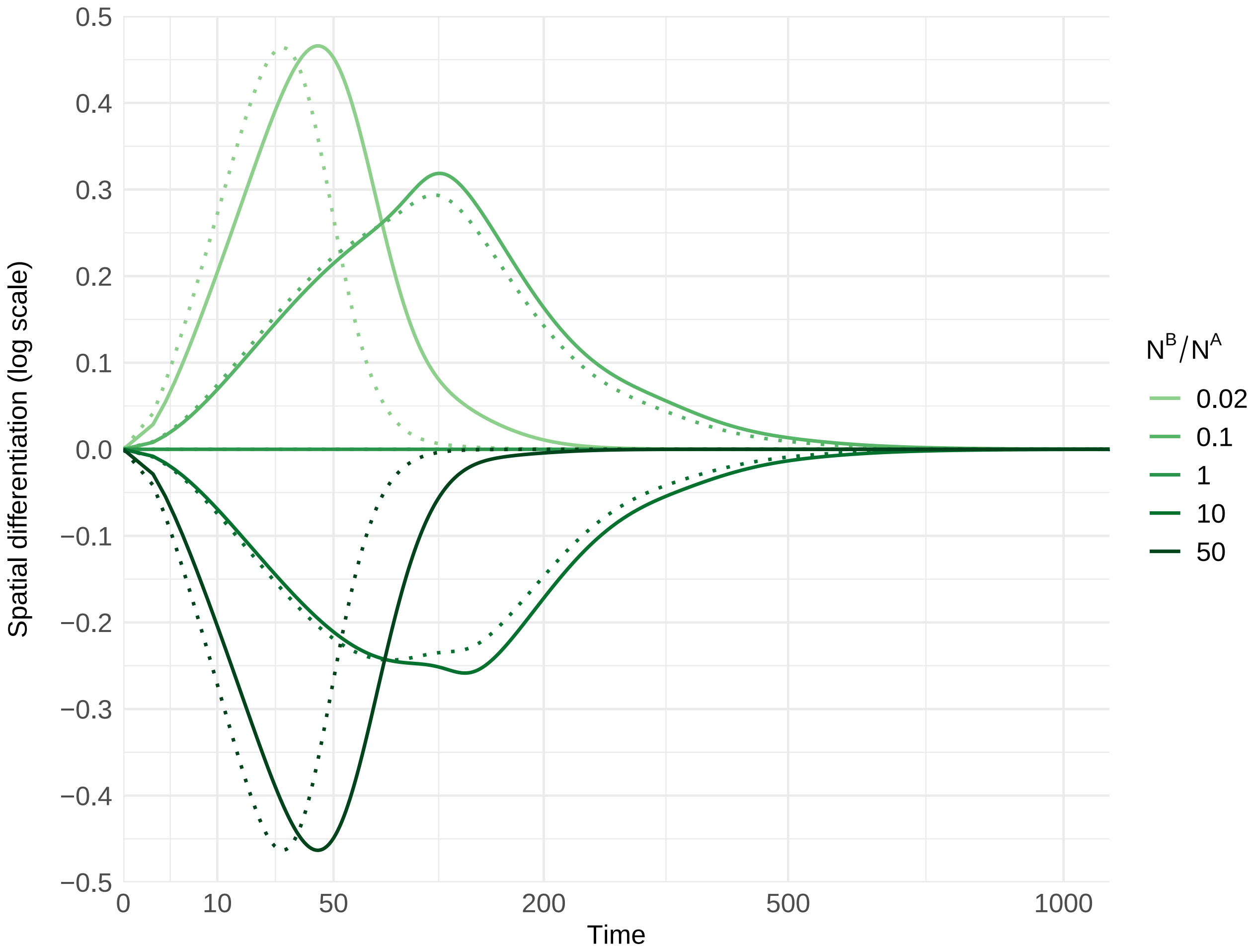


Figure S6: **The transient disruption of spatial differentiation.** The host metapopulation is divided in two populations, $A$ and $B$. Varying the total population density ratio $N^B/N^A$ (in all cases, $N^A = 100$), we simulate our full model (**SI Appendix §S2.2**, system (S5), solid lines) with parameter values (same for both populations): $\beta_w = 0.15$, $\beta_m = 0.195$ (+30% transmission rate, $\Delta\beta = 0.045$), $\gamma_w = \gamma_m = 0.1$ ($\Delta\gamma = 0$), $\omega = 0.01$ and $\mu^A = \mu^B = 10^{-2}$. We also run these simulations using our first-order approximation under weak migration (dotted lines). At $t = 0$, the wildtype and the variant are introduced at the same low *density* in both populations ($\forall i \in \{A, B\}$, $I_w^i = I_m^i = 10^{-3}$) – both populations are otherwise fully susceptible. The log-differentiation $\ln(\mathcal{Q})$ is then calculated as $\ln(\mathcal{Q}) = \text{logit}\left(q^A\right) - \text{logit}\left(q^B\right)$. Although there is no spatial differentiation initially ($\ln(\mathcal{Q}) = 0$), the spatial differentiation is then transiently disrupted as soon as $N^A \neq N^B$ because the initial differential in the proportions of infected and susceptible hosts results in a differential in selection when $\Delta\beta \neq 0$.

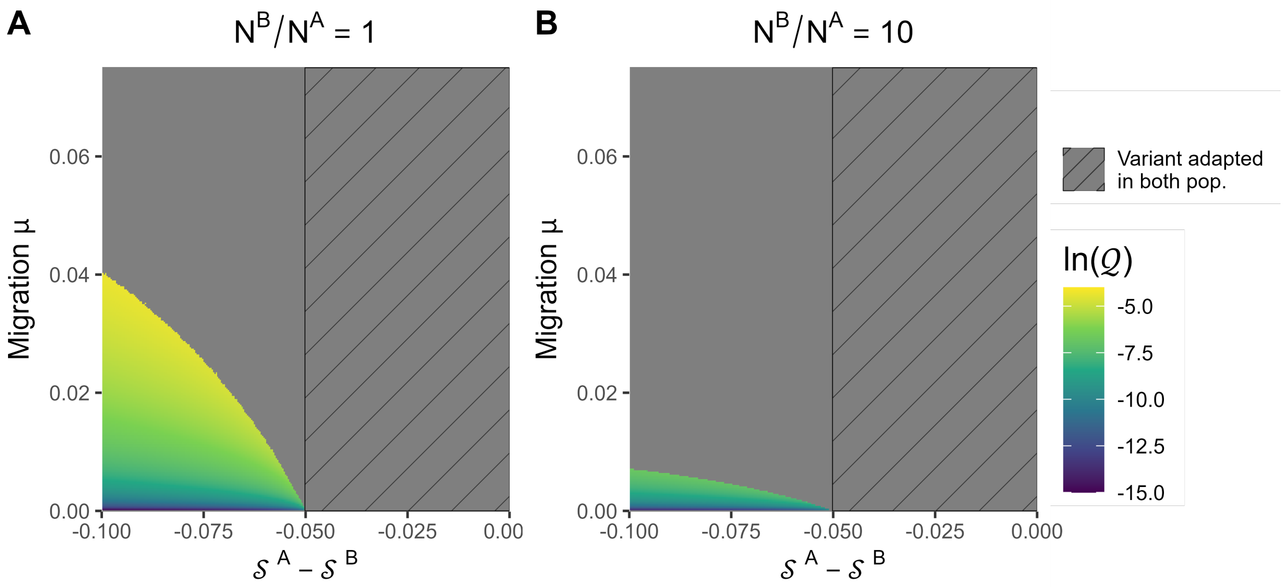


Figure S7: **Long-term spatial differentiation and local adaptation.** For the sake of simplicity, we assume that both strains share the same transmissibility and only differ in terms of recovery rates. This allows us to set aside the dynamical effects in the selection coefficients such that the difference $\mathcal{S}^A - \mathcal{S}^B$ reduces to $-\Delta\gamma^A + \Delta\gamma^B$. Parameter values shared by both populations are: $\beta_w = \beta_m = 0.15$, $\gamma_w = 0.1$, $\gamma_m^B = 0.05$ and $\omega = 0.01$; the variant is thus selected for in population $B$ ($\mathcal{S}^B = -\Delta\gamma^B > 0$). We compute the long-term spatial differentiation by varying the parameter value of $\gamma_m^A$ from 0.05 to 0.15 and of the migration probability $\mu = \mu^A = \mu^B$ from 0 to 0.075. In population $A$, depending on the value of $\gamma_m^A$, the variant is either selected for (hatching background, $\mathcal{S}^A = -\Delta\gamma^A > 0$) or against ($\mathcal{S}^A = -\Delta\gamma^A < 0$). Simulations were performed with (A) $N^A = N^B = 100$ or (B) $N^A = 100$ and $N^B = 1000$. The gray color corresponds to cases where a strain has numerically reached fixation in a population.

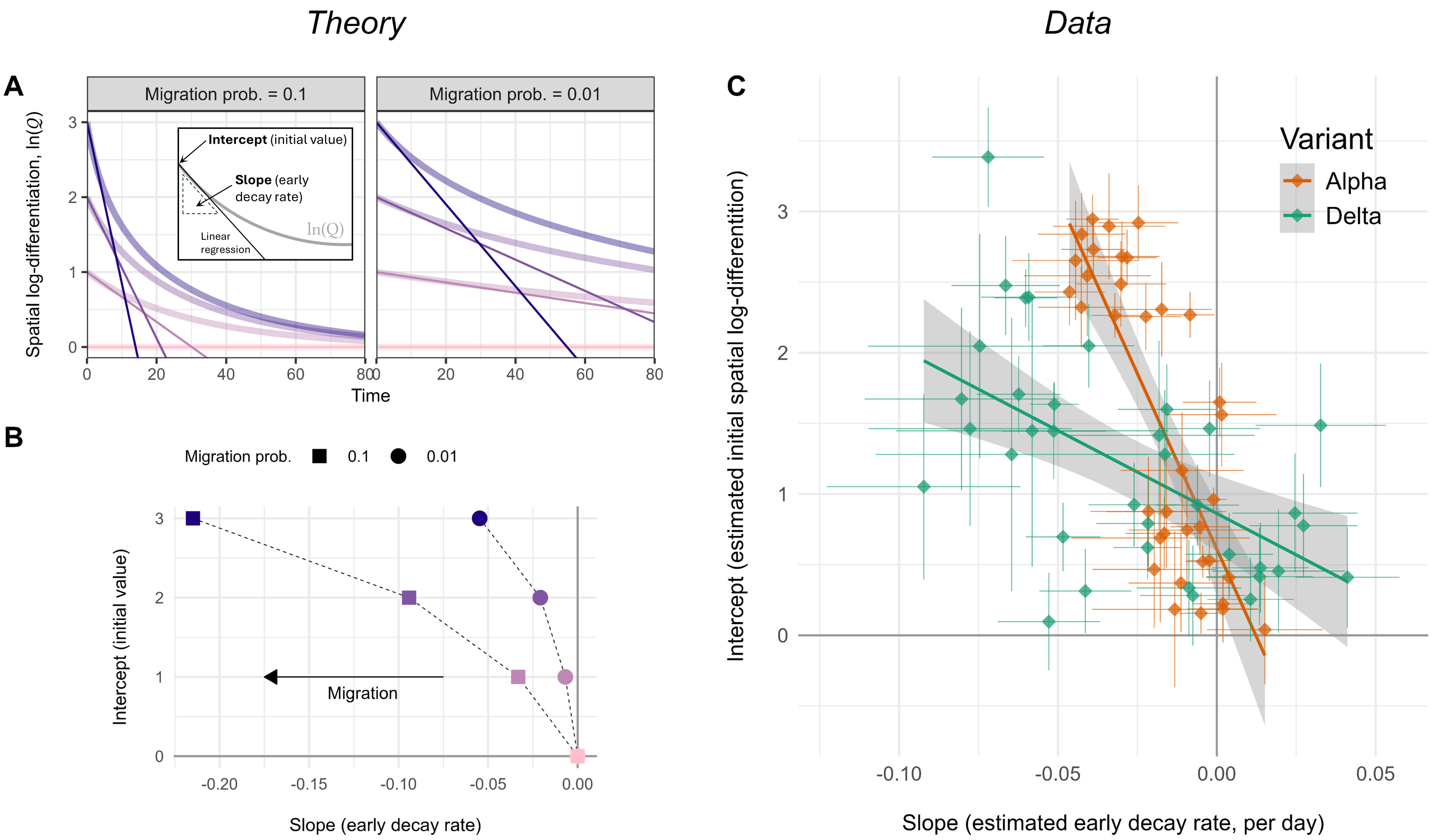


Figure S8: **Approximations of the early dynamics of the spatial differentiation.** (A-B) 'Theory' panel. According to equation (3), the dynamics of the spatial differentiation $\mathcal{Q}$ depends on the differential in selection between the two populations, the current level of differentiation and the intensity of migration. In the following we assume that selection is homogeneous across populations and we only focus on the last two. (A) Dynamics of the spatial log-differentiation $\ln(\mathcal{Q}) = \text{logit}\left(q^A\right) - \text{logit}\left(q^B\right)$ starting from different initial conditions and with two level of migration ($\mu^A = \mu^B = 0.01$ or $0.1$) – parameter values (same for all populations): $\beta_w = \beta_m = 0.15$, $\gamma_w = 0.1$ and $\gamma_m = 0.05$. Straight lines represent linear approximations of the early dynamics of $\ln(\mathcal{Q})$ and we reported the corresponding slopes and intercepts in panel B. (C) 'Data' panel. We estimated the early dynamics of the spatial log-differentiation (slope and intercept) of the Alpha and Delta variants using simple linear regressions. We used the data and the pairs of regions presented in **Fig. 4-C** and **D** (using daily data for Delta) for which elapsed time did not exceed 35 days. We oriented each pair so that the point estimate of the intercept was always positive. Interestingly, the patterns we obtain for SARS-CoV-2 are consistent with the hypothesis that migration was higher during the sweep of Delta because the drop in differentiation is steeper than for the Alpha variant (straight lines and gray envelopes show a linear regression on point estimates).

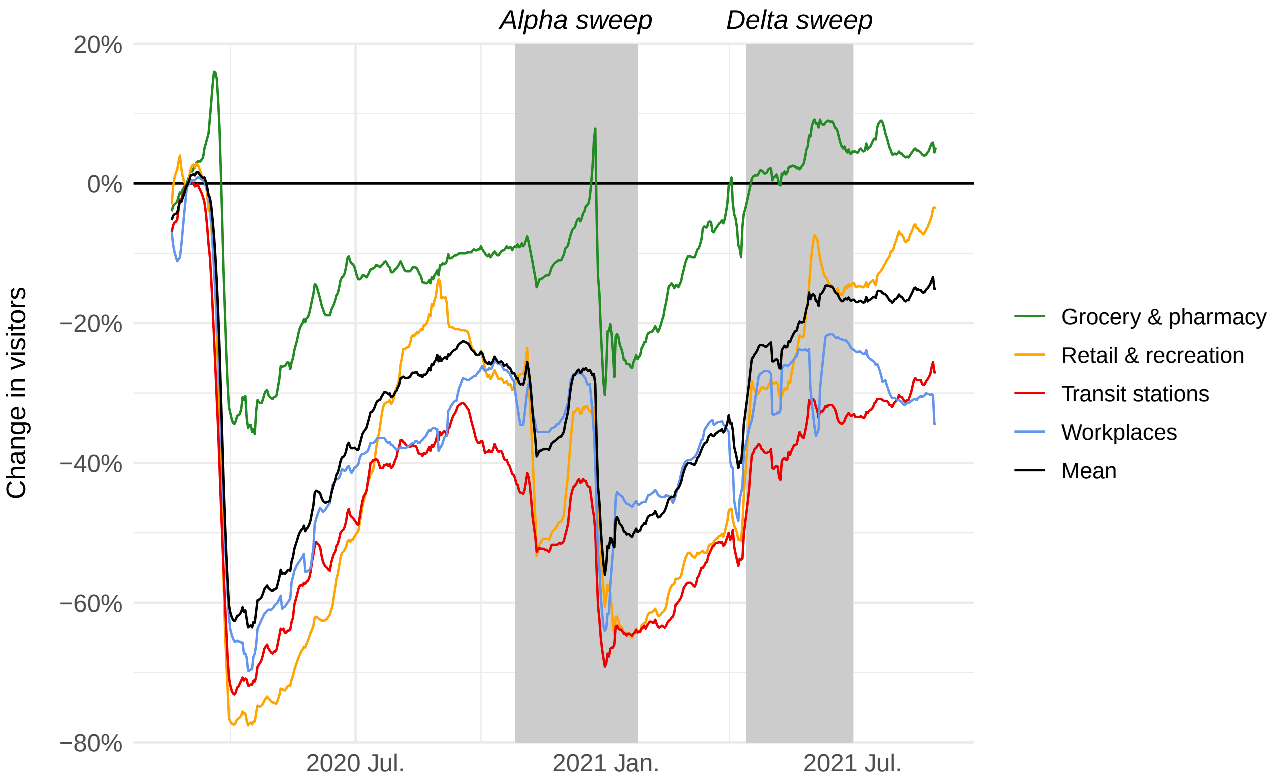


Figure S9: **Change in visitors in the United Kingdom (Google mobility reports).** Changes in the number of visitors compared to baseline days (median value from the 5-week period from Jan. 3 to Feb. 6, 2020). Mobility data were downloaded from the website *Our World in Data* (https://ourworldindata.org/covid-mobility-trends, last visited 2025-10-10) and we only focused on the following locations: grocery & pharmacy, retail & recreation, transit stations and workplaces (the mean across these four place categories is shown in black). The gray rectangles indicate the period of the sweep of the Alpha and Delta variants.

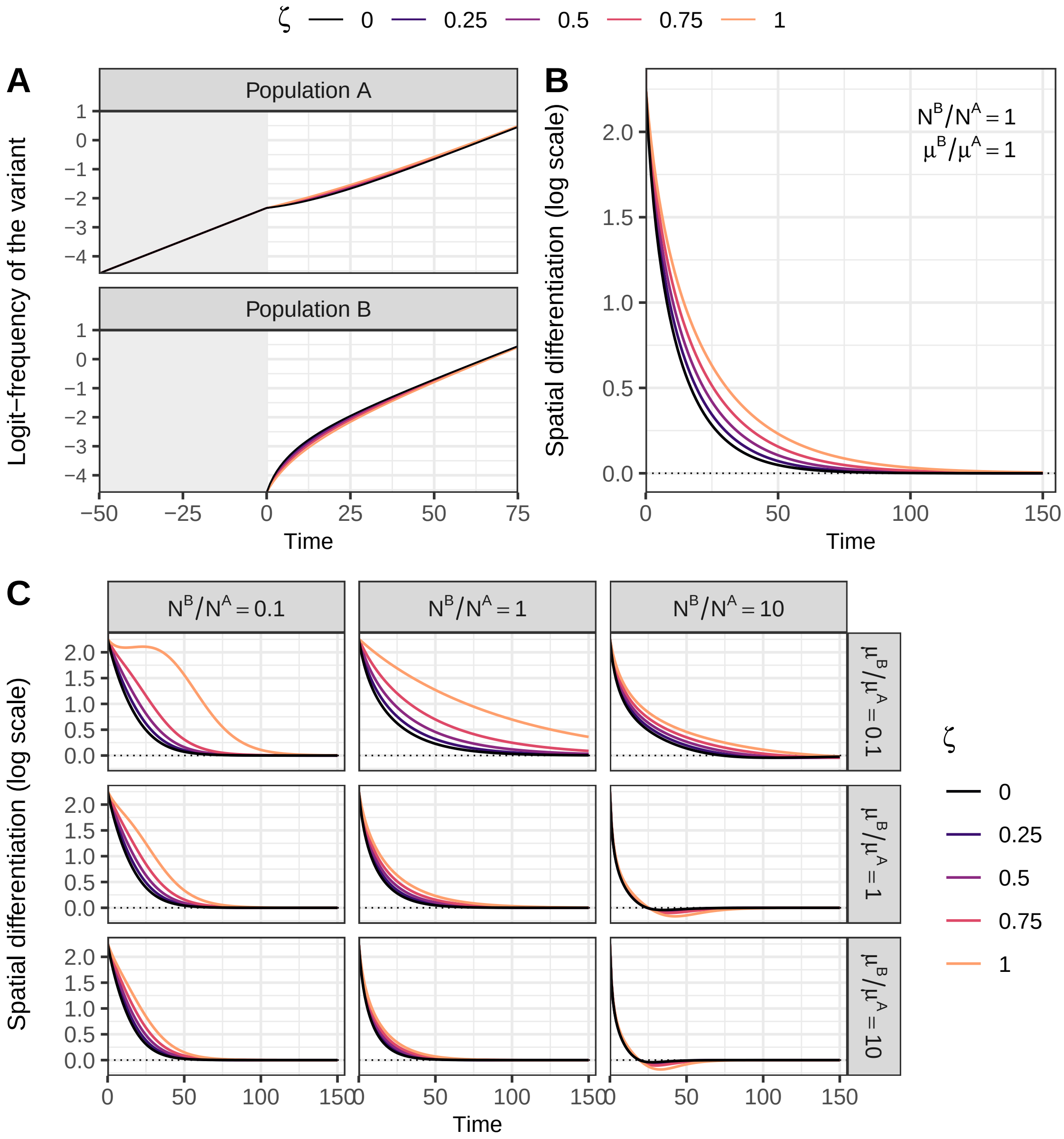


Figure S10: **The effect of reduced mobility in infected hosts on the evolutionary dynamics.** The host metapopulation is divided in two populations, $A$ and $B$. The variant has a +30% transmission rate selective advantage over the wildtype. We simulate the model with identical total population densities ($N^A = N^B = 100$), and parameter values (same for both populations): $\beta_w = 0.15$, $\beta_m = 0.195$, $\gamma_w = \gamma_m = 0.1$ and $\omega = 0$. The wildtype is initially introduced at very low density ($10^{-7}$) in both populations and the variant is only introduced in population $A$ such that it represents 1% of the infected hosts $I^A$ – both populations are otherwise fully susceptible. Before $t = 0$, the two populations are isolated ($\mu^A = \mu^B = 0$, gray background). At $t = 0$, the variant is introduced in population $B$ (1% of $I^B$) and, from that time point onward, hosts may visit the other population ($\mu^A = \mu^B = 0.1$). In **SI Appendix §S6.2**, we relax the assumption that infected hosts ($I$) commute with the same probability that non-infected hosts ($S$ and $R$), such that $\zeta$ represents the reduction in host mobility due to the disease, ranging from 0 (no impact, same as in the main text) to 1 (no mobility of infected hosts). Varying the value of $\zeta$, we plot the temporal dynamics of (A) the variant logit-frequency and of (B) the spatial log-differentiation $\ln(\mathcal{Q}) = \text{logit}\left(q^A\right) - \text{logit}\left(q^B\right)$. In (C), we focus on the dynamics of $\ln(\mathcal{Q})$, varying also population size and migration asymmetries between the two populations.
